# Interactional decision-making processes and communication on aid in dying: an international scoping review

**DOI:** 10.1101/2025.04.07.25325321

**Authors:** Claudia Gugel, Zoe Henning, Pola Hahlweg

## Abstract

**Background and objectives:** Aid in dying (AID) means assisting a person in their voluntary death. Despite a trend toward AID legalization, knowledge on how AID decisions (should) unfold is limited. We synthesized the current scientific understanding of interactional AID decision-making processes in the healthcare context.

**Methods:** For this scoping review, we searched Web of Science, PubMed, and CINAHL. Records were that reported own results on interactional AID decision-making processes in healthcare were eligible. Study designs or populations were not restricted. Data was extracted and synthesized with a structured excel sheet by three researchers.

**Results:** We identified 109 eligible records with diverse study designs and populations. They mentioned various participants in interactional AID decision-making processes, some specifically trained. Organizational and system characteristics also influenced such processes. Records described a longitudinal and iterative process, including several phases (beginning, assessment, preparation, realization, aftercare) and overarching themes (coordination, patient-centered care, inter-/multidisciplinary teamwork). A range of decisions were identified that need to be made by different stakeholders throughout the process. This complex process requires appropriate communication skills. Examples of facilitators were a trusting pre-existing patient-clinician-relationship, guidelines, and an open, non-judgmental attitude. Examples of challenges were power imbalances, clinicians’ subjective interpretations, and insufficient self-reflection. Future research to better understand and improve AID processes was called for.

**Conclusion:** Several publications explored interactional AID decision-making processes. Our comprehensive synthesis is a start to painting a comprehensive picture, e.g. in a scientific model. The knowledge gained can structure future AID research and inform implementation efforts.

**HIGHLIGHTS:**

– We synthesized the scientific understanding of interactional decision-making processes on aid in dying in the healthcare context from 109 publications.
– We synthesized across countries, patient and health care professional populations, settings, and study designs.
– Interactional decision-making processes on aid in dying are highly complex and can involve various participants.
– Interactional decision-making processes on aid in dying are a longitudinal and iterative process with several phases.
– A range of decisions were identified to be made by different stakeholders throughout the process.

## INTRODUCTION

While there is a trend towards legalizing aid in dying (AID) in many countries worldwide, attitudes, legal regulations, and implementation in practice remain manifold and complex (Emanuel et al., 2016; Mroz et al., 2021). The phenomenon of interest of this scoping review was interactional decision-making processes about AID in healthcare. AID decisions are highly consequential. However, many open questions remain. For example, how decision-making processes around AID unfold or what a high-quality AID decision should entail.

As terminology around AID is not used and understood uniformly (Mroz et al., 2021), we make some clarifications in the following. We understand AID as one person seeking to voluntarily ending their life and receiving assistance from another person in doing so. There are various synonymous or similar terms to AID such as assistance in dying/death, medical aid/assistance in dying, assisted suicide (AS), and (voluntary) euthanasia (VE). Depending on the country and legislature, these terms might differ in their meaning. AS has been defined as “a physician providing a patient who requests aid-in-dying a prescription that the patient can self-administer to end his or her life” (National Academies of Sciences Engineering and Medicine, 2018). There are also other realizations of AS such as attaching an infusion with the deadly medication and the person seeking to die opening the infusion. For AS, it is necessary that the person wanting to die is the person performing the act that leads to their death. On the other hand, VE means that “a physician administers lethal medication at the explicit request of the patient” (National Academies of Sciences Engineering and Medicine, 2018). In VE, the person assisting is acting on behalf of the person who wants to die. Furthermore, while these two definitions focus on physicians, a shift towards other health care professionals’ (HCP) assistance has been discussed in recent years (Stokes, 2017). In this publication, we used the terms AID, AS, and VE. We used AID as the umbrella term for both AS and VE and only referred to AS or VE if we explicitly exclude the other. As we focused on the healthcare context, we use the term patients for people seeking AID in this paper. For AID, communication and decision-making processes have been identified as important aspects (Brooks, 2019; Patel et al., 2021; Selby & Bean, 2019). In general, making a decision in healthcare is not one point in time, but a process. Much of it happens as interactional processes between people. For example, a shared decision-making process has been described as forming a decision-making team, sharing information both ways between patients and HCPs, eliciting preferences, and choosing an option (Elwyn et al., 2017). Overall, interactional decision-making processes in healthcare can be described as at least one patient and one HCP exchanging information and agreeing on a specific path forward regarding the patient’s state of health and medical care (Braddock et al., 1997; Ofstad et al., 2016). These interactional aspects are also essential in AID decisions, which was why we focused our scoping review on them. We did not look into aspects of AID decision-making that take place solely within one person’s mind.

Prior literature reviews have looked at experiences and roles in AID of HCPs from their perspective (e.g., Brooks (2019); Patel et al. (2021)) or of specific groups of HCPs such as oncologists, psychiatrists, psychologists, or speech-language pathologists (e.g., DeZeeuw and Myers (2020); Marina et al. (2021a); McCormack and Fléchais (2012); Selby and Bean (2019)). They often focused on legal aspects (e.g., Marina et al. (2021a); McCormack and Fléchais (2012); (Mroz et al., 2021)) or qualitative studies (e.g., Brooks (2019); Patel et al. (2021)). Another review looked in to the actual realization of AID (Zworth et al., 2020). Complementary, our study aimed to synthesize the current scientific understanding of interactional decision-making processes regarding AID (including AS and VE) in the health care context. We set out to do this comprehensively and broadly across patient populations, disciplines, settings, and study designs. We were guided by the following research questions:

- RQ1 What are the characteristic of studies that evaluated interactional decision-making processes on AID?
- RQ2 What is known about AID decision-making processes from scientific studies?

a. Who participates in AID decision-making processes?
b. What phases are described for AID decision-making processes?
c. What is known about aspects of communication and/or interaction during AID decision-making processes?
- RQ3 What research gaps and implications for future studies have been identified in scientific publications on AID decision-making processes?

## METHODS

### Study Design

We conducted a scoping review following the framework by Arksey and O’Malley (2005). We deemed a scoping review, which is “a form of knowledge synthesis that addresses an exploratory research question aimed at mapping key concepts, types of evidence, and gaps in research related to a defined area or field by systematically searching, selecting, and synthesizing existing knowledge” (Colquhoun et al., 2014), especially suitable for our research questions. We followed the Preferred Reporting Items for Systematic reviews and Meta-Analyses extension for Scoping Reviews (PRISMA-ScR) checklist (Tricco et al., 2018) (cp. Additional file 1). We used an internal review protocol and did not pre-register.

### Eligibility criteria

We sought publications that reported own results on interpersonal decision-making processes on AID in a professional healthcare context. A complete list of inclusion and exclusion criteria can be found in Table 1.

**Table 1.** Inclusion and exclusion criteria.

| **<u>Inclusion criteria</u>** |  |
| --- | --- |
| **1** | **The main subject or one of the main subjects of the publication is aid in dying (AID, including <u>assisted suicide</u> and/or <u>voluntary euthanasia</u>).**<br>*Definition of assisted suicide (AS): AS is a particular type of suicide. In AS, the person who wants to die is the person taking action when taking his/her own life. The suicide is supported by another person. Assistance of the third party can take different forms. It can lie in the preparation or execution of the suicide and express itself, among other things, in the provision of a lethal medication or in the procurement of opportunities. Physician-assisted suicide (PAS) is a subcategory of AS where the person supporting the suicide is a physician.*<br>*Definition of voluntary euthanasia (VE): Voluntary euthanasia means ending one's life by a medical professional administering a lethal medication at the person's explicit request. In voluntary euthanasia, the person taking action is the third party supporting the person who wants to die.*<br>*Note: Depending on the country and legislation, synonymous terms may occur: medical assistance in dying, medical aid in dying, etc. The terms "wish for hastened death"/"wish to hasten death" and "withholding treatment"/"withdrawing treatment" are not synonyms of AS/VE.* |
| **2** | **The publication reports on <u>decision-making processes</u> on AID, after the person decided to seek information about AID.**<br>*Definition of decision-making processes: Decision-making is a process resulting in the commitment to a belief or course of action chosen from more than one option (at least do or do not). During decision-making processes, individuals explore and incorporate different aspects such as knowledge/information, prior experiences, values, beliefs and attitudes, preferences, and/or emotions. The process of making a decision is understood as a longitudinal event and may involve interactional components. Interactional decision-making processes involve several steps: e.g., relationship building, information exchange, and choosing an option.* |
| **3** | **The publication reports on <u>interactions between people</u> being involved (i.e., person with a wish for hastened death, relatives, health care professionals (HCPs), others directly involved) during decision-making processes on AID.**<br>*Examples: Any interaction regarding information seeking/provision or counseling regarding AID; roles and functions of HCPs in the decision-making process on AID; behavioral and/or communicative reactions following requests for AID.* |
| **4** | **The publication reports on AID in a professional <u>health care context</u>.**<br>*This scoping review focuses on AID supported by professionals as opposed to support by family members or friends. The term "professional" is understood broadly and may include anyone dealing with AID in their professional capacities (e.g. HCPs, counselors, etc.).*<br>*Definition health care context: Health care context in this scoping review is understood broadly and may include any professional context concerned with communication about and/or requests for AID.* |
| **<u>Exclusion criteria</u>** |  |
| **1** | **The full-text is <u>not accessible</u>.** |
| 2 | **No author or no year** can be found for the full text. |
| 3 | The language of the full text is **not English or German**. |
| 4 | The publication is an opinion piece, commentary, editorial, letter, or study protocol that **does not report own results**. |
| 5 | **The publication reports on decision-making processes on AS/VE on a broader society level only.**<br>*Examples: political, ethical, or religious debate; legal matters; philosophical discussion of free will or voluntariness.*<br>*<u>This exclusion criterion does not include</u> publications that report on values and/or other ethical concepts applied to concrete situations of interactions between HCPs and people seeking AID.* |
| 6 | **The publication is a systematic review, a scoping review or a structured literature review.**<br>*Full texts excluded because of being reviews were reviewed for use in the secondary search strategy (reference tracking).* |

### Search strategy and information sources

We searched Web of Science, PubMed, and CINAHL on April 20^th^, 2022. We did not restrict the publication date and included articles published in English or German, as these were the languages spoken by the review team. Search strings for each database can be found in Additional file 2. As a secondary search strategy, literature review articles identified in the data base search were screened for relevant records in their reference list. In addition, we searched for annual country reports on AID on respective websites.

### Selection process

We imported all identified records into the reference management software Endnote and removed duplicates. For the title and abstract screening, each record was initially screened by one assessor. Subsequently, all records with uncertainty were discussed by all assessors and a consensus decision was made. Additional file 3 describes the process of title and abstract screening. For full-text eligibility assessment, we began by joint assessment of five records, followed by three rounds of double assessment of a limited number of records (5 to 10% of the data set each) to refine our understanding of the eligibility criteria and ensure sufficient inter-rater-reliability. Afterwards, the remaining full texts were assessed by one assessor each. In case of doubt, double or triple assessment was sought until a consensus about in- or exclusion was reached. A description of the complete full-text eligibility assessment process can be found in Additional file 4.

### Data charting process and synthesis of results

Guided by the aim and research questions of this study, a data extraction form was developed by the study team. The development was an iterative process combining team discussions and pilot testing of the extraction form. Data extracted included general information of the record, study characteristics, results regarding interactional AID decision-making processes (e.g., participants, phases, decisions to be made, content of communication), additional relevant aspects before and after the interactional decision-making process, future research, and relevance assessment by the study team. More detail on the development and use of the extraction form is given in Additional file 5. Initial data extraction was undertaken by CG and ZH. It was done as close to the original records as possible. Results were extracted, but not interpreted. In a second step, PH reviewed and revised the extracted data. This step included streamlining with respect to the research questions to allow further synthesis and interpretation. The final data extraction sheet can be found in Additional file 6.

### Consultation

To corroborate the interpretation and synthesis of the data set made by the study team, we presented and discussed our preliminary results in an expert workshop. We conducted this workshop with 17 participants in February 2023. The workshop was conducted online and participants came from seven out of 16 states in Germany. They were patient representatives (n=2), physicians with backgrounds in general medicine (n=2), palliative care (n=5), oncology (n=1), nurses (n=3), legal experts (n=1), clinical ethicists (n=3), chaplains (n=1), psychologists (n=1), and worked for AID organizations (n=2) (multiple may apply to one person). Five additional potential participants had to cancel with short notice due to patient care (n=3) or acute illness (n=2).

## RESULTS

### Included records and study characteristics (RQ1)

We included 109 records. The initial data base search had yielded 9,622 records, plus 49 additional country reports on AID. After removal of duplicates, 7,963 records remained. During title and abstract screening, we removed another 7,647 records, leaving 316 full-texts to be assessed. Figure 1 shows which criterions led to exclusion of an additional 207 records.

**Figure 1.**
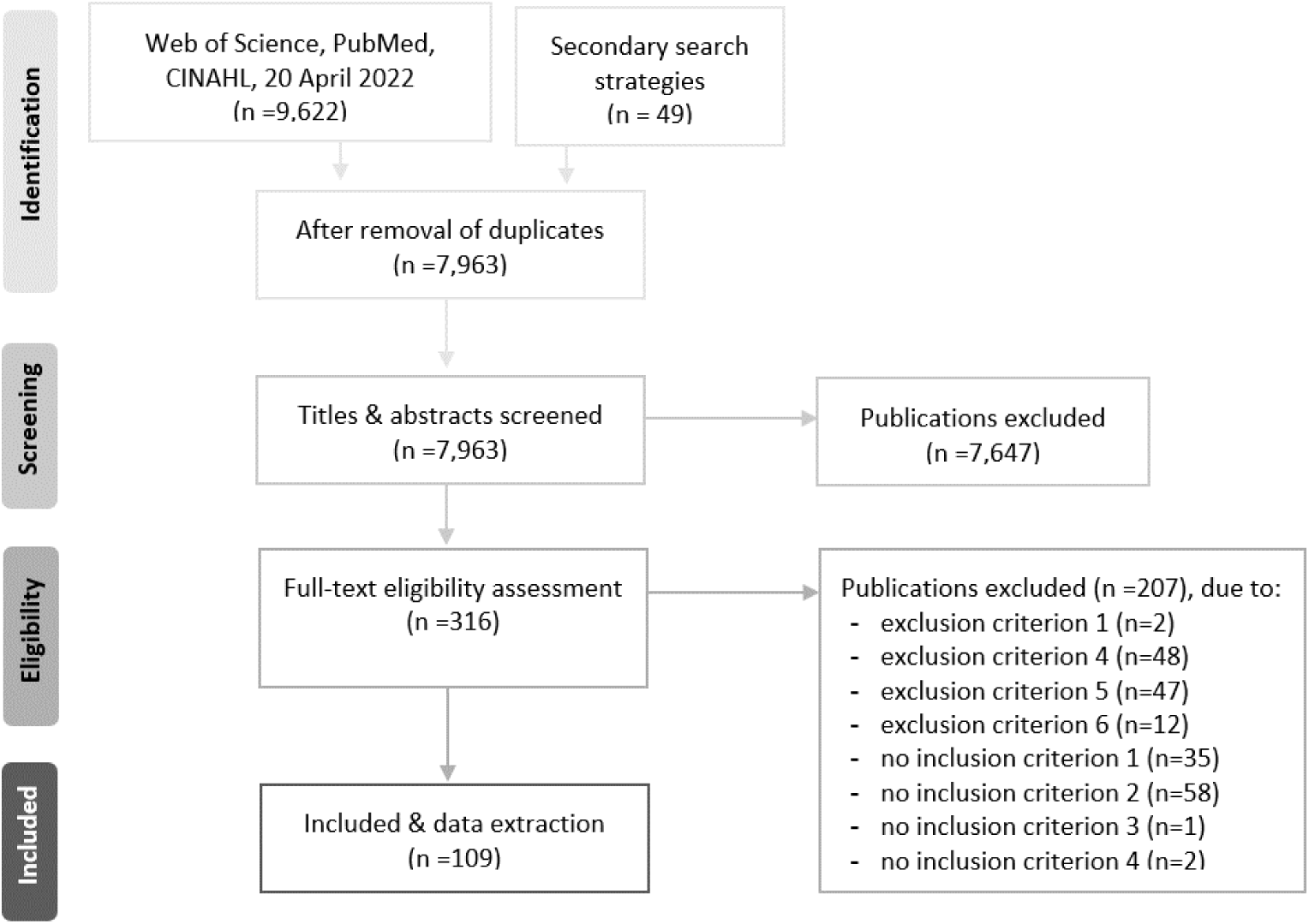
Flow chart.

Frequencies of records per time interval suggested an increase in the number of relevant records over time. The records reported from twelve countries, with 105 records giving insights from one country, three from two or three countries, and one not specifying the country. Included records most frequently studied patients (n=44, 40%) and/or physicians (n=27, 25%), with some reporting on more than one study population. The most often used study designs in our data set were qualitative interviews (n=42, 39%), case reports (n=20, 18.3), and quantitative surveys (n=19, 17%). In addition, study designs such as the description of one organization’s (implementation) process or guideline development had been facilitated. Table 2 gives an overview of selected study characteristics, for more detail consult Additional file 6.

**Table 2.**
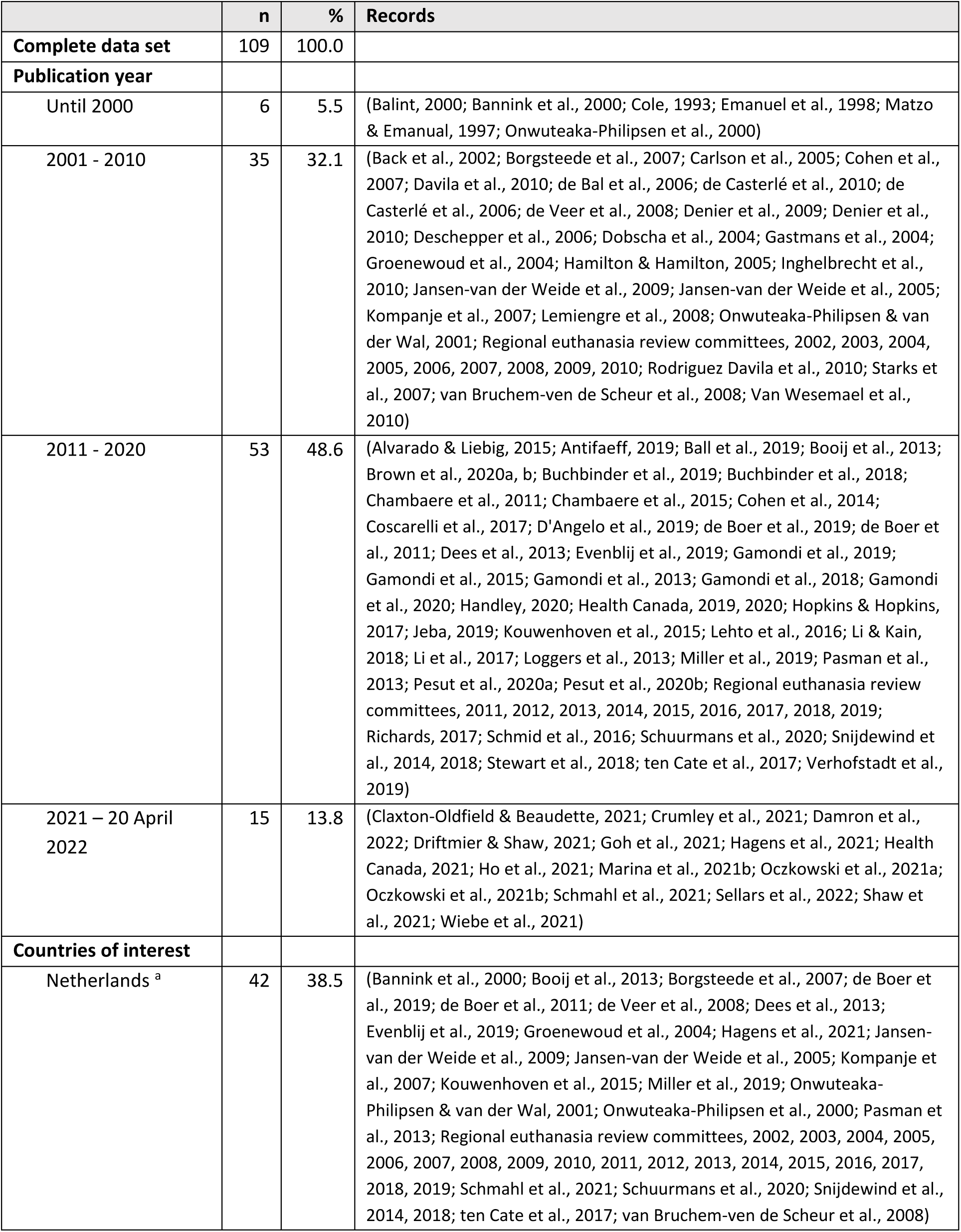
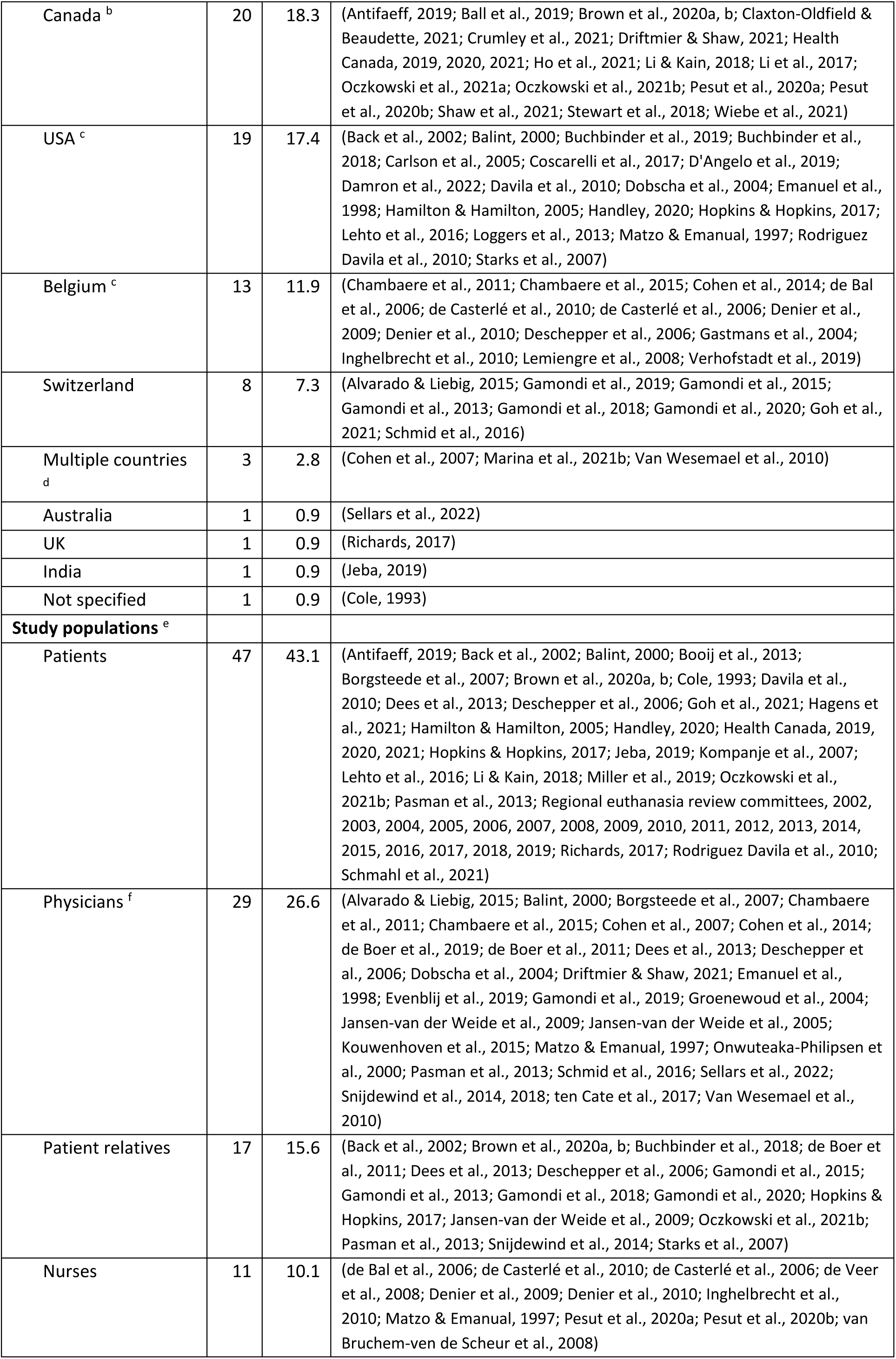
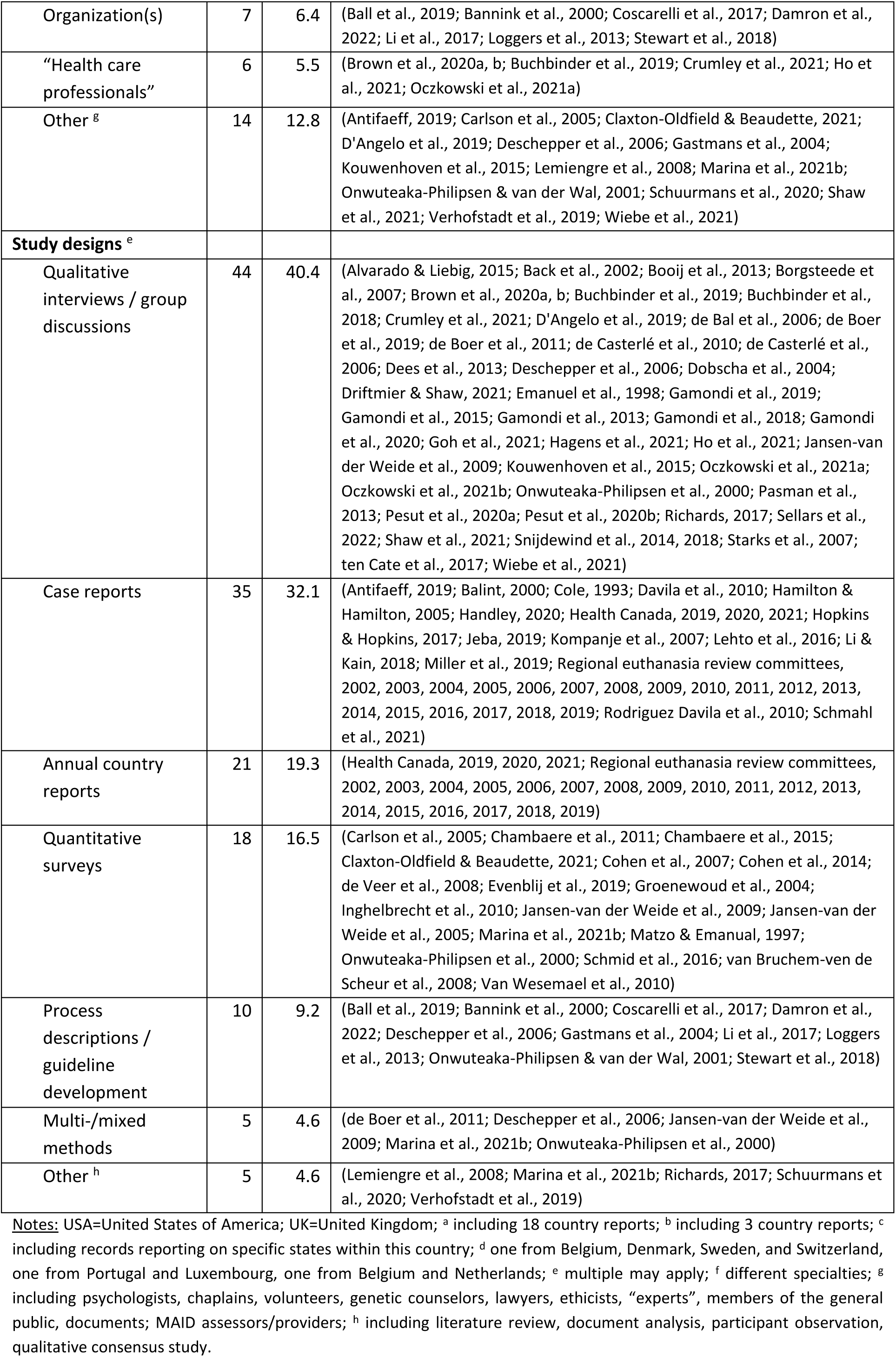
Characteristics of included records.

|  | n | % | Records |
| --- | --- | --- | --- |
| **Complete data set** | 109 | 100.0 |  |
| **Publication year** |  |  |  |
| Until 2000 | 6 | 5.5 | (Balint, 2000; Bannink et al., 2000; Cole, 1993; Emanuel et al., 1998; Matzo & Emanuel, 1997; Onwuteaka-Philipsen et al., 2000) |
| 2001 - 2010 | 35 | 32.1 | (Back et al., 2002; Borgsteede et al., 2007; Carlson et al., 2005; Cohen et al., 2007; Davila et al., 2010; de Bal et al., 2006; de Casterlé et al., 2010; de Casterlé et al., 2006; de Veer et al., 2008; Denier et al., 2009; Denier et al., 2010; Deschepper et al., 2006; Dobscha et al., 2004; Gastmans et al., 2004; Groenewoud et al., 2004; Hamilton & Hamilton, 2005; Inghelbrecht et al., 2010; Jansen-van der Weide et al., 2009; Jansen-van der Weide et al., 2005; Kompanje et al., 2007; Lemiengre et al., 2008; Onwuteaka-Philipsen & van der Wal, 2001; Regional euthanasia review committees, 2002, 2003, 2004, 2005, 2006, 2007, 2008, 2009, 2010; Rodriguez Davila et al., 2010; Starks et al., 2007; van Bruchem-ven de Scheur et al., 2008; Van Wesemael et al., 2010) |
| 2011 - 2020 | 53 | 48.6 | (Alvarado & Liebig, 2015; Antifaeff, 2019; Ball et al., 2019; Booij et al., 2013; Brown et al., 2020a, b; Buchbinder et al., 2019; Buchbinder et al., 2018; Chambaere et al., 2011; Chambaere et al., 2015; Cohen et al., 2014; Coscarelli et al., 2017; D'Angelo et al., 2019; de Boer et al., 2019; de Boer et al., 2011; Dees et al., 2013; Evenblij et al., 2019; Gamondi et al., 2019; Gamondi et al., 2015; Gamondi et al., 2013; Gamondi et al., 2018; Gamondi et al., 2020; Handley, 2020; Health Canada, 2019, 2020; Hopkins & Hopkins, 2017; Jeba, 2019; Kouwenhoven et al., 2015; Lehto et al., 2016; Li & Kain, 2018; Li et al., 2017; Loggers et al., 2013; Miller et al., 2019; Pasman et al., 2013; Pesut et al., 2020a; Pesut et al., 2020b; Regional euthanasia review committees, 2011, 2012, 2013, 2014, 2015, 2016, 2017, 2018, 2019; Richards, 2017; Schmid et al., 2016; Schuurmans et al., 2020; Snijdewind et al., 2014, 2018; Stewart et al., 2018; ten Cate et al., 2017; Verhofstadt et al., 2019) |
| 2021 – 20 April 2022 | 15 | 13.8 | (Claxton-Oldfield & Beaudette, 2021; Crumley et al., 2021; Damron et al., 2022; Driftmier & Shaw, 2021; Goh et al., 2021; Hagens et al., 2021; Health Canada, 2021; Ho et al., 2021; Marina et al., 2021b; Oczkowski et al., 2021a; Oczkowski et al., 2021b; Schmahl et al., 2021; Sellars et al., 2022; Shaw et al., 2021; Wiebe et al., 2021) |
| **Countries of interest** |  |  |  |
| Netherlands <sup>a</sup> | 42 | 38.5 | (Bannink et al., 2000; Booij et al., 2013; Borgsteede et al., 2007; de Boer et al., 2019; de Boer et al., 2011; de Veer et al., 2008; Dees et al., 2013; Evenblij et al., 2019; Groenewoud et al., 2004; Hagens et al., 2021; Jansen-van der Weide et al., 2009; Jansen-van der Weide et al., 2005; Kompanje et al., 2007; Kouwenhoven et al., 2015; Miller et al., 2019; Onwuteaka-Philipsen & van der Wal, 2001; Onwuteaka-Philipsen et al., 2000; Pasman et al., 2013; Regional euthanasia review committees, 2002, 2003, 2004, 2005, 2006, 2007, 2008, 2009, 2010, 2011, 2012, 2013, 2014, 2015, 2016, 2017, 2018, 2019; Schmahl et al., 2021; Schuurmans et al., 2020; Snijdewind et al., 2014, 2018; ten Cate et al., 2017; van Bruchem-ven de Scheur et al., 2008) |
| Canada <sup>b</sup> | 20 | 18.3 | (Antifaef, 2019; Ball et al., 2019; Brown et al., 2020a, b; Claxton-Oldfield & Beaudette, 2021; Crumley et al., 2021; Driftmier & Shaw, 2021; Health Canada, 2019, 2020, 2021; Ho et al., 2021; Li & Kain, 2018; Li et al., 2017; Oczkowski et al., 2021a; Oczkowski et al., 2021b; Pesut et al., 2020a; Pesut et al., 2020b; Shaw et al., 2021; Stewart et al., 2018; Wiebe et al., 2021) |
| USA <sup>c</sup> | 19 | 17.4 | (Back et al., 2002; Balint, 2000; Buchbinder et al., 2019; Buchbinder et al., 2018; Carlson et al., 2005; Coscarelli et al., 2017; D'Angelo et al., 2019; Damron et al., 2022; Davila et al., 2010; Dobscha et al., 2004; Emanuel et al., 1998; Hamilton & Hamilton, 2005; Handley, 2020; Hopkins & Hopkins, 2017; Lehto et al., 2016; Loggers et al., 2013; Matzo & Emanuel, 1997; Rodriguez Davila et al., 2010; Starks et al., 2007) |
| Belgium <sup>c</sup> | 13 | 11.9 | (Chambaere et al., 2011; Chambaere et al., 2015; Cohen et al., 2014; de Bal et al., 2006; de Casterlé et al., 2010; de Casterlé et al., 2006; Denier et al., 2009; Denier et al., 2010; Deschepper et al., 2006; Gastmans et al., 2004; Inghelbrecht et al., 2010; Lemiengre et al., 2008; Verhofstadt et al., 2019) |
| Switzerland | 8 | 7.3 | (Alvarado & Liebig, 2015; Gamondi et al., 2019; Gamondi et al., 2015; Gamondi et al., 2013; Gamondi et al., 2018; Gamondi et al., 2020; Goh et al., 2021; Schmid et al., 2016) |
| Multiple countries <sup>d</sup> | 3 | 2.8 | (Cohen et al., 2007; Marina et al., 2021b; Van Wesemael et al., 2010) |
| Australia | 1 | 0.9 | (Sellars et al., 2022) |
| UK | 1 | 0.9 | (Richards, 2017) |
| India | 1 | 0.9 | (Jeba, 2019) |
| Not specified | 1 | 0.9 | (Cole, 1993) |
| **Study populations <sup>e</sup>** |  |  |  |
| Patients | 47 | 43.1 | (Antifaef, 2019; Back et al., 2002; Balint, 2000; Booij et al., 2013; Borgsteede et al., 2007; Brown et al., 2020a, b; Cole, 1993; Davila et al., 2010; Dees et al., 2013; Deschepper et al., 2006; Goh et al., 2021; Hagens et al., 2021; Hamilton & Hamilton, 2005; Handley, 2020; Health Canada, 2019, 2020, 2021; Hopkins & Hopkins, 2017; Jeba, 2019; Kompanje et al., 2007; Lehto et al., 2016; Li & Kain, 2018; Miller et al., 2019; Oczkowski et al., 2021b; Pasman et al., 2013; Regional euthanasia review committees, 2002, 2003, 2004, 2005, 2006, 2007, 2008, 2009, 2010, 2011, 2012, 2013, 2014, 2015, 2016, 2017, 2018, 2019; Richards, 2017; Rodriguez Davila et al., 2010; Schmahl et al., 2021) |
| Physicians <sup>f</sup> | 29 | 26.6 | (Alvarado & Liebig, 2015; Balint, 2000; Borgsteede et al., 2007; Chambaere et al., 2011; Chambaere et al., 2015; Cohen et al., 2007; Cohen et al., 2014; de Boer et al., 2019; de Boer et al., 2011; Dees et al., 2013; Deschepper et al., 2006; Dobscha et al., 2004; Driftmier & Shaw, 2021; Emanuel et al., 1998; Evenblij et al., 2019; Gamondi et al., 2019; Groenewoud et al., 2004; Jansen-van der Weide et al., 2009; Jansen-van der Weide et al., 2005; Kouwenhoven et al., 2015; Matzo & Emanuel, 1997; Onwuteaka-Philipsen et al., 2000; Pasman et al., 2013; Schmid et al., 2016; Sellars et al., 2022; Snijdewind et al., 2014, 2018; ten Cate et al., 2017; Van Wesemael et al., 2010) |
| Patient relatives | 17 | 15.6 | (Back et al., 2002; Brown et al., 2020a, b; Buchbinder et al., 2018; de Boer et al., 2011; Dees et al., 2013; Deschepper et al., 2006; Gamondi et al., 2015; Gamondi et al., 2013; Gamondi et al., 2018; Gamondi et al., 2020; Hopkins & Hopkins, 2017; Jansen-van der Weide et al., 2009; Oczkowski et al., 2021b; Pasman et al., 2013; Snijdewind et al., 2014; Starks et al., 2007) |
| Nurses | 11 | 10.1 | (de Bal et al., 2006; de Casterlé et al., 2010; de Casterlé et al., 2006; de Veer et al., 2008; Denier et al., 2009; Denier et al., 2010; Inghelbrecht et al., 2010; Matzo & Emanuel, 1997; Pesut et al., 2020a; Pesut et al., 2020b; van Bruchem-ven de Scheur et al., 2008) |
| Organization(s) | 7 | 6.4 | (Ball et al., 2019; Bannink et al., 2000; Coscarelli et al., 2017; Damron et al., 2022; Li et al., 2017; Loggers et al., 2013; Stewart et al., 2018) |
| “Health care professionals” | 6 | 5.5 | (Brown et al., 2020a, b; Buchbinder et al., 2019; Crumley et al., 2021; Ho et al., 2021; Oczkowski et al., 2021a) |
| Other <sup>g</sup> | 14 | 12.8 | (Antifaef, 2019; Carlson et al., 2005; Claxton-Oldfield & Beaudette, 2021; D'Angelo et al., 2019; Deschepper et al., 2006; Gastmans et al., 2004; Kouwenhoven et al., 2015; Lemiengre et al., 2008; Marina et al., 2021b; Onwuteaka-Philipsen & van der Wal, 2001; Schuurmans et al., 2020; Shaw et al., 2021; Verhofstadt et al., 2019; Wiebe et al., 2021) |
| **Study designs <sup>e</sup>** |  |  |  |
| Qualitative interviews / group discussions | 44 | 40.4 | (Alvarado & Liebig, 2015; Back et al., 2002; Booij et al., 2013; Borgsteede et al., 2007; Brown et al., 2020a, b; Buchbinder et al., 2019; Buchbinder et al., 2018; Crumley et al., 2021; D'Angelo et al., 2019; de Bal et al., 2006; de Boer et al., 2019; de Boer et al., 2011; de Casterlé et al., 2010; de Casterlé et al., 2006; Dees et al., 2013; Deschepper et al., 2006; Dobscha et al., 2004; Driftmier & Shaw, 2021; Emanuel et al., 1998; Gamondi et al., 2019; Gamondi et al., 2015; Gamondi et al., 2013; Gamondi et al., 2018; Gamondi et al., 2020; Goh et al., 2021; Hagens et al., 2021; Ho et al., 2021; Jansen-van der Weide et al., 2009; Kouwenhoven et al., 2015; Oczkowski et al., 2021a; Oczkowski et al., 2021b; Onwuteaka-Philipsen et al., 2000; Pasman et al., 2013; Pesut et al., 2020a; Pesut et al., 2020b; Richards, 2017; Sellars et al., 2022; Shaw et al., 2021; Snijdewind et al., 2014, 2018; Starks et al., 2007; ten Cate et al., 2017; Wiebe et al., 2021) |
| Case reports | 35 | 32.1 | (Antifaef, 2019; Balint, 2000; Cole, 1993; Davila et al., 2010; Hamilton & Hamilton, 2005; Handley, 2020; Health Canada, 2019, 2020, 2021; Hopkins & Hopkins, 2017; Jeba, 2019; Kompanje et al., 2007; Lehto et al., 2016; Li & Kain, 2018; Miller et al., 2019; Regional euthanasia review committees, 2002, 2003, 2004, 2005, 2006, 2007, 2008, 2009, 2010, 2011, 2012, 2013, 2014, 2015, 2016, 2017, 2018, 2019; Rodriguez Davila et al., 2010; Schmahl et al., 2021) |
| Annual country reports | 21 | 19.3 | (Health Canada, 2019, 2020, 2021; Regional euthanasia review committees, 2002, 2003, 2004, 2005, 2006, 2007, 2008, 2009, 2010, 2011, 2012, 2013, 2014, 2015, 2016, 2017, 2018, 2019) |
| Quantitative surveys | 18 | 16.5 | (Carlson et al., 2005; Chambaere et al., 2011; Chambaere et al., 2015; Claxton-Oldfield & Beaudette, 2021; Cohen et al., 2007; Cohen et al., 2014; de Veer et al., 2008; Evenblij et al., 2019; Groenewoud et al., 2004; Inghelbrecht et al., 2010; Jansen-van der Weide et al., 2009; Jansen-van der Weide et al., 2005; Marina et al., 2021b; Matzo & Emanuel, 1997; Onwuteaka-Philipsen et al., 2000; Schmid et al., 2016; van Bruchem-ven de Scheur et al., 2008; Van Wesemael et al., 2010) |
| Process descriptions / guideline development | 10 | 9.2 | (Ball et al., 2019; Bannink et al., 2000; Coscarelli et al., 2017; Damron et al., 2022; Deschepper et al., 2006; Gastmans et al., 2004; Li et al., 2017; Loggers et al., 2013; Onwuteaka-Philipsen & van der Wal, 2001; Stewart et al., 2018) |
| Multi-/mixed methods | 5 | 4.6 | (de Boer et al., 2011; Deschepper et al., 2006; Jansen-van der Weide et al., 2009; Marina et al., 2021b; Onwuteaka-Philipsen et al., 2000) |
| Other <sup>h</sup> | 5 | 4.6 | (Lemiengre et al., 2008; Marina et al., 2021b; Richards, 2017; Schuurmans et al., 2020; Verhofstadt et al., 2019) |

**Notes:** USA=United States of America; UK=United Kingdom; <sup>a</sup> including 18 country reports; <sup>b</sup> including 3 country reports; <sup>c</sup> including records reporting on specific states within this country; <sup>d</sup> one from Belgium, Denmark, Sweden, and Switzerland, one from Portugal and Luxembourg, one from Belgium and Netherlands; <sup>e</sup> multiple may apply; <sup>f</sup> different specialties; <sup>g</sup> including psychologists, chaplains, volunteers, genetic counselors, lawyers, ethicists, “experts”, members of the general public, documents; MAID assessors/providers; <sup>h</sup> including literature review, document analysis, participant observation, qualitative consensus study.

### Participants of AID decision-making processes (RQ2)

Included records mentioned various groups of people that can be involved in interpersonal AID decision-making processes in healthcare. Figure 2 gives an overview of potential participants identified; Additional file 6 gives full details for each record.

**Figure 2.**
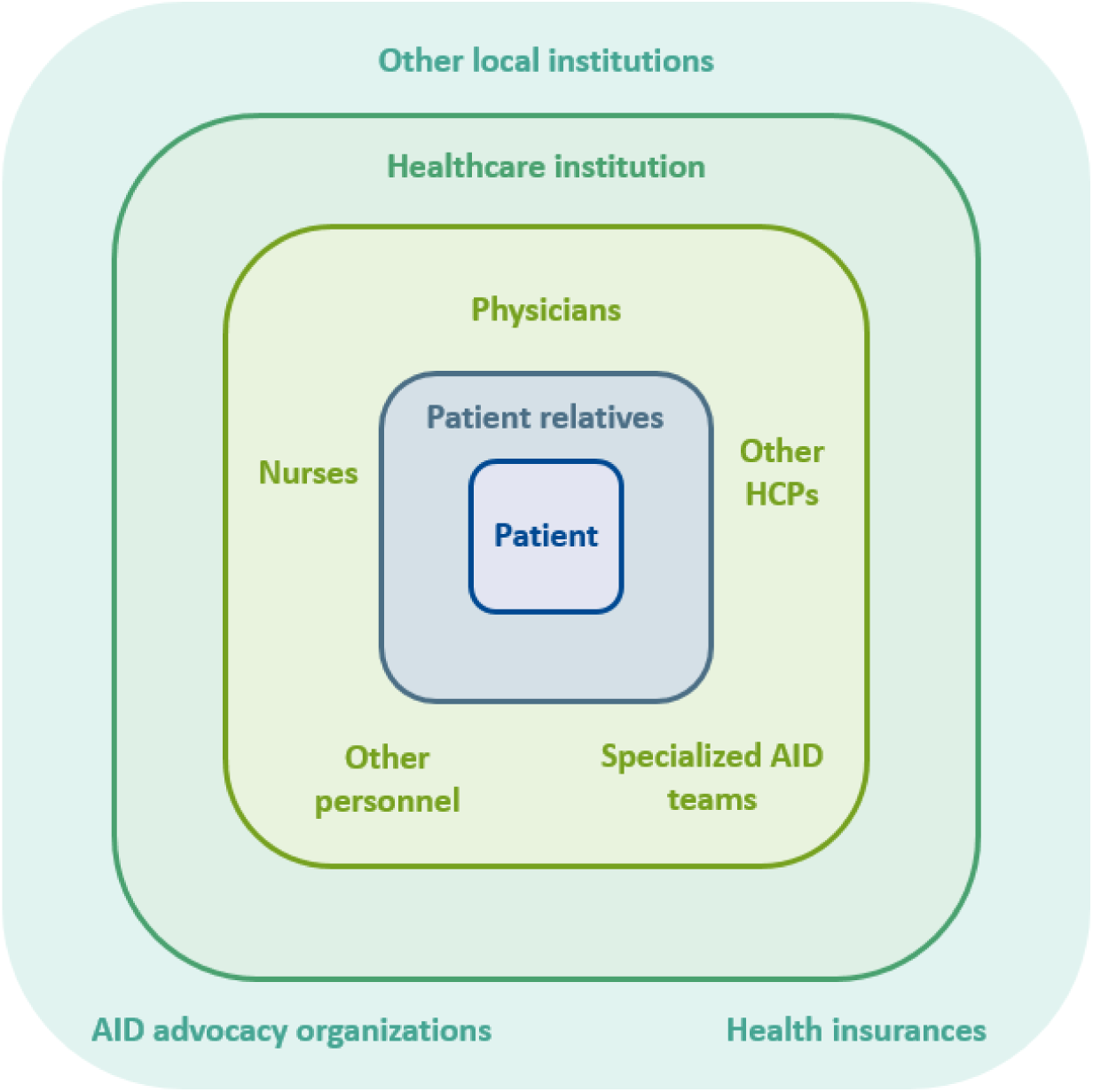
Participants of interactional AID decision-making processes in the healthcare context. Examples of “other HCPs” are social work, spiritual care, psychology, occupational and physical therapy, speech therapy, genetic counseling, pharmacy; examples of “other personnel” are ethicists, lawyers; examples of “other local institutions” are nursing homes.

As we focused our review on interactional decision-making processes in the healthcare context, we will use patients seeking AID as the point of reference and describe other participants’ involvement in the following. In the private sphere, patient relatives, especially family members, were described as participants. The nature of their involvement can vary. They can be crucial partners and support in the AID decision-making process, but there is also potential for conflict. It was also emphasized that patient relatives need support themselves, as the AID process can be very challenging for them. In some records, it was mentioned that patients and family members might not view healthcare as the right context for AID decision-making processes, but would rather locate them in the private sphere.

Regarding the professional healthcare context, physicians from different specialties were mentioned as participants in AID decision-making processes. This included general medicine, palliative care, psychiatry, oncology, geriatrics, and the specialty of the patient’s underlying disease. Often, more than one physician was described to be involved in the AID process for one patient. Sometimes, they had different roles, such as being the treating physician for the underlying disease, being the assessor, and being the physician to assist in dying. Some of the physicians involved were reported to have received specialized AID training. In some records, hesitancy of patients to involve physicians regarding AID was mentioned.

Nurses, other HCPs (e.g., social work, spiritual care, psychology, occupational and physical therapy, speech therapy, genetic counseling, pharmacy), other professionals (e.g., ethicists, lawyers), and volunteers were described as also being involved in AID decision-making processes in healthcare. Furthermore, specialized teams such as palliative care services or venous access support teams can have an important role in AID processes, as they have relevant specialized knowledge and skills. Others than physicians (i.e., other HCPs or volunteers) might be the first to be approached by a person seeking AID, sometimes unexpectedly, because they can have an especially trustful relationship with the patient. The involvement of other HCPs than physicians was described as currently under-recognized and under-used. In a multidisciplinary team reacting to AID requests, roles and responsibilities would need further clarification, good coordination and information exchange between HCPs would be crucial, and conscientious objections would need to be respected. Openness and support within teams was emphasized as paramount.

Some records described specialized AID teams. These could be a hospital’s internal AID resource group or committee; specified physician AID providers; AID navigators, coordinators, advocates, counsellors, pharmacists, or volunteers; or a centralized AID access point. When records mentioned such, they again emphasized the need for team-based approaches. Some records also mentioned the need to have different people responsible for regular clinical care, assessment of AID eligibility, and AID provision (e.g., Li et al. (2017)).

Beyond the individual and interactional level, how the healthcare institution handles AID was found to influence AID decision-making processes. For example, if an institution prohibits their employees to engage in AID processes or, on the contrary, gives them guidance and security when doing so. AID advocacy organizations can play an important role, especially in some countries such as Switzerland, where they coordinate much of the AID process. Furthermore, the involvement or disengagement in AID of other local institutions (e.g., nursing homes) and healthcare insurances (i.e., do they pay for AID) may make them participants in AID decision-making processes.

### AID as a longitudinal, iterative process (RQ2)

#### Phases of interactional AID decision-making processes

Many records described several phases or steps within interactional AID decision-making processes in healthcare (see Figure 3). We organized these phases into beginning, assessment, preparation, realization, and aftercare. In addition, we found the following transcendent themes described: coordination, person-centered care, and inter-/multidisciplinary teamwork. In the records, the mention of phases ranged from simple descriptions to complex flow-charts and frameworks (e.g., Crumley et al. (2021); Oczkowski et al. (2021b)). The phases were sometimes described as consecutive, sometimes as overlapping. The process can (and should) include loops and re-evaluations within and across the phases. The different phases will be described in the following.

**Figure 3.**
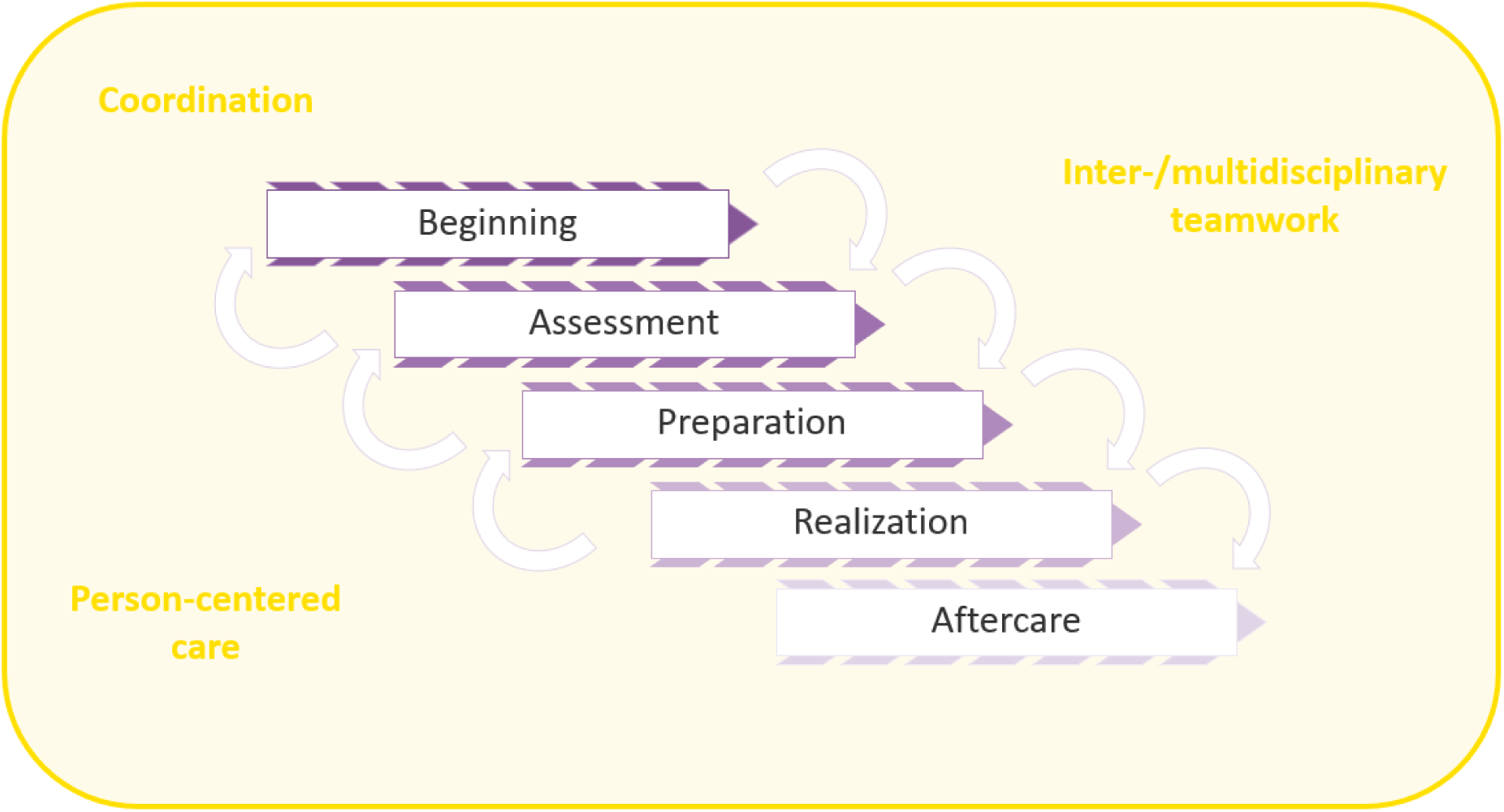
Phases of interactional AID decision-making processes in the healthcare context.

In the beginning phase, we found that AID is almost exclusively brought up by patients (or their family members), not by HCPs. It is thus important for HCPs to be prepared for and recognize AID requests voiced by patients. It was also suggested for HCPs to address these AID requests explicitly and openly, to listen, and to try to understand the patient’s perspective. The beginning phase included aspects such as relationship building, value clarification, contemplation, formation of intent, bringing up AID, recognizing AID requests, open discussion of requests, understanding underlying causes, talking about alternatives, possibly involving others such as patient relatives or AID specialists, and in some contexts a formal (written) AID request by the patient.

The assessment phase is the formal step of evaluating eligibility. Included records described that eligibly criteria for AID need to be assessed and due diligence needs to be ensured in this phase. This may include the involvement of more than one (independent) physician or HCP who assess the respective eligibility criteria.

The so-called preparation phase encompasses the concrete planning of the death. It was described to involve emotional processing as well as technical preparation. Anticipatory guidance on the physical process of dying and possible complications should be given.

The actual realization of AID was not the focus of this review, as most of the interactional decision-making process is concluded by then. However, it is important to mention that patients should have opportunities to re-evaluate and possibly change their mind about their decision up until the last moment. After a patient died with AID, aftercare was reported as an additional phase. Aftercare includes emotional processing as well as administrative duties and concerns for both HCPs and patient relatives.

#### Decisions made during AID processes

Different participants were found to make multiple decisions throughout the AID process. We attempted to structure them into different themes in the following. Patients form the intent to seek AID and decide to voice it. Predominantly HCPs make medical and psychosocial decisions (e.g., regarding prognostication, medical indication for AID, provision of a prescription, decision-making capacity and voluntariness, psychiatric disorders, coercion). Informational decisions involve, for example, the question, if the patient received sufficient information about their medical decision and alternatives to AID. Existential decisions such as the point at which suffering becomes unbearable to someone are rather patient-led. There are also organizational decisions (e.g., where, when, and with whom AID is planned to be carried out), safety-related decisions (e.g., who is responsible and/or reachable in case of adverse events during AID), and decisions if, when and how patient relatives are to be involved.

### Characteristics of communication during AID decision-making processes (RQ2)

AID decision-making has been described as a difficult process with high intensity for all participants, including HCPs. At the same time, the records mentioned potential for growth when participating in AID decision-making processes. A shared reflection process can help to go from vague requests to informed decisions. There is a need to meet both patient-centered care needs and procedural and legal requirements. To do so, it was highlighted that HCPs should avoid focusing predominantly on legal and biomedical aspects and need adequate personal, interpersonal, and structural resources. More detail on the content of communication can be found in Additional file 6.

When looking at influencing factors on interactional AID decision-making processes described in our data set, we found several facilitators and challenges. Selected facilitators were: 1) A good and trusting preexisting relationship between patient and HCP; 2) an open, curious, caring, non-judgmental attitude of HCPs; 3) guidelines on AID processes and communication; 4) confidential space and adequate time for communicating about AID; 5) maintaining appropriate boundaries as an HCP; 6) acknowledging the need for self-care for HCPs; and 7) having prior experience with AID requests and thus learning curves and growth processes. Selected challenges were: 1) AID requests challenging the therapeutic relationship and medical culture; 2) power imbalances between patients and HCPs; 3) avoiding HCPs’ subjective interpretations of AID requests; and 4) the need for self-reflection and self-awareness for HCPs.

### Identified research gaps and implications (RQ3)

When identifying research gaps, we primarily looked at interactional AID decision-making processes in particular, but also included research needs regarding AID processes in general. Overall, the records called for a) research to enhance the understanding of and b) research to improve AID processes.

Regarding methodology, 1) qualitative research (e.g., interviews, longitudinal participant observation, analyzing audio recordings of AID communication), 2) quantitative research based on previous qualitative results, with larger samples, and nationwide or international, 3) implementation research (e.g., for guidelines), and 4) participatory research were identified. Regarding research participants, the records called for the perspectives of patients, patient relatives, various specific groups of HCPs (e.g., nurses, psychologists, genetic counselors, volunteers), and research including different stakeholder perspectives.

Research gaps regarding the understanding of AID processes where reported for the development of theoretical frameworks, patient-centered care in AID (e.g., adapting the concept to AID) and the complex triadic decision-making between patients, their relatives, and HCPs. It should also become clearer, when to involve which specialty (e.g., psychiatry) and how psychological approaches can be used for AID. The content of AID communication, what information is given, and challenges in early communication about AID (analogously to advance care planning in palliative care) should be further explored. What happens after a patient is denied AID is currently insufficiently understood and studies on bereavement support are lacking. It should be investigated scientifically how to go about people seeking AID without a terminal illness and whether specific (vulnerable) populations should be eligible for AID (e.g., people with dementia, incarcerated people, minors). Access, affordability, and social disparities in the context of AID should be better understood. HCP-related factors such as gender, attitudes towards AID, and cognitive states (e.g., procedural action-focused or existential interpretative (Denier et al., 2009)) and their respective effects on AID decision-making processes need further research. Pressure experienced by HCPs and the involvement of HCPs with conscientious objections need clarification. Furthermore, it was suggested to observe AID implementation when AID becomes legal (including the impact on views of HCPs) and assess changes over time (i.e., after years of experience with AID in a country). Records called for international comparative studies (including those comparing countries where AID is legal and illegal), comparisons of different AID process models, and between different settings (including home care and elder care).

Research to improve AID processes should include the development and testing of AID-related questionnaires and screening tools for patients. AID educational tools and training for HCPs should be developed and evaluated. In addition, best practice for disseminating knowledge and reducing stigma is needed.

As for implications for practice, the records in this review called for thorough (institutional) policies and guidelines, clear standards for medical and psychiatric evaluation, better communication skills of HCPs and the need to enable them to sufficiently self-reflect, the creation of AID networks, and the establishment of registries.

### Expert workshop review

During the expert workshop, participants valued the work done, voiced that they were not surprised by the preliminary results, and did not question or criticize any findings. During discussion, they 1) highlighted the importance of team aspects and asked questions about 2) whether the records also reported on organizational level factors and 3) implementation processes of AID, 4) about the influence of different legal regulations, and 5) about related concepts (i.e., voluntary stopping of eating and drinking, aftercare of AID).

## DISCUSSION

In this scoping review, we found a large number of records with diverse methodology and study populations describing aspects of interactional AID decision-making processes. Besides patients seeking AID, we identified their relatives, physicians, nurses, other HCPs, and other personnel (beyond HCPs) as potentially involved in these processes. Some participate in specialized AID capacities, others untrained or even unexpected. In addition, we found characteristics of the healthcare institution and system level aspects such as characteristics of other local institutions, AID advocacy organizations, and health insurers to have an influence. The process of AID decision-making in healthcare is longitudinal and iterative, spanning from beginning through assessment, preparation, and realization to aftercare. Different participants make a range of decision throughout the process – some jointly, some not. Coordination, inter- and multidisciplinary teamwork, and person-centered care were mentioned as paramount. Communication during AID decision-making processes has been described as intense, highly complex, and influenced by various factors, but with potential for growth. The records in this scoping review called for additional research to improve the understanding of AID processes and the processes themselves.

In this scoping review, we used a wide angle to look at interactional AID decision-making and communication processes across patient and HCP populations, settings, and institutions. This allowed us to extend prior review articles that used a more narrow focus to understand the phenomenon for a specific context or profession (e.g., DeZeeuw and Myers (2020); Marina et al. (2021a); McCormack and Fléchais (2012); Selby and Bean (2019)).

From a practice-oriented point of view, we identified various people, who patients might talk to about AID requests. This scoping review highlighted that every person in healthcare, if HCP or volunteer, should be prepared to respond to AID being brought up, as they might be the first to hear about it. What their respective reaction should be depends on their respective role and how they are willing to be involved. Nevertheless, being aware and having self-reflected on AID seems advisable.

Regarding the normative question who should be involved in AID decision-making processes, we identified several groups of potential participants besides physicians, for example nurses and other HCPs such as psychologists. They were currently described as involved to a lesser degree and with less clear roles than desired. Furthermore, some records mentioned that patients might not even view the healthcare setting as the appropriate context for AID decision-making.

Records highlighted the high complexity of AID decision-making processes in a healthcare context, with several phases and decisions. This calls for appropriate guidance and support for the individuals involved that does not yet seem to exist on a regular basis. Such guidance could for example be practice guidelines or training. Training should include education, self-reflection, and communication skill development, and could be more basic or more advanced depending on the respective role. Support tools should give as much clarity as possible to make individual growth through these processes more likely and avoid harm for both patients and others involved.

Besides individual and interactional aspects, we also identified the need to be aware of organizational and system level influences. We found that AID decision-making processes are shaped also by characteristics of the healthcare institution and the broader contextual factors such as other local institutions, AID advocacy organizations or health insurances. Further, their role and impact largely depends on the respective healthcare system and legislation regarding AID. Implementation research frameworks, such as the Consolidated Framework for Implementation Research (CFIR) (Damschroder et al., 2022), could be a useful tool to structure and further interpret these findings.

From a scientific perspective, our results offer a first attempt to structure AID decision-making processes in healthcare on a higher level of abstraction. Therewith, they could serve as a basis for a precise conceptualization of such processes. The development of a scientific model of AID processes could be grounded on a framework derived from the results of this study. Scientific models of complex realities can help to better understand and explain the world and learning from the use of scientific models in the natural sciences might be beneficial (Bokulich, 2011; Weisberg, 2013). In addition, Metro Mapping, a methodology used to describe care paths in oncology, could be used as an innovative approach to describe AID decision-making processes in healthcare (Stiggelbout et al., 2023).

### Strengths and limitations

The broad search and inclusion criteria of this scoping review are a major strength that enabled us to find a broad range of records on our phenomenon of interest. This allowed a comprehensive synthesis of international findings on AID communication and decision-making. Furthermore, the methodology of a scoping review gave us sufficient degrees of freedom to capture the complexity of AID decision-making processes in healthcare. However, our decision to include such a variegated set of records came with limitations. Heterogeneity in the included study designs and populations was a challenge during the synthesis of results. Great differences in legal situations and healthcare structures across countries further complicated the interpretation of results. Furthermore, we were not able to carry out an assessment of the scientific quality of the included records. Nevertheless, we deem our decision to prioritize breadth over specificity for this scoping review appropriate as it enabled us to gain a comprehensive understanding of the current state of knowledge regarding interactional AID decision-making processes in healthcare.

## Conclusion

The scoping review at hand is the first to synthesize a large number of records addressing interactional decision-making processes on AID in a healthcare context. These records described AID decision-making as longitudinal and iterative, highly complex, and challenging. Many different people can be involved with varying perspectives and degrees of specific training, making various decisions. How communication unfolds is crucial in these processes and influenced by different factors such as HCPs stance on AID, the HCP-patient-relationship, and appropriate guidelines. Included records also identified the need for further research to better understand and improve AID processes.

## Supporting information

Gugel, Henning, Hahlweg. Supplemental material

## Data Availability

All data used in this scoping review is either published within the included records or added to this publication as supplementary material.

## Acknowledgements

We thank the advisory board of this study and the participants of the expert workshop for their valuable questions, input, and discussions. We also thank Robert Reincke who contributed to this publication as a student intern. Pola Hahlweg furthermore thanks Isabelle Scholl and Martin Härter for their supervision during conceptualization and funding acquisition. This work was supported by the Research Promotion Fund of the Medical Faculty of the University Hamburg (grant number NWF-22/02). The sponsor did not have a role in study design; data collection, analysis, and interpretation; writing of the article; or the decision to submit it for publication.

## GLOSSARY

AID: Aid in dying
AS: Assisted suicide
CG: Claudia Gugel
HCP: Health care professional
PH: Pola Hahlweg
PRISMA-ScR: Preferred Reporting Items for Systematic reviews and Meta-Analyses extension for Scoping Reviews
RQ: Research question
VE: Voluntary euthanasia
ZH: Zoe Henning

## DECLARATIONS

### Author contributions

CG contributed to formal analysis, investigation, methodology, project administration, and writing by reviewing and editing. ZH contributed to formal analysis, investigation, methodology, and writing by reviewing and editing. PH contributed to conceptualization, formal analysis, funding acquisition, investigation, methodology, project administration, supervision, visualization, and writing the original draft.

### Declaration of generative AI and AI-assisted technologies in the writing process

During the preparation of this work the authors used DeepL in order to improve language and readability. After using this tool, the authors reviewed and edited the content as needed and take full responsibility for the content of the publication.

## REFERNCES

Alvarado, V., & Liebig, B. (2015). “I want to die” - dealing with suicidal wishes in primary palliative care. Therapeutische Umschau, 72, 643–648.

Antifaeff, K. (2019). Social work practice with medical assistance in dying: A case study. Health & Social Work, 44, 185–192.

Arksey, H., & O’Malley, L. (2005). Scoping studies: towards a methodological framework. International Journal of Social Research Methodology, 8, 19–32.

Back, A.L., Starks, H., Hsu, C., Gordon, J.R., Bharucha, A., & Pearlman, R.A. (2002). Clinician-patient interactions about requests for physician-assisted suicide: a patient and family view. Arch Intern Med, 162, 1257–1265.

Balint, J.A. (2000). Decisions at the end of life. Croatian Medical Journal, 41, 144–149.

Ball, I.M., Hodge, B., Jansen, S., Nickle, S., & Sibbald, R.W. (2019). A Canadian academic hospital’s initial MAID experience: A health-care systems review. Journal of Palliative Care, 34, 78–84.

Bannink, M., Van Gool, A.R., van der Heide, A., & van der Maas, P.J. (2000). Psychiatric consultation and qualiity of decision making in euthanasia. Lancet, 356, 2067–2068.

Bokulich, A. (2011). How scientific models can explain. Synthese, 180, 33–45.

Booij, S.J., Rödig, V., Engberts, D.P., Tibben, A., & Roos, R.A. (2013). Euthanasia and advance directives in Huntington’s disease: qualitative analysis of interviews with patients. J Huntingtons Dis, 2, 323–330.

Borgsteede, S.D., Deliens, L., Graafland-Riedstra, C., Francke, A.L., van der Wal, G., & Willems, D.L. (2007). Communication about euthanasia in general practice: Opinions and experiences of patients and their general practitioners. Patient Education and Counseling, 66, 156–161.

Braddock, C.H., Fihn, S.D., Levinson, W., Jonsen, A.R., & Pearlman, R.A. (1997). How doctors and patients discuss routine clinical decisions: Informed decision making in the outpatient setting. Journal of General Internal Medicine, 12, 339–345.

Brooks, L. (2019). Health care provider experiences of and perspectives on medical assistance in dying: a scoping review of qualitative studies. Canadian Journal on Aging / La Revue canadienne du vieillissement, 38, 384–396.

Brown, J., Goodridge, D., Harrison, A., Kemp, J., Thorpe, L., & Weiler, R. (2020a). Care considerations in a patient- and family-centered Medical Assistance in Dying program. Journal of Palliative Care, 11.

Brown, J., Goodridge, D., Harrison, A., Kemp, J., Thorpe, L., & Weiler, R. (2020b). Medical assistance in dying: Patients’, families’, and health care providers’ perspectives on access and care delivery. Journal of Palliative Medicine, 23, 1468–1477.

Buchbinder, M., Brassfield, E.R., & Mishra, M. (2019). Health care providers’ experiences with implementing Medical Aid-in-Dying in Vermont: a qualitative study. Journal of General Internal Medicine, 34, 636–641.

Buchbinder, M., Ojo, E., Knio, L., & Brassfield, E.R. (2018). Caregivers’ experiences with Medical Aid-In-Dying in Vermont: A qualitative study. Journal of Pain and Symptom Management, 56, 936–943.

Carlson, B., Simopolous, N., Goy, E.R., Jackson, A., & Ganzini, L. (2005). Oregon hospice chaplains’ experiences with patients requesting physician-assisted suicide. Journal of Palliative Medicine, 8, 1160–1166.

Chambaere, K., Bilsen, J., Cohen, J., Onwuteaka-Philipsen, B.D., Mortier, F., & Deliens, L. (2011). Trends in Medical End-of-Life decision making in Flanders, Belgium 1998-2001-2007. Medical Decision Making, 31, 500–510.

Chambaere, K., Cohen, J., Robijn, L., Bailey, S.K., & Deliens, L. (2015). End-of-life decisions in individuals dying with dementia in Belgium. Journal of the American Geriatrics Society, 63, 290–296.

Claxton-Oldfield, S., & Beaudette, S. (2021). Hospice palliative care volunteers’ attitudes, opinions, experiences, and perceived needs for training around medical assistance in dying (MAiD). Am J Hosp Palliat Care, 38, 1282–1290.

Cohen, J., Bilsen, J., Fischer, S., Loefmark, R., Norup, M., van der Heide, A., et al. (2007). End-of-life decision-making in Belgium, Denmark, Sweden and Switzerland: Does place of death make a difference? Journal of Epidemiology and Community Health, 61, 1062–1068.

Cohen, J., Van Wesemael, Y., Smets, T., Bilsen, J., Onwuteaka-Philipsen, B., Distelmans, W., et al. (2014). Nationwide survey to evaluate the decision-making process in euthanasia requests in Belgium: do specifically trained 2nd physicians improve quality of consultation? Bmc Health Services Research, 14, 9.

Cole, R.M. (1993). Communicating with people who request euthanasia. Palliat Med, 7, 139–143.

Colquhoun, H.L., Levac, D., O’Brien, K.K., Straus, S., Tricco, A.C., Perrier, L., et al. (2014). Scoping reviews: time for clarity in definition, methods, and reporting. J Clin Epidemiol, 67, 1291–1294.

Coscarelli, A., Bonet, L., Cleary, E., Eselun, M., Johnson, A., Kim, S.J., et al. (2017). Does psychosocial oncology have a greater role in aid-in-dying prescriptions than assessing decisional capacity? Psycho-Oncology, 26, 38–39.

Crumley, E.T., Kelly, S., Young, J., Phinney, N., McCarthy, J., & Gubitz, G. (2021). How is the medical assistance in dying (MAID) process carried out in Nova Scotia, Canada? A qualitative process model flowchart study. BMJ Open, 11, e048698.

D’Angelo, A., Ormond, K.E., Magnus, D., & Tabor, H.K. (2019). Assessing genetic counselors’ experiences with physician aid-in-dying and practice implications. Journal of Genetic Counseling, 28, 164–173.

Damron, L., Bayram, E., McGehrin, K., Reynolds, J., Hess, R., Coughlin, D.G., et al. (2022). Physician-assisted dying: Access and utilization in patients with movement disorders. Movement Disorders, 6.

Damschroder, L.J., Reardon, C.M., Widerquist, M.A.O., & Lowery, J. (2022). The updated Consolidated Framework for Implementation Research based on user feedback. Implementation Science, 17, 75.

Davila, S.L.R., Vidal, E., Stewart, J.T., & Caserta, M.T. (2010). Management of a request for physician-assisted suicide. American Journal of Hospice & Palliative Medicine, 27, 63–65.

de Bal, N., de Casterlé, B.D., de Beer, T., & Gastmans, C. (2006). Involvement of nurses in caring for patients requesting euthanasia in Flanders (Belgium): A qualitative study. International Journal of Nursing Studies, 43, 589–599.

de Boer, M.E., Depla, M., den Breejen, M., Slottje, P., Onwuteaka-Philipsen, B.D., & Hertogh, C. (2019). Pressure in dealing with requests for euthanasia or assisted suicide. Experiences of general practitioners. Journal of Medical Ethics, 45, 425–429.

de Boer, M.E., Droees, R.M., Jonker, C., Eefsting, J.A., & Hertogh, C. (2011). Advance directives for euthanasia in dementia: How do they affect resident care in Dutch nursing homes? Experiences of physicians and relatives. Journal of the American Geriatrics Society, 59, 989–996.

de Casterlé, B.D., Denier, Y., de Bal, N., & Gastmans, C. (2010). Nursing care for patients requesting euthanasia in general hospitals in Flanders, Belgium. *Journal of Advanced Nursing (John Wiley & Sons*, Inc.), 66, 2410–2420.

de Casterlé, B.D., Verpoort, C., de Bal, N., & Gastmans, C. (2006). Nurses’ views on their involvement in euthanasia: a qualitative study in Flanders (Belgium). Journal of Medical Ethics, 32, 187–192.

de Veer, A.J., Francke, A.L., & Poortvliet, E.P. (2008). Nurses’ involvement in end-of-life decisions. Cancer Nurs, 31, 222–228.

Dees, M.K., Vernooij-Dassen, M.J., Dekkers, W.J., Elwyn, G., Vissers, K.C., & van Weel, C. (2013). Perspectives of decision-making in requests for euthanasia: A qualitative research among patients, relatives and treating physicians in the Netherlands. Palliative Medicine, 27, 27–37.

Denier, Y., de Casterle, B.D., De Bal, N., & Gastmans, C. (2009). Involvement of nurses in the euthanasia care process in Flanders (Belgium): An exploration of two perspectives. Journal of Palliative Care, 25, 264–274.

Denier, Y., Dierckx de Casterlé, B., De Bal, N., & Gastmans, C. (2010). “It’s intense, you know.” Nurses’ experiences in caring for patients requesting euthanasia. Med Health Care Philos, 13, 41–48.

Deschepper, R., Vander Stichele, R., Bernheim, J.L., De Keyser, E., Van der Kelen, G., Mortier, F., et al. (2006). Communication on end-of-life decisions with patients wishing to die at home: the making of a guideline for GPs in Flanders, Belgium. British Journal of General Practice, 56, 14–19.

DeZeeuw, K., & Myers, E.L. (2020). The role of speech-language pathologists in medical assistance in dying: Canadian experience to inform clinical practice. Canadian Journal of Speech-Language Pathology and Audiology, 44, 49–56.

Dobscha, S.K., Heintz, R.T., Press, N., & Ganzini, L. (2004). Oregon physicians’ responses to requests for assisted suicide: a qualitative study. J Palliat Med, 7, 451–461.

Driftmier, P., & Shaw, J. (2021). Medical Assistance in Dying (MAiD) for Canadian prisoners: A case series of barriers to care in completed MAiD deaths. Health Equity, 5, 847–853.

Elwyn, G., Durand, M.A., Song, J., Aarts, J., Barr, P.J., Berger, Z., et al. (2017). A three-talk model for shared decision making: multistage consultation process. BMJ, 359, j4891.

Emanuel, E.J., Daniels, E.R., Fairclough, D.L., & Clarridge, B.R. (1998). The practice of euthanasia and physician-assisted suicide in the United States - Adherence to proposed safeguards and effects on physicians. Jama-Journal of the American Medical Association, 280, 507–513.

Emanuel, E.J., Onwuteaka-Philipsen, B.D., Urwin, J.W., & Cohen, J. (2016). Attitudes and practices of euthanasia and physician-assisted suicide in the United States, Canada, and Europe. Jama, 316, 79–90.

Evenblij, K., Pasman, H.R.W., Pronk, R., & Onwuteaka-Philipsen, B.D. (2019). Euthanasia and physician-assisted suicide in patients suffering from psychiatric disorders: a cross-sectional study exploring the experiences of Dutch psychiatrists. Bmc Psychiatry, 19, 10.

Gamondi, C., Borasio, G.D., Oliver, P., Preston, N., & Payne, S. (2019). Responses to assisted suicide requests: an interview study with Swiss palliative care physicians. Bmj Supportive & Palliative Care, 9, 9.

Gamondi, C., Pott, M., Forbes, K., & Payne, S. (2015). Exploring the experiences of bereaved families involved in assisted suicide in Southern Switzerland: a qualitative study. Bmj Supportive & Palliative Care, 5, 146–152.

Gamondi, C., Pott, M., & Payne, S. (2013). Families’ experiences with patients who died after assisted suicide: a retrospective interview study in southern Switzerland. Annals of Oncology, 24, 1639–1644.

Gamondi, C., Pott, M., Preston, N., & Payne, S. (2018). Family caregivers’ reflections on experiences of assisted suicide in Switzerland: A qualitative interview study. Journal of Pain and Symptom Management, 55, 1085–1094.

Gamondi, C., Pott, M., Preston, N., & Payne, S. (2020). Swiss families’ experiences of interactions with providers during assisted suicide: A secondary data analysis of an interview study. Journal of Palliative Medicine, 23, 506–512.

Gastmans, C., Van Neste, F., & Schotsmans, P. (2004). Facing requests for euthanasia: a clinical practice guideline. Journal of Medical Ethics, 30, 212–217.

Goh, E., Wen, J., & Yu, C.E. (2021). Lessons from the departed: A planned behavior approach to understand travelers’ actual physician-assisted suicide behavior. Journal of Hospitality & Tourism Research, 15.

Groenewoud, J.H., van der Heide, A., Tholen, A.J., Schudel, W.J., Hengeveld, M.W., Onwuteaka-Philipsen, B.D., et al. (2004). Psychiatric consultation with regard to requests for euthanasia or physician-assisted suicide. General Hospital Psychiatry, 26, 323–330.

Hagens, M., Snijdewind, M.C., Evenblij, K., Onwuteaka-Philipsen, B.D., & Pasman, H.R.W. (2021). Experiences with counselling to people who wish to be able to self-determine the timing and manner of one’s own end of life: a qualitative in-depth interview study. Journal of Medical Ethics, 47, 39–46.

Hamilton, N.G., & Hamilton, C.A. (2005). Competing paradigms of response to assisted suicide requests in Oregon. Am J Psychiatry, 162, 1060–1065.

Handley, M.A. (2020). The “Foresty Way”: My mother’s brave choice of Medical Aid in Dying. Annals of Family Medicine, 18, 553–554.

Health Canada. (2019). First annual report on medical assistance in dying in Canada.

Health Canada. (2020). Second annual report on medical assistance in dying in Canada.

Health Canada. (2021). Third annual report on medical assistance in dying in Canada.

Ho, A., Norman, J.S., Joolaee, S., Serota, K., Twells, L., & William, L. (2021). How does Medical Assistance in Dying affect end-of-life care planning discussions? Experiences of Canadian multidisciplinary palliative care providers. Palliative Care & Social Practice, 1–14.

Hopkins, H.J., & Hopkins, K.D. (2017). When choosing death is an affirmation of life: A public remembrance of our family’s journey. Review & Expositor, 114, 414–423.

Inghelbrecht, E., Bilsen, J., Mortier, F., & Deliens, L. (2010). The role of nurses in physician-assisted deaths in Belgium. Canadian Medical Association Journal, 182, 905–910.

Jansen-van der Weide, M.C., Onwuteaka-Philipsen, B.D., Heide, A., & Wal, G. (2009). How patients and relatives experience a visit from a consulting physician in the euthanasia procedure: a study among relatives and physicians. Death Stud, 33, 199–219.

Jansen-van der Weide, M.C., Onwuteaka-Philipsen, B.D., & van der Wal, G. (2005). Granted, undecided, withdrawn, and refused requests for euthanasia and physician-assisted suicide. Arch Intern Med, 165, 1698–1704.

Jeba, J. (2019). Did I hear you right? Journal of Pain & Palliative Care Pharmacotherapy, 33, 59–61.

Kompanje, E.J.O., de Beaufort, I.D., & Bakker, J. (2007). Euthanasia in intensive care: A 56-year-old man with a pontine hemorrhage resulting in a locked-in syndrome. Critical Care Medicine, 35, 2428–2430.

Kouwenhoven, P.S.C., Raijmakers, N.J.H., van Delden, J.J.M., Rietjens, J.A.C., van Tol, D.G., van de Vathorst, S., et al. (2015). Opinions about euthanasia and advanced dementia: a qualitative study among Dutch physicians and members of the general public. Bmc Medical Ethics, 16, 6.

Lehto, R.H., Olsen, D.P., & Chan, R.R. (2016). When a patient discusses assisted dying: Nursing practice implications. Journal of Hospice & Palliative Nursing, 18, 184–191.

Lemiengre, J., de Casterle, B.D., Denier, Y., Schotsmans, P., & Gastmans, C. (2008). How do hospitals deal with euthanasia requests in Flanders (Belgium)? A content analysis of policy documents. Patient Education and Counseling, 71, 293–301.

Li, M., & Kain, D. (2018). The other side of sorrow: physician reflections on assisted dying. Cmaj, 190, E169–e170.

Li, M., Watt, S., Escaf, M., Gardam, M., Heesters, A., O’Leary, G., et al. (2017). Medical assistance in dying - Implementing a hospital-based program in Canada. New England Journal of Medicine, 376, 2082–2088.

Loggers, E.T., Starks, H., Shannon-Dudley, M., Back, A.L., Appelbaum, F.R., & Stewart, F.M. (2013). Implementing a Death with Dignity program at a comprehensive cancer center. New England Journal of Medicine, 368, 1417–1424.

Marina, S., Wainwright, T., & Ricou, M. (2021a). The role of psychologists in requests to hasten death: A literature and legislation review and an agenda for future research. International Journal of Psychology, 56, 64–74.

Marina, S., Wainwright, T., & Ricou, M. (2021b). Views of psychologists about their role in hastened death. Omega-Journal of Death and Dying, 16.

Matzo, M.L., & Emanual, E.J. (1997). Oncology nurses’ practices of assisted suicide and patient-requested euthanasia. Oncol Nurs Forum, 24, 1725–1732.

McCormack, R., & Fléchais, R. (2012). The role of psychiatrists and mental disorder in assisted dying practices around the world: a review of the legislation and official reports. Psychosomatics, 53, 319–326.

Miller, D.G., Dresser, R., & Kim, S.Y.H. (2019). Advance euthanasia directives: a controversial case and its ethical implications. Journal of Medical Ethics, 45, 84–89.

Mroz, S., Dierickx, S., Deliens, L., Cohen, J., & Chambaere, K. (2021). Assisted dying around the world: a status quaestionis. Ann Palliat Med, 10, 3540–3553.

National Academies of Sciences Engineering and Medicine. (2018). Physician-assisted death: Scanning the landscape: Proceedings of a workshop. Washington DC, USA.

Oczkowski, S.J.W., Crawshaw, D., Austin, P., Versluis, D., Kalles-Chan, G., Kekewich, M., et al. (2021a). How we can improve the quality of care for patients requesting medical assistance in dying: A qualitative study of health care providers. J Pain Symptom Manage, 61, 513–521.e518.

Oczkowski, S.J.W., Crawshaw, D.E., Austin, P., Versluis, D., Kalles-Chan, G., Kekewich, M., et al. (2021b). How can we improve the experiences of patients and families who request medical assistance in dying? A multi-centre qualitative study. BMC Palliative Care, 20, 1–12.

Ofstad, E.H., Frich, J.C., Schei, E., Frankel, R.M., & Gulbrandsen, P. (2016). What is a medical decision? A taxonomy based on physician statements in hospital encounters: a qualitative study. BMJ Open, 6, e010098.

Onwuteaka-Philipsen, B.D., & van der Wal, G. (2001). A protocol for consultation of another physician in cases of euthanasia and assisted suicide. Journal of Medical Ethics, 27, 331–337.

Onwuteaka-Philipsen, B.D., van der Wal, G., Kostense, P.J., & van der Maas, P.J. (2000). Consultation with another physician on euthanasia and assisted suicide in the Netherlands. Social Science & Medicine, 51, 429–438.

Pasman, H.R., Willems, D.L., & Onwuteaka-Philipsen, B.D. (2013). What happens after a request for euthanasia is refused? Qualitative interviews with patients, relatives and physicians. Patient Educ Couns, 92, 313–318.

Patel, T., Christy, K., Grierson, L., Shadd, J., Farag, A., O’Toole, D., et al. (2021). Clinician responses to legal requests for hastened death: a systematic review and meta-synthesis of qualitative research. BMJ Supportive & Palliative Care, 11, 59–67.

Pesut, B., Thorne, S., Schiller, C., Greig, M., Roussel, J., & Tishelman, C. (2020a). Constructing good nursing practice for medical assistance in dying in Canada: An interpretive descriptive study. Global Qualitative Nursing Research, 7, 11.

Pesut, B., Thorne, S., Schiller, C.J., Greig, M., & Roussel, J. (2020b). The rocks and hard places of MAiD: a qualitative study of nursing practice in the context of legislated assisted death. BMC Nursing, 19, 1–14.

Regional euthanasia review committees. (2002). Annual report 2002.

Regional euthanasia review committees. (2003). Annual report 2003.

Regional euthanasia review committees. (2004). Annual report 2004.

Regional euthanasia review committees. (2005). Annual report 2005.

Regional euthanasia review committees. (2006). Annual report 2006.

Regional euthanasia review committees. (2007). Annual report 2007.

Regional euthanasia review committees. (2008). Annual report 2008.

Regional euthanasia review committees. (2009). Annual report 2009.

Regional euthanasia review committees. (2010). Annual report 2010.

Regional euthanasia review committees. (2011). Annual report 2011.

Regional euthanasia review committees. (2012). Annual report 2012.

Regional euthanasia review committees. (2013). Annual report 2013.

Regional euthanasia review committees. (2014). Annual report 2014.

Regional euthanasia review committees. (2015). Annual report 2015.

Regional euthanasia review committees. (2016). Annual report 2016.

Regional euthanasia review committees. (2017). Annual report 2017.

Regional euthanasia review committees. (2018). Annual report 2018.

Regional euthanasia review committees. (2019). Annual report 2019.

Richards, N. (2017). Assisted suicide as a remedy for suffering? The end-of-life preferences of British “suicide tourists”. Medical Anthropology, 36, 348–362.

Rodriguez Davila, S.L., Vidal, E., Stewart, J.T., & Caserta, M.T. (2010). Management of a request for physician-assisted suicide. Am J Hosp Palliat Care, 27, 63–65.

Schmahl, O., Oude Voshaar, R., van de Poel-Mustafayeva, A., & Marijnissen, R. (2021). Request for euthanasia by a psychiatric patient with undetected intellectual disability. BMJ Case Rep, 14.

Schmid, M., Zellweger, U., Bosshard, G., Bopp, M., & Swiss Med End-Of-Life, D. (2016). Medical end-of-life decisions in Switzerland 2001 and 2013: Who is involved and how does the decision-making capacity of the patient impact? Swiss Medical Weekly, 146, 10.

Schuurmans, J., Vos, S., Vissers, P., Tilburgs, B., & Engels, Y. (2020). Supporting GPs around euthanasia requests from people with dementia: a qualitative analysis of Dutch nominal group meetings. British Journal of General Practice, 70, E833-E842.

Selby, D., & Bean, S. (2019). Oncologists communicating with patients about assisted dying. Curr Opin Support Palliat Care, 13, 59–63.

Sellars, M., White, B.P., Yates, P., & Willmott, L. (2022). Medical practitioners’ views and experiences of being involved in assisted dying in Victoria, Australia: A qualitative interview study among participating doctors. Social Science & Medicine, 292, N.PAG-N.PAG.

Shaw, J., Harper, L., Preston, E., Wright, A., Kelly, M., & Wiebe, E. (2021). Perceptions and experiences of medical assistance in dying among illicit substance users and people living in poverty. Omega: Journal of Death & Dying, 84, 267–288.

Snijdewind, M.C., van Tol, D.G., Onwuteaka-Philipsen, B.D., & Willems, D.L. (2014). Complexities in euthanasia or physician-assisted suicide as perceived by Dutch physicians and patients’ relatives. Journal of Pain and Symptom Management, 48, 1125–1134.

Snijdewind, M.C., van Tol, D.G., Onwuteaka-Philipsen, B.D., & Willems, D.L. (2018). Developments in the practice of physician-assisted dying: perceptions of physicians who had experience with complex cases. Journal of Medical Ethics, 44, 292–296.

Starks, H., Back, A.L., Pearlman, R.A., Koenig, B.A., Hsu, C., Gordon, J.R., et al. (2007). Family member involvement in hastened death. Death Studies, 31, 105–130.

Stewart, D.E., Rodin, G., & Li, M. (2018). Consultation-liaison psychiatry and physician-assisted death. General Hospital Psychiatry, 55, 15–19.

Stiggelbout, A., Griffioen, I., Brands, J., Melles, M., Rietjens, J., Kunneman, M., et al. (2023). Metro Mapping: development of an innovative methodology to co-design care paths to support shared decision making in oncology. BMJ Evid Based Med, 28, 291–294.

Stokes, F. (2017). The emerging role of nurse practitioners in physician-assisted death. The Journal for Nurse Practitioners, 13, 150–155.

ten Cate, K., van Tol, D.G., & van de Vathorst, S. (2017). Considerations on requests for euthanasia or assisted suicide; a qualitative study with Dutch general practitioners. Family Practice, 34, 723–729.

Tricco, A.C., Lillie, E., Zarin, W., O’Brien, K.K., Colquhoun, H., Levac, D., et al. (2018). PRISMA Extension for Scoping Reviews (PRISMA-ScR): Checklist and explanation. Annals of Internal Medicine, 169, 467–473.

van Bruchem-ven de Scheur, G.G., van der Arend, A.J.G., Abu-Saad, H.H., Spreeuwenberg, C., van Wijmen, F.C.B., & ter Meulen, R.H.J. (2008). The role of nurses in euthanasia and physician-assisted suicide in The Netherlands. Journal of Medical Ethics, 34, 254–258.

Van Wesemael, Y., Cohen, J., Bilsen, J., Onwuteaka-Philipsen, B.D., Distelmans, W., & Deliens, L. (2010). Consulting a trained physician when considering a request for euthanasia: An evaluation of the process in Flanders and the Netherlands. Evaluation & the Health Professions, 33, 497–513.

Verhofstadt, M., Van Assche, K., Sterckx, S., Audenaert, K., & Chambaere, K. (2019). Psychiatric patients requesting euthanasia: Guidelines for sound clinical and ethical decision making. International Journal of Law and Psychiatry, 64, 150–161.

Weisberg, M. (2013). Simulation and similarity: Using models to understand the world: Oxford University Press.

Wiebe, E., Kelly, M., McMorrow, T., Tremblay-Huet, S., & Hennawy, M. (2021). Assessment of capacity to give informed consent for medical assistance in dying: a qualitative study of clinicians’ experience. CMAJ Open, 9, E358–e363.

Zworth, M., Saleh, C., Ball, I., Kalles, G., Chkaroubo, A., Kekewich, M., et al. (2020). Provision of medical assistance in dying: a scoping review. BMJ Open, 10, e036054.

