## Supplementary material for "Interactional decision-making processes and communication on aid in dying: an international scoping review": Gugel, Henning, Hahlweg. Supplemental material

- **Additional file 1**: Preferred Reporting Items for Systematic reviews and Meta-Analyses extension for Scoping Reviews (PRISMA-ScR) Checklist
- **Additional file 2**: Literature search
- **Additional file 3**: Title & abstract screening
- **Additional file 4**: Full-text assessment
- **Additional file 5**: Data extraction process
- **Additional file 6**: Data extraction sheet

Gugel C, Henning Z, Hahlweg P. Interactional decision-making processes and communication on aid in dying: an international scoping review. **Additional file 1.**

**Preferred Reporting Items for Systematic reviews and Meta-Analyses extension for Scoping Reviews (PRISMA-ScR) Checklist**

| SECTION | ITEM | PRISMA-ScR CHECKLIST ITEM | REPORTED ON PAGE # |
|---|---|---|---|
| **TITLE** | | | |
| Title | 1 | Identify the report as a scoping review. | 1 |
| **ABSTRACT** | | | |
| Structured summary | 2 | Provide a structured summary that includes (as applicable): background, objectives, eligibility criteria, sources of evidence, charting methods, results, and conclusions that relate to the review questions and objectives. | 2 |
| **INTRODUCTION** | | | |
| Rationale | 3 | Describe the rationale for the review in the context of what is already known. Explain why the review questions/objectives lend themselves to a scoping review approach. | 4-5 |
| Objectives | 4 | Provide an explicit statement of the questions and objectives being addressed with reference to their key elements (e.g., population or participants, concepts, and context) or other relevant key elements used to conceptualize the review questions and/or objectives. | 5 |
| **METHODS** | | | |
| Protocol and registration | 5 | Indicate whether a review protocol exists; state if and where it can be accessed (e.g., a Web address); and if available, provide registration information, including the registration number. | 5 |
| Eligibility criteria | 6 | Specify characteristics of the sources of evidence used as eligibility criteria (e.g., years considered, language, and publication status), and provide a rationale. | 6 |
| Information sources* | 7 | Describe all information sources in the search (e.g., databases with dates of coverage and contact with authors to identify additional sources), as well as the date the most recent search was executed. | 7 |
| Search | 8 | Present the full electronic search strategy for at least 1 database, including any limits used, such that it could be repeated. | Add. File 2 |
| Selection of sources of evidence† | 9 | State the process for selecting sources of evidence (i.e., screening and eligibility) included in the scoping review. | 7-8, Add. Files 3 and 4 |
| Data charting process‡ | 10 | Describe the methods of charting data from the included sources of evidence (e.g., calibrated forms or forms that have been tested by the team before their use, and whether data charting was done independently or in duplicate) and any processes for obtaining and confirming data from investigators. | 8, Add. File 5 |
| Data items | 11 | List and define all variables for which data were sought and any assumptions and simplifications made. | 8, Add. File 5 |
| Critical appraisal of individual sources of evidence§ | 12 | If done, provide a rationale for conducting a critical appraisal of included sources of evidence; describe the methods used and how this information was used in any data synthesis (if appropriate). | n/a |

St. Michael's
Inspired Care.
Inspiring Science.

Gugel C, Henning Z, Hahlweg P. Interactional decision-making processes and communication on aid in dying: an international scoping review. **Additional file 1.**

| SECTION | ITEM | PRISMA-ScR CHECKLIST ITEM | REPORTED ON PAGE # |
|---|---|---|---|
| Synthesis of results | 13 | Describe the methods of handling and summarizing the data that were charted. | 8, Add. File 5 |
| **RESULTS** | | | |
| Selection of sources of evidence | 14 | Give numbers of sources of evidence screened, assessed for eligibility, and included in the review, with reasons for exclusions at each stage, ideally using a flow diagram. | 9, Add. Files 3 and 4 |
| Characteristics of sources of evidence | 15 | For each source of evidence, present characteristics for which data were charted and provide the citations. | 9-13, Add File 6 |
| Critical appraisal within sources of evidence | 16 | If done, present data on critical appraisal of included sources of evidence (see item 12). | n/a |
| Results of individual sources of evidence | 17 | For each included source of evidence, present the relevant data that were charted that relate to the review questions and objectives. | Add. File 6 |
| Synthesis of results | 18 | Summarize and/or present the charting results as they relate to the review questions and objectives. | 13-19 |
| **DISCUSSION** | | | |
| Summary of evidence | 19 | Summarize the main results (including an overview of concepts, themes, and types of evidence available), link to the review questions and objectives, and consider the relevance to key groups. | 20 |
| Limitations | 20 | Discuss the limitations of the scoping review process. | 22 |
| Conclusions | 21 | Provide a general interpretation of the results with respect to the review questions and objectives, as well as potential implications and/or next steps. | 23 |
| **FUNDING** | | | |
| Funding | 22 | Describe sources of funding for the included sources of evidence, as well as sources of funding for the scoping review. Describe the role of the funders of the scoping review. | 24 |

JBI = Joanna Briggs Institute; PRISMA-ScR = Preferred Reporting Items for Systematic reviews and Meta-Analyses extension for Scoping Reviews.

* Where *sources of evidence* (see second footnote) are compiled from, such as bibliographic databases, social media platforms, and Web sites.

† A more inclusive/heterogeneous term used to account for the different types of evidence or data sources (e.g., quantitative and/or qualitative research, expert opinion, and policy documents) that may be eligible in a scoping review as opposed to only studies. This is not to be confused with *information sources* (see first footnote).

‡ The frameworks by Arksey and O'Malley (6) and Levac and colleagues (7) and the JBI guidance (4, 5) refer to the process of data extraction in a scoping review as data charting.

§ The process of systematically examining research evidence to assess its validity, results, and relevance before using it to inform a decision. This term is used for items 12 and 19 instead of "risk of bias" (which is more applicable to systematic reviews of interventions) to include and acknowledge the various sources of evidence that may be used in a scoping review (e.g., quantitative and/or qualitative research, expert opinion, and policy document).

*From:* Tricco AC, Lillie E, Zarin W, O'Brien KK, Colquhoun H, Levac D, et al. PRISMA Extension for Scoping Reviews (PRISMAScR): Checklist and Explanation. Ann Intern Med. 2018;169:467–473. doi: 10.7326/M18-0850.

### Additional file 2 | Literature search

#### A Primary database search

**<u>Search strings</u>**

***Web of Science***

Filters:

- Date limit: none
- Language: English, German
- Web of Science Categories: ausgeschlossen sind alle Kategorien, die sich nicht auf Menschen beziehen, z.B. Tiermedizin oder Ingenieurswissenschaften o.ä.

*TS = ("assisted suicide" OR euthanasia OR "assis* dying" OR "aid in dying" OR "hasten* death" OR "termination of life" OR "assistiert* Suizid" OR "Tötung auf Verlangen" OR Sterbehilfe OR Suizidbeihilfe OR Suizidass* OR "assistiert* Selbsttötung" OR "hilfe zum Suizid" OR "hilfe zur Selbsttötung") AND ALL = (decision* OR interact* OR communic* OR counsel* OR choice* OR guid* OR support* OR interaktion* OR kommunikation* OR beratung* OR entscheidung* OR begleit* OR unterstütz*)*

***PubMed***

Filters:

- Date limit: none
- Language: English, German
- Species: Human

*((((((((((assisted suicide[Title/Abstract])) OR ("Euthanasia, Active"[Mesh]) OR (("Euthanasia, Active, Voluntary"[Mesh] ))) OR (assisted dying[Title/Abstract])) OR (assistance in dying[Title/Abstract])) OR (aid in dying[Title/Abstract])) OR (hasten* death[Title/Abstract])) OR (termination of life[Title/Abstract])) OR (assistierter Suizid[Title/Abstract])) OR (Sterbehilfe[Title/Abstract])) OR (Suizidbeihilfe[Title/Abstract])) AND (((((((((((((decision*) OR (("Decision Making"[Mesh]))) OR (communication)) OR (interact*)) OR (interakt*)) OR (("Counseling"[Mesh]))) OR (beratung*)) OR (counsel*)) OR (entscheidung*)) OR (choice)) OR (begleit*)) OR (unterstütz*)) OR (support*)) OR (guid*))*

***CINAHL***

Filters:

- Date limit: none
- Language: English, German
- Exclude medline records
- Species: Human

*(((MH "Suicide, Assisted") OR (MH "Euthanasia+") OR "assis* dying" OR "aid in dying" OR "hasten* death" OR "termination of life" OR "assistiert* suizid" OR "tötung auf verlangen" OR sterbehilfe OR suizidbeihilfe OR suizidassis* OR "hilfe zum suizid" OR "hilfe zur selbsttötung")) AND TX (((MH "Decision Making, Patient+") OR (MH "Decision Making+") OR interact* OR communic* OR counsel* OR choice* OR guid* OR support* OR interaktion* OR kommunikation* OR beratung* OR entscheidung* OR begleit* OR unterstütz*))*

**Records found in database searches**

Date of search (for all databases): April 20, 2022

| | n |
|---|---:|
| Web of Science | 5,560 |
| PubMed | 3,669 |
| CINAHL | 393 |
| **No. of records found in database searches [a]** | **9,622** |
| Duplicates found by Endnote | - 1,303 |
| Duplicates found by hand | - 405 |
| **Total no. of records (after removal of duplicates)** | **7,914** |

Notes: [a] All records found were imported to Endnote.

### B Secondary search strategies

**References of reviews found in database searches**

All review articles found in the literature search meeting inclusion criteria were excluded from data extraction, but screened for additional relevant records to include in the review (see table below). All records identified through this secondary search strategy had already been included via the primary search.

| DOI of the review | No. of references | N/a for screening | Excluded during title & abstract screening | Excluded during full text assessment | **Included [a]** |
|---|---:|---:|---:|---:|---:|
| 10.1017/S0714980818000600 | 21 | -3 | -7 | -1 | **10** |
| 10.1016/j.ijnurstu.2007.06.009 | 35 | -9 | -22 | -1 | **3** |
| 10.1136/jme.2003.004028 | 15 | -2 | -12 | 0 | **1** |
| n/a (DeZeeuw&Myers, 2020) | 24 | -23 | 0 | -1 | **0** |
| 10.1177/0269216319857630 | 19 | -4 | -5 | -2 | **8** |
| 10.1002/ijop.12680 | 20 | -4 | -14 | -1 | **1** |
| 10.1177/0269216311397688 [b] | n/a | n/a | n/a | n/a | **n/a** |
| 10.1136/bmjspcare-2019-002018 | 23 | -1 | -11 | -1 | **11** |
| 10.1186/s12910-019-0361-2 | 66 | -20 | -34 | -1 | **11** |
| 10.1136/bmjopen-2017-016659 | 14 | -2 | -11 | -1 | **0** |

Notes: [a] All these records were already included though the primary search strategy, no additional records identified; [b] deemed not applicable for screening of referenced records, as this was a review of attitudes towards legalization.

**Annual country reports**

| Country | Name of report | n |
|---|---|---:|
| Canada | Medical Assistance in dying | 7 |
| Oregon | Death with Dignity Act | 24 |
| Netherlands | Regional euthanasia review committees | 18 |
| | **Total no. of records** | **49** |

### Additional file 3 | Title & Abstract Screening

***Note:*** AS/VE were replaced by AID during the preparation of the manuscript to align with the terminology used in the publication.

##### *Inclusion criteria*

1. The subject of the paper is aid in dying (AID, including assisted suicide and/or voluntary euthanasia) (compare list of synonyms/related terms).
2. The paper reports on decision-making processes on AID between people being involved (i.e. the person with a wish for hastened death, relatives, health care professionals (HCPs), others directly involved), after the person decided to seek information about AID.
3. The paper reports on AID for humans.
4. The paper reports on a project or study that collected own results (regardless of the scientific method and study design used).
5. The paper can report theoretical and/or emprirical findings.
6. The paper can report results about current state and/or needs assessments.

##### *Exclusion criteria*

1. The language of the paper is not English or German.
2. The paper reports only on decision-making processes on AID on a broader society level (e.g. political debate, legal matters).
3. The paper reports on AID for any other group but human beings (e.g., animals).
4. The paper is an opinion piece (inculing editorials, letters, commentaries) that does not report own results.
5. The paper is a study protocol that does not report results.
6. No author can be found for the paper.
7. No year of publication can be found for the article.
8. The publisher/journal is no scientific publisher.

##### Screening process

- Initially, one assessor each screened every record. Subsequently, all records with uncertainty (i.e., rated "maybe") were discussed between all assessors.
- Assessors: CG/Claudia Gugel, ZH/Zoe Henning, RR/Robert Reincke (see acknowledgements)
- Screening completed 28 June 2022

| | PART A | PART B | PART C | TOTAL |
|---|---|---|---|---|
| Data set | author initials A to L, records without author, annual country reports | author initials M to R | author initials S to Z | |
| Assessor | CG | ZH | RR | n/a |
| **Total no. of records screened** | 4,299 | 1,759 | 1,905 | **7,963** |
| **First round of title & abstract screening (n=7,963)** | | | | |
| Excluded | - 4,087 | - 1,661 | - 1,836 | **-5,934** |
| Full texts needed | 193 | 51 | 37 | **281** |
| Maybe | 19 | 47 | 32 | **98** |
| **Consensus discussion of all "maybe" records (n=98)** | | | | |
| Additional exclusions | -14 | -28 | -21 | **63** |
| Additional full texts needed | 5 | 19 | 11 | **35** |
| **Summary** | | | | |
| **Total no. of records excluded** | -4,101 | -1,689 | -1,857 | **-7,647** |
| **Total no. of full texts needed** | 198 | 70 | 48 | **316** |

### Additional file 4 | Full-Text Assessment

#### Assessment process

- Five records assessed jointly by two assessors (CG, ZH)
- Three rounds of double assessment between CG, ZH, and PH of a total of 55 records
- Single assessment of the remaining data set (n=256 records)
- In case of uncertainty during single assessment, double (n=29 records) or triple (n=5) assessment until consensus was reached.

***1st round of double assessment (10% of full texts assessed)***

Overlap between CG and ZH (27 full texts in total):

| | Inclusion or exclusion | Which in-/exclusion criterion [a] |
|---|---|---|
| Agreement between assessors | 23 (85 %) | 4 (15 %) |
| Disagreement between assessors | 4 (15 %) | 11 (41 %) |

Notes: [a] Only applicable if assessors agreed to exclude.

The first round of double assessment and subsequent consensus discussion between CG and ZH led to small revisions in the exact wording of the inclusion and exclusion criteria and an instruction of how to use the criteria. It was agreed to have a second round of double assessment (5% of records) to check whether agreement could be increased.

***2nd round of double assessment (5% of full texts assessed)***

Overlap between CG and ZH (14 full texts in total):

| | Inclusion or exclusion | Which in-/exclusion criterion [a] |
|---|---|---|
| Agreement between assessors | 8 (57%) | 2 (14 %) |
| Disagreement between assessors | 6 (43 %) | 3 (21 %) |

Notes: [a] Only applicable if assessors agreed to exclude.

The second round of double assessment showed less agreement between the two assessors. A consensus discussion between ZH and PH showed that one disagreement was due to a mistake of one of the assessors (decision for exclusion although no reason for exclusion was found). This led to an improved agreement rate of 64% and disagreement rate of 36%. The consensus discussion led to small additions in one exclusion criterion and the general agreement that minimal presence of the inclusion criteria 2-4 was sufficient for inclusion. It was agreed to have a third round of double assessment (5% of records) to check whether agreement could be increased.

***3rd round of double assessment (5% of full texts assessed)***

Overlap between PH and ZH (14 full texts in total):

| | Inclusion or exclusion | Which in-/exclusion criterion [a] |
|---|---|---|
| Agreement between assessors | 12 (86 %) | 6 (43 %) |
| Disagreement between assessors | 2 (14 %) | 0 (0 %) |

Notes: [a] Only applicable if assessors agreed to exclude

The third round of double assessment showed improved agreement between the two assessors. A consensus discussion between ZH and PH showed that both disagreements were due to different understandings of the term “wish for hastened death”. This led to a clarifying addition to inclusion criterion 1. For one of the two publications, ZH had been in doubt about inclusion or exclusion during screening.

***Further assessment***

After three rounds of double assessment (see above), which led to adaptations and an increase in agreement regarding inclusion and exclusion of full texts, the team agreed to continue with single assessment using a conservative approach. This meant that whenever the single assessor, was in slight doubt about whether to include or exclude a full text, she asked a second reviewer to assess this full text and subsequently discuss it.

Twenty-nine full texts needed this form of double assessment (internal record IDs): #65, #70, #75, #77, #80, #94, #95, #99, #101, #112, #116, #134, #137, #139, #140, #141, #144, #152, #154, #168, #181, #182, #204, #216, #228, #233, #247, #259, #261 (double assessment by CG and ZH)

Five full texts needed a triple assessment: #1, #7, #38, #116, #126 (triple assessment by CG, ZH and PH)

Across all records, five full texts were excluded at a later stage (i.e., during data extraction) through consensus in the study team.

***Instructions for documentation of exclusion in the excel sheet:***

1. Use exclusion criteria prior to inclusion criteria (e.g., if study protocol without own results, no need to check if main subject is AID).
2. Only document one criterion in order of appearance listed in the inclusion/exclusion criteria list (i.e., once one criterion led to exclusion, other criteria do not need to be examined and/or documented).
3. If exclusion criteria applies, document an x.
4. If inclusion criteria does not apply, document an o.
5. All inclusion criteria must be met for a publication to be included.

The excel sheet documenting the full text assessment for each record can be obtained from the principal investigator upon reasonable request.

### Additional file 5 | Data extraction

**Aims:**
Synthesis of the current scientific understanding of interactional decision-making processes regarding aid in dying (AID, including assisted suicide (AS) and voluntray euthanasia (VE)) in the health care context.

**Research questions (RQs):**

RQ1 What are the characteristic of studies that evaluated interactional decision-making processes on AID?

RQ2 What is known about decision-making processes on AID from scientific studies?
   a. Who participates in decision-making processes on AID?
   b. What phases are described for decision-making processes on AID?
   c. What is known about aspects of communication and/or interaction during decision-making processes on AID?

RQ3 What research gaps and implications for future studies have been identified in scientific publications on decision-making processes on AID?

**Development process of the data extraction form**
The research team worked together to determine which variables should be extracted to answer the research questions. Charting was viewed as an iterative process, with reviewers continually extracting data and updating the data extraction form. One team member initially developed the data extraction form based on team discussion and then pilot tested it using two or three included records. Afterwards, two reviewers tested the extraction form on five to ten studies to determine if their approach to data extraction is consistent with the research questions and purpose.

**Key information** (cp. data extraction form)

- General information of the record: internal publication ID, author, year, title, journal, doi
- Study characteristics: concepts evaluated, study design, research question(s), study population, sample size, language, country (AID legislation)
- Results regarding interactional decision-making (DM) processes on AID:
  - Participants of interactional DM processes on AID
  - Phases/steps of interactional DM processes on AID
  - Decisions to be made during interactional DM processes on AID (who makes these decisions)
  - Type of interactional DM process
  - Content of communication during interactional DM processes on AID
  - Other aspects of communication during interactional DM processes on AID
  - Other aspects of interactional DM processes on AID
- Additional relevant aspects before or after DM processes on AID: before, after
- Future research: Research gaps / implications for future research on interactional DM processes on AID
- Assessment by study team: subjective relevance for RQs, comments

**Data extraction process**
Initial data extraction was done as close to the original publications as possible. Results were extracted, but not interpreted by the researchers.

After a discussion regarding data extraction, the two raters (CG, ZH) agreed on new and more specific guidelines on how to proceed with the data extraction. The raters agreed on these following:

- Study characteristic – concepts evaluated:
  - Always write down the concept as it is referred to in the article (e.g., write down PAD (phisician assisted death) referred to in the text instead of the more common term PAS (physician assisted suicide)).
  - All concepts covered in the article should be listed.
- Study characteristic – study design: The specific name of the study design should be given.

- Study characteristic – study population: The study population refers to the group of people who were included in the study as participants (e.g., there are death certificate studies in which physicians are interviewed about deceased patients and their end-of-life decisions).
- Study characteristic – sample size: The sample size should always be provided with the indication to which group of persons the number refers to (e.g., the sample size in death certificate studies refers to deceased patients and not to the physicians participating).
- Study characteristic – country (legislation): The legislation should be noted if it is described in the text. If there is nothing in the text, then the article by Mroz et al. (2021) about the legislation can be used. Note: The year of the study is important to keep in mind, as the legislation may have changed over time. If the legal situation is unclear, the field should simply be left blank.
- To distinguish different extracted paragraphs in a column, use: (...).
- In the conclusion, duplications to other parts of the record may occur. These should not be extracted.

**Data synthesis**
The initially extracted data was subsequently reviewed and revised by the prinicpal investigator. This step included streamlining with respect to the research questions to allow further synthesis and interpretation.

Gugel C, Henning Z, Hahlweg P. Interactional decision-making processes and communication on aid in dying: an international scoping review.

**Additional file 6. Data extraction sheet.**

| # | **General information of the records**<br>author | year | title | journal | doi | **Study characteristics**<br>concepts evaluated | study design | Research question(s) | study population | sample size | language | country (legislation) |
|---|---|---|---|---|---|---|---|---|---|---|---|---|
| examples | | | | | | AS (= assisted suicide, incl. physician assisted suicide), VE (=voluntary euthanasia, also called euthanasia), AS/VE (=assisted suicide and voluntary eithanasia), MAID (medical assistance/aid in dying) | e.g., RCT, cross-sectional survey, qualitative interview study, case report, … | | e.g., patients with cancer, physicians, nurses | | ENG = English, GER = German | country X (AS legalized, VE illegal) |
| 1 | Alvarado V & Liebig B | 2015 | ["I want to die" - Dealing with suicidal wishes in primary palliative care] [GER: „Ich möchte sterben" – Zum Umgang mit Suizidwünschen in der hausärztlichen Palliativbetreuung] | Therapeutische Umschau, 72(10):643-8 | 10.1024/0040-5930/a000731 | AS, (also other end of life decisions) | qualitative study with group discussions | How do family doctors experience th | physicians (=GPs/"family doctors") | n=89 in 11 group discussions | GER | Switzerland (AS legalized) |
| 2 | Antifaeff K | 2019 | Social Work Practice with Medical Assistance in Dying: A Case Study | Health & Social Work, 44(3):185-92 | 10.1093/hsw/hlz002 | MAID | case report | | social workers, pts (w/ cancer) | n=2 pts | ENG | Canada (MAID legalized since 2015) |
| 3 | Back AL et al | 2002 | Clinician-patient interactions about requests for physician-assisted suicide: a patient and family view | Archives of Internal Med, 162(11):1257-65 | 10.1001/archinte.162.11.1257 | AS | longitudinal qualitative interview study | to describe the reasons for pt to pursue AS, the narrative of events leading to death, and interactions with physicians and other clinicians | pt (pursuing AS), their family members | n=35 pt cases (prospective: n=12 pt, n=20 family members, retrospective: n=28 family members (across 23 deceased pt)) | ENG | USA (AS legalized in some states) |
| 4 | Balint JA | 2000 | Decisions at the end of life | Croatian Medical Journal, 41(2):144-9 | n/a | AS/VE, MAID | case report | | pts (w/ cancer), physicians | n=1 pt | ENG | USA, Oregon (MAID legalized) |
| 5 | Ball I M et al | 2019 | A canadian academic hospital's initial MAID experience: a health-care systems review | Journal of Palliative Care, 34(2):78-84 | 10.1177/0825859718812446 | MAID | description of one organization's processes | to describe the manner in which London Health Sciences Center has approached local and regional requests for MAID, including the administration, ethics, privacy, and clinical process | hospital | n=1 hospital | ENG | Canada (MAID legalized since 2015) |
| 6 | Bannink M et al | 2000 | Psychiatric consultation and qualility of decision making in euthanasia | Lancet, 356(9247):2067-8 | 10.1016/s0140-6736(00)03407-3 | VE | description of one organization's processes | | hospital | n=1 hospital (reporting on n=22 pt requesting VE) | ENG | Netherlands (VE/AS legalized) |
| 7 | Booij SJ et al | 2013 | Euthanasia and advance directives in Huntington's disease: qualitative analysis of interviews with patients | J Huntingtons Dis, 2(3):323-30 | 10.3233/jhd-130060 | AS/VE (advance directives) | qualitative interview study | to gain insight into the thoughts & attitudes of pt regarding end-of-life issues & use of advance directives and to examined which difficulties pts experience when thinking about end-of-life wishes | pts (with Huntington´s disease) | n=14 pts | ENG | Netherlands (VE/AS legalized) |
| 8 | Borgsteede SD et al | 2007 | Communication about euthanasia in general practice: Opinions and experiences of patients and their general practitioners | Patient Education and Counseling, 66(2):156-61 | 10.1016/j.pec.2006.10.006 | VE | qualitative interview study | to investigate whether pts and their GPs talk about euthanasia, and if so, how they communicate about this | physicians (=GPs), pts (w/ cancer, heart failure, or COPD; life expectancy < 6 months) | n=20 GPs, n=30 pts | ENG | Netherlands (VE lagalized) |

| # | **Results regarding decision-making (DM) processes on AS/VE**<br>participants of DM processes on AS/VE | phases/steps of DM processes on AS/VE | decisions to be made during DM processes on AS/VE (who makes these decisions) | type of DM process | content of communication during DM processes on AS/VE |
|---|---|---|---|---|---|
| examples | | e.g. relationship building, information giving, discussion of preferences… | e.g., whether to support the request (physician), requesting another physicians evaluation, prescription of lethal medication | Who leads the DM process? Physician-led, shared, requestor-led, divided (physician-led or requestor-led for different aspects during the DM process), Who initiated the topic? | What is talked about? |
| 1 | GP, pt | | | | |
| 2 | pt, social worker, spouse, palliative care team, (spiritual care support - declined by pt), nurse educator | symptom control; consideration of end-of-life options; decision on initiation of assessment process; initiation of assessment process; conduction of biopsychosocial assessment; decision to host one's own wake & preparation to do so; support preparedness of family members | prognosis (palliative care team); decision to pursue MAID (patient); decision to host one's own wake (patient); is patient lacking support or coerced towards MAID (social worker) | | fears (incl. fear of dying, poor symptom amangement, loss of dignity, loneliness/feeling left); uncertainty; what does patient want to be remebered as; how does patient want to die; when is the right time for MAID; who are the most important people in patient's life (are they informed about MAID? how to inform? do they support/oppose?); what do family members need; grief<br><br>nurse educator from the Assisted Dying Program reviewed MAID medication administration process with patient and family |
| 3 | physician, pt, family members, caseworker from advocacy organizations | | | | guideline for responding to a pt requesting AS (1-4 out of 6, for 5-6 see other aspects of communication): 1) address request explicitly & openly; 2) ask what kind of death pt would like to have; 3) after understanding pt wishes and expectations, offer to discuss how dying could be managed; 4) check pt perception by asking "What are you taking away from our talk today?"; also discuss other worries, fears, vunerabilities abour illness & dying; physicians sometimes focus too much on legality of AS; physicians bring too much of their own feelings & concerns about AS into the conversation; physicians sometimes want to maintain a biomedical focus and are unwilling to talk about AS; sometimes physicians say "you do not want to die like that" which might be understood by patients as endorsement of AS |
| 4 | physician, pt | Patient visits her physician to seek his help in committing suicide when suffering becomes unbearable; physician presents treatment options/alternatives to AS & patient voices preference; physician considers request for MAID incl/ second involvement of physician; pt putting request in writing and meeting w/ family; physician provides prescription for Phenobarbital, patient fills it and keeps meds in her safe | decision on medical indication for AS (physician & second physician/oncologist); decision on provision of prescription (physician) | divided; pt requests AS | medical indication/prognosis; need for condultation w/ second physician; written requests & family meeting; pt preferences (withholding high technology treatments, being tired, enjoy time w/ family, die at home in peace); quality of life issues; contextual issues |
| 5 | pt, health care providers (for referral), hospital's internal resource group (multidisciplinary committee, incl. hospital ethicist, medical advisory committee member, senior leadership, departmental leadership, pharmacy leadership, professional practice disciplines, legal councel, Director of Patient Safety and Experience), physician providers ( MAID assessors, MAID providers), pharmacy, local homecare nursing, local catholic hospital (for referrals), venous access support team | recognizing the request and referral (health care providers) | | | |
| 6 | pt, primary physician, psychiatrist | | Does pt have decision making capacity? (physician, psychiatrist); Is wish longlasting and wee-considered & not from undue pressure or intolerable pain? (physician, psychiatrist); did pt get adequate and understandable information about medical situation? (physician, psychiatrist, pt); does pt have current psychiatric disorder? (psychiatrist) | | |
| 7 | pt (w/ Huntington's disease), physicians (e.g. GP or at nursing home), family members | | | | pt end-of-life wishes; (im)possibilities of advance directives; information about AS/VE |
| 8 | pt, GP | communication between pt and GP about potential situations and preferences might give rise to VE requests; written VE request by pt; detailed discussion as a prerequisite to the communication process about VE | who initiates VE talk? (GP & pt); vague general requests: is what the pt voices a request for VE? (GP); support of VE request (GP); at what point does life become unbearable (mostly pt) | initiation of talk about VE: mostly pt-initited, sometimes GP initiated if GP foresees problems in near future | when will be the point in time at which VE will be the best option? (when is suffering unbearable?) |

| # | other aspects of communication during DM processes on AS/VE | other aspects of DM processes on AS/VE | **Additional relevant aspects before or after DM processes on AS/VE**<br>before | after | **Future research**<br>Research gaps / implications for future research on DM processes on AS/VE | **Assessment by**<br>Subjective relevance for RQs | Comments |
|---|---|---|---|---|---|---|---|
| examples | e.g., How many consultations? Who was involved? Where did communication happen? | e.g., consequences on the physician-patient-relationship, role of different health care providers | e.g., what happens before a person considering AS/VE gets in contact with the professional support system | e.g., what happens after a person executed AS/VE | | complete match; partial match; low match | |
| 1 | Need for professional strategies to overcome conflicts of values and attitudes between patients and physicians; GPs react in three ways to personal conflicts: "deliminating behavior" (GER "abgrenzendes Verhalten"), "empathic adaptation" (GER "empathische Anpassung"), or "convergence of perspectives" (GER "Perspektivenannäherung"); "convergence of perspectives" helps creating a common basis for negotiation through continued negotiation despite differing ideas | GPs may experience personal conflicts if patients ask for asissted suicide; "convergence of perspectives" with patients can reduce strain for GPs & guarantee high quality of care | | | | partial match | |
| 2 | offer a confidential place to process thoughts and emotions to gain clarity; offer psychososical and spiritual support; social worker maintained stance of neutrality toward MAID to support patients self-determination; family meetings were helf | | | follow up with family; offer referrals to community-based bereavement supports to family | Need for qualitative studies that examine the client experience throughout the MAID process, a population currently underserved by social work research; engaging clients as co-researchers will generate invaluable insight and guidance for developing a patient-centered model of psychosocial care for clients pursuing MAID | complete match | |
| 3 | providers willingness & openness to discuss AS is important (facilitates complete evaluation of request); guideline for responding to a pt requesting AS (5-6 out of 6, for 1-4 see content of communication): 5) remember: process of building therapeutic relationship more important than providing lethal prescription; 6) monitor yourself for underinvolvement or overinvolvement in the clinician-patient relationship | AS reqeust is challenging for therapeutic relationship (can be maintained even if clinician is not willng to praticipate in AS); clinicians self-awareness of emotional needs and vulnerabilities needed; clinicians need to maintain appropriate boundaries; avoid prescriptions without thorough evaluationa nd discussion of AS; physicians need to know which medication, dose, & how to administer; importance to addess request over time; request often includes wish to for therapeutic alliance and have meaningful conversations | clinician openness to discuss AS & expertise in dealing w/ dying process make continued therapeutic relationship possible | family members want closure with clinicans (if meaningful therapeutic relationship existed) | | complete match | changed from partial match |
| 4 | | | | pt died w/o taking Phenobarbital; knew that it was ther if she needed it | | partial match | changed from complete match |
| 5 | internal resource group acts as intermedicary between requestors and provider group; MAID assessments should be done in hospital, private residence, or long term care facility | providers being aware & well informed is essential; effective referrals are essential (incl. referral hotline, CAVE: delays); internal resource group supports referrals from patients & health-care providers from within our hospital, the city & our region; have designated areas for MAID procedure (privacy, quiet etc.) | | | | low match | |
| 6 | | VE request might be denied as result ofpsychiatrist's evaluation; staff appreciated expertise, help, & support of consultant psychiatrist (might add to decision-making quality for some cases) | benefits and disadvantages of mandatory involvement of psychiatrist should be weighed | | | low match | |
| 7 | pt seemt o want to talk about end-of-life wishes, but sometimes do not know how; physicians should actively ask all their pt about end-of-life wishes; discussing end-of-life wishes requires good and trusting relationship; end-of-life wishes should eb discussed early in the course of disease; family members & friends are important in the process | pt need awareness of possibility to write advance directive; some pt w/ Huntington's disease do have advance directives for VE (incl. threshold of acceptable life) | | | | low match | changed from partial match |
| 8 | long-standing GP-pt-relationship as advantage; pt home as setting as advantage; not all pt needed VE talk; VE talk more frequent w/ pt w/ cancer (than other diagnoses, potentially due to more uncertain prognoses) | | | | | complete match | |

| | | | | | | | | | | | | | | | | | | | | | | | | |
|---|---|---|---|---|---|---|---|---|---|---|---|---|---|---|---|---|---|---|---|---|---|---|---|---|
| 9 | Brown J et al | 2020 | Care considerations in a patient- and family-centered medical assistance in dying program | Journal of Palliative Care, 37(3):11 | 10.1177/0825859720951661 | MAID | qualitative interview study | to explore patient, family, and healthcare provider experiences with MAID to enhance high-quality patient- and family-centered MAID care | pts, family members, HCPs | n=5 pts, n=11 family members, n=14 HCPs | ENG | Canada (MAID legalized since 2015) | patient, family members, team-based approach for health care providers (physicians, palliative care nurse, occupational therapists, physiotherapists, speech therapists, social workers) | continuous investment in relationship by HCPs; anticipatory guidance for family members; team meeting for HCPs; MAID team should eb commissioned early in trajectory of lifelimiting illness to optimize support and care options for pt & family (before official request & assessment) | eligibility assessment (HCP); choosing an appropriate location & setting for pt and family (pt, family); whom to inform about MAID plans (pt) | | acknowledge the person behind MAID request and keep human life as focus of assisted death; acknowledge all forms fo loss (incl/ physical abilities, quality of life); anticipatory guidance: information about the phsical process fo dying (for family members); location & setting for MAID; emotional processing (desperation, fatigue, fear) | HCPs must offer enough time for pts & family to share and discuss emotional pathway towards MAID; sensitivity in eligibility assessment (potentially multiple encounters); pt & family need time to understand & debrief; interdisciplinary care as significant care asset | process not simple or well-defined; pts &families are balancing quality of life, MAID timing, capacity for final consent, & practitioner availability; success of MAID experience rested heavily on relational approach of HCP & intensive investment in relationship with pts, families, and other care team members (relational investment as demonstration of caring); judgemental statements from HCPs can be harmful; faith-influences policies complicate process (e.g. location chosing); pts & families grateful that choices were respected & honored by closest family & MAID healthcare team | | debriefing the circle of support; emotions of family members; emotions of HCPS (team-absed approach might offer support for HCPs) | continuously evaluate patient- and family-centered care approaches to MAID (including outcomes) | complete match | 28&29: same sample, different RQ |
| 10 | Brown J et al | 2020 | Medical assistance in dying: patients', families', and health care providers' perspectives on access and care delivery | Journal of Palliative Medicine, 23(11):1468-77 | 10.1089/jpm.2019.0509 | MAID | qualitative interview study | to explore pt, family, and health care provider (HCP) perspectives on MAID access and care delivery and improve regional MAID care delivery | pts, family members, HCPs | n=5 pts (4 MAID eligible, 1 not MAID eligible), n=11 family members, n=14 HCPs | ENG | Canada (MAID legalized since 2015) | pt, family members, primary HCPs (physicians, nurses, spiritual care ppl), CEO of nursing home, MAID providers, Worker'S Compensation Board (regarding MAID coverage for work related condition) | Beginning the conversation (How and to whom to bring up the topic?) | whether to bring up the topic (pt, family members); whether to refer to MAID providers (primary HCPs) | | anticipatory guidance to pts, families, & care team re/ final consent process (incl options for care if risk of loss of capacity) | who brings up MAID?; who directs the process? (nurses? physicians?); | have a centralized MAID program access point; religion plays a role (e.g. Catholic institutions); who pays for MAID?; teams need to know who has conscientious objection (possibly adjust scheduling correspondingly) | establish a centralized MAID program access point; know as a team who has a conscientious objection; need for interdisciplinary education for HCPs (incl/ legal knowledge, role & scope of practice of interdisciplinary team members, procedures & care standards, sensitive & holsitic communication, respect for staff choices to participate/object; open & respectful dialogue btw/ MAID programs and institutions | clear processes for post-death reporting | | partial match | 28&29: same sample, different RQ |
| 11 | Buchbinder M et al | 2019 | Health care providers' experiences with implementing medical aid-in-dying in Vermont: a qualitative study | Journal of General Internal Medicine, 34(4):636-41 | 10.1007/s11606-018-4811-1 | MAID | qualitative interview study | to describe Vermont HCPs' experiences practicing under the Patient Choice and Control at End of Life Act | HCPs (incl/ physicians, nurses, social workers) | n=37 HCPs (n=19 physicians, n=16 nurse, n= 2 social workers) | ENG | USA, Vermont (MAID legalized) | pt, physicians, nurses, social workers, advocacy organizations (for general information) | | Is criterion of prognosis of less than 6 months given? (physicians); Who is responsible and reachable in case of adverse events? (nurses, physcians) | | assuring pt that HCPs would be there for them; exploring request and reasons for request; worries about finances and being a burden; untreated symptoms such as pain and depression; discuss alternative options (incl/ hospice, palliative care, VSED); after pt initiated conversation about MAID, HCPs feel more free to adress end-of-life concerns and care in general (incl/ alternatives); plan for the day of MAID; how to prepare the mediaction; possible complications (such as not being able to swallow the medication) and prepare for that | HCPs should do everything they can so pt do not feel like they need to use MAID; sufficient time needed for MAId conversations; HCPs need to find support from other HCPs w/ more experience in MAID | prescribing the medicine is physicians' task; nurses & social workers helped with administrative tasks (e.g. completing paperwork, finding witnesses, finding second physician, identify pharmacies, determine insurance coverage); HCPs need to make decision if they were willing tobe present during MAID; good prepapration for MAID procedure and potential complications is needed; nurses & social workers role seems to be critical, but underrecognized | | | | partial match | 30&31: same sample, different RQ |
| 12 | Buchbinder M et al | 2018 | Caregivers' experiences with medical aid-in-dying in Vermont: a qualitative study | Journal of Pain and Symptom Management, 56(6):936-43 | 10.1016/j.jpainsymman.2018.08.010 | MAID | qualitative interview study | to explore the experiences of lay caregivers involved with MAID in Vermont, focusing on the day of death | family members ("caregivers" incl/ family members, close friends, other individuals who were closely involved with the care) | n=19 caregivers (who cared for 11 pts) | ENG | USA, Vermont (MAID legalized) | pt, caregivers, HCPs | | | | what does pt want at end of life; who should be present during MAID; what does pt expect dying to be like | including caregivers in education & planning, where warranted, can mitigate feelings of unpreparedness & ensure a smoother experience for everyone | | | | | low match | changed from partial match; 30&31: same sample, different RQ |
| 13 | Carlson B et al | 2005 | Oregon hospice chaplains' experiences with patients requesting physician-assisted suicide | Journal of Palliative Medicine, 8(6):1160-6 | 10.1089/jpm.2005.8.1160 | AS | cross-sectional quantitative surevy | to survey hospice chaplains regarding their views on the Oregon Death With Dignity Act and experiences working with patients who request AS | hospice chaplains | n=50 | ENG | USA, Oregon (MAID legalized) | pt, hospice chaplain | | | | alternatives to AS; teaching hospice phiylosophy; role of faith & spirituality; reasons for wanting AS; family concers ans reactions; what might be gained from natural dying process; difference betw/ suicide and AS; concerns about AS process; preparation for circumstances of AS procedure; clarification of pt position on AS | nonfearing presence of chaplains; non-judgemental support; relationship easier if pt had prior positive relationship w/ clergy or chaplain was involved before AS request; chaplains would not stop caring for pt b/c of choice for AS | chaplains did not want to influence pt decision to pursue AS | hospice chaplains have different opinions reg/ AS; high value to support pt irrespective of what they decide reg/ AS | | | low match | changed from partial match |
| 14 | Chambaere K et al | 2011 | Trends in medical end-of-life decision making in Flanders, Belgium 1998-2001-2007 | Medical Decision Making, 31(3):500-10 | 10.1177/0272989x10392379 | VE/AS, (also other end-of-life decisions such as non-treatment, intensified pain management, life-ending w/o pt request) | cross-sectional quantitativ survey | to examine trends in the occurrence and decision-making process of end-of-life practices in different patient groups since these legal changes | physicians (identified through a stratified random sample of death certificates) | n=2,540 physician- | ENG | Belgium, Flanders (VE legalized since 2002) | physician, colleague physician, pt, nurses, pt relatives | | | | | | open atmosphere for end-of-life discussions was created by legalization of VE (incl/ earlier discussion in disease trajectory) | | | | low match | changed from partial match |
| 15 | Chambaere K et al | 2015 | End-of-life decisions in individuals dying with dementia in Belgium | Journal of the American Geriatrics Society, 63(2):290-6 | 10.1111/jgs.13255 | VE, (also withholding or withdrawing life-prolonging treatment) | cross-sectional quantitative survey | to describe the prevalence and characteristics of end-of-life decisions in individuals dying with dementia in Belgium | physicians (identified through a large random sample of death certificates) | n=1,637 physician-reported deaths (incl/ n=361 dementia pt, n = 1,276 cancer pt) | ENG | Belgium, Brussels & Flanders (VE legalized since 2002) | pt, physician, colleague physician, nursing team, palliative care specialist, family members, caregivers | | Does pt have decision-making capacity? (physician) | | | | pt w/ dementia often do not have decision-making capacity and thus are not actively involved in end-of-life decision-making; 1.3% of pt w/ dementia had made VE request, but VE did not occur for any pt | | | empirical evidence on whether to include pt w/ dementia in the scope of the Belgian euthanaisa law | low match | changed from partial match |
| 16 | Claxton-Oldfield S & Beaudette S | 2021 | Hospice Palliative Care Volunteers' Attitudes, Opinions, Experiences, and Perceived Needs for Training Around Medical Assistance in Dying (MAiD) | American Journal of Hospice & Palliative Medicine, 38(11):1282-1290 | 10.1177/1049909120985130 | MAID | cross-sectional quantitative survey | to exame hospice palliative care vounteers' attitudes, opinions, experiences, & perceived needs for training around MAID | volunteers (hospice palliative care (HPC) volunteers from 2 HPC volunteer programs (one community-based, one residential)) | n=48 HPC volunteers | ENG | Canada (MAID legalized since 2015) | pt, hospice palliative care volunteers | | | volunteers should not raise the issue of MAID; sometimes patients initiate conversation about MAID with volunteers | | volunteers should respond with open-ended questions to MAID inquiries; volunteers should react respectful and supportive; volunteers should not engage in conversations/discussions about MAID and not voice their personal beliefs; volunteers should refer to medical staff for inquiries about MAID (after asking pt permission) and to their volunteer supervisor | 96% of volunteers wanted to be able to object to participation | existing lack of policies for volunteers regarding MAID converations | Volunteers support grief and bereavement support after MAID | assess the effectiveness of training module designed to improve HPC volunteers' knowledge about and skills and confidence in responding to patients or patients' family members/caregivers who want to have a conversation with them about MAiD; nationwide survey on MAID training and policies in volunteer services | partial match | |
| 17 | Cohan J et al | 2007 | End-of-life decision-making in Belgium, Denmark, Sweden and Switzerland: does place of death make a difference? | Journal of Epidemiology and Community Health, 61(12):1062-8 | 10.1136/jech.2006.056341 | AS/VE, (also other end-of-life decisions such as continuous deep sedation, life-ending w/o pt request, pain/symptom management, non-treatment, taking into account/partly intending/explicitly intending life-shortening) | cross-sectional quantitative survey | to examine differences in end-of-life decision-making in patients dying at home, in a hospital, or in a care home | physicians (identified through representative sample of death certificates) | n=12,492 physician-reported deaths (n=2,950 in Belgium; n=2,939 in Denmark; n=3,248 in Sweden; n=3,355 in Switzerland) | ENG | Belgium (Ve legalized since 2002), Denmark (AS/VE illegal), Sweden (AS/VE illegal), Switzerland (AS legalized) | | | | | | | differences in decision-making outcome found across different settings (e.g., hopsital, home) | | | investigate differences in end-of-life decision-making between settings; explore information given on the course of dying | low match | changed from partial match |
| 18 | Cohan J et al | 2014 | Nationwide survey to evaluate the decision-making process in euthanasia requests in Belgium: do specifically trained 2nd physicians improve quality of consultation? | BMC Health Services Research, 14:9 | 10.1186/1472-6963-14-307 | VE | cross-sectional quantitative survey | to assess quality of consultations and compare these between LEIF (Life End Information Forum) and non-LEIF consultants | physicians (who had recieved VE request, incl/ general practice, anaesthesiology, gynaecology, internal medicine, neurology, pulmonary medicine, gastro-enterology, neuropsychiatry, psychiatry, cardiology, radiotherapy, surgery) | n=244 physicians | ENG | Belgium, Brussels & Flanders (VE legalized since 2002) | pt, physician, second independent physician (some specifically trained by Life End Information Forum, LEIF) | | | | | | requirements for independet second physician not always achieved; some differences in procedures and decisions made between LEIF physicians and non-LEIF physcicians; LEIF physicians statistically significantly more likely to deem pt eligible for VE (compared to non-LEIF); not statistically significant trend towards VE being carried out being more likely if LEIF physician was consulted (compared to non-LEIF pfysician) | | | | low match | changed from partial match |

| | | | | | | | | | | | | | | | | | | | | | | | |
|---|---|---|---|---|---|---|---|---|---|---|---|---|---|---|---|---|---|---|---|---|---|---|---|
| 19 | Cole RM | 1993 | Communicating with people who request euthanasia | Palliat Med, 7(2):139-43 | 10.1177/026921639300700208 | VE | case report | to demonstrate widely varied motives or hidden agendas for requesting euthanasia | pts (case 1: 46 y/o, Chinese, female, carcinoma of cervix; case 2: 33 y/o, man, HIV-positive; case 3: 35 y/o, man, AIDS; case 4: 52 y/o, man, advanced AIDS) | n=4 pts | ENG | not specified (likel | pt, physician, family members, spiritual caregivers (e.g. Buddhist monk), pro-euthanaisa orgniazation, other medical practitioners | longitudinal process; initially open up conversation about request w/o avoiding the issue; hear all issues/resonst of pt; discuss all issues/resons; then come back to initial request for VE and re-evaluate pt perspective | | pt initiate talking about VE | try to understand pt reasons for request and explore feelings; offer alternatives (such as palliative care); explore spirituality; explore ideas about death and dying; repeatedly presenting ethical and legal arguments against VE might escalate conflict; pt prognosis | give pt room to express their views, values, resons, feeling first; pt angry and resentful towards physicians after ethical resentment of VE request; potentially talk to pt and family members seperately | | | | | partial match |
| 20 | Coscarelli A et al | 2017 | Does psychosocial oncology have a greater role in aid-in-dying prescriptions than assessing decisional capacity? | Psycho-Oncology, 26(S1):38-9 | 10.1002/pon.4353 | MAID | description of one organization's processes (conference abstract) | to examine the role of psychosocial oncology in achieving optimal end of life care | hospital (clinical consultants (i.e., psychologists, social workers), pts being cared for following an institutional guideline) | n=1 hospital (n=40 pts) | ENG | USA, California (MAID legalized since 2016) | clinical consultant (psychologist, social worker), pt, families, physicians | meeting with psychosocial clinical consultant (as one step in MAID process) | | | clinical consultant meets w/ pts/families, providing a comprehensive packet of materials (education on the law, clarity in choices, resources, creating legacies, preparing for death, expectations for death for loved ones, bereavement, palliative care, safety and disposal of medication, assesses, counsels, educates, and supports) | | added value in this expanded role of psychosocial clinical consultants (incl/ detailed conversations around meaning, legacies, and ethical wills, assistance in preparation w/ end-of-life planning for pts/families, earlier referral to hospice and palliative care); may expand conversations about end of life and lead to better quality of life and prepared deaths | implement broad role for psychosocial oncology in MAID processes, mandated by institutional policy; extensive role for clinical consultants (psychologists and social workers) to facilitate quality end-of-life care including psychosocial assessment, counseling, psychoeducation, and integrated physician communication | | | low match |
| 21 | Crumley ET et al | 2021 | How is the medical assistance in dying (MAID) process carried out in Nova Scotia, Canada? A qualitative process model flowchart study | BMJ Open, 11(7):1-13 | 10.1136/bmjopen-2021-048698 | MAID | qualitative interview study (process model flowchart study) | How is the MAID process carried out and which professionals are involved at which points? Which roles and activities do professionals carry out during MAID? | HCPs (=individuals professionally involved in MAID (physicians, nurse practitioners (NP), nurses, pharmacists, healthcare administrators and advocates)) | n=12 physicians, n=3 nurse practitioners (NP), n=6 nurses, n=6 pharmacists, n=5 healthcare administrators and advocates | ENG | Canada (MAID legalized since 2015) | pt; family members; HCPs (e.g. family clinician, surgeon, nurse practitioner, nurses); MNN (MAID nurse navigator); pharmacist | Flowchart Process Model with 5 stages: 1) starting the MAID process, 2) MAID assessments, 3) MAID preparation (hospital in-patient, hospital outpatient, non-hospital), 4) day of MAID, 5) post-MAID (hospital in-patient and outpatient, non-hospital, after leaving setting) | eligibility assessment (clinician); timepoint for MAID (pt) | MAID conversations initiated by pts | 1) starting the process: pt may seek information themselves, physicians may give information or refer to MNN, MNN has central coordinating and informing role;<br>2) MAID assessments: two assessments need to indicate elegibility (assessment form returned to MNN), assessments take 45min to 5hrs, first assessor dicusses the process w/ pt & family (including legislation, criteria, timeline);<br>3) divided in hospital in-patient, hospital out-patient, non-hospital: prepare location & medication (time- and work-intensive for pharmacists with little/no compensation), find nurse/physician willing to administer the medication;<br>4) day of MAID: After determining th pt has capacity to request MAID, clinician explains procedure including what each medication does (if the patient wishes to know), dying process & potential medication responses (i.e. twitching). Only if pt still wishes MAID, clinician administers medications over 10–15min. | MNN has centras role in MAID process: conducts intake interview & stays in contact with & gives information to pt, clinicians, circle of care throughout the process. | 19 points where MAID process migth stop or be delayed; clinicians might refer/transfer pt to MNN or other clinician if not comfortable participation in MAID process; pts can stop MAID process at any time & pursue other options OR pursue multiple options simultaneously; pts need to sign MAID consent form with two indipendent witnesses present; differences in reimbursement for physicians & NPs | | 5) after MAID (divided in hospital, non-hospital, after leaving): clinician documents MAID process & death; body is being transferred; possibly staff debriefing; MNN documents MAID statistics; MNN stays in contact with family; clinicians dispose of left-over medications & equipment | compare and contrast different process models; participant observations throughout the process to understand each participants role in MAID; conduct interviews with different professionals, pts, informal support system to obtain further insights into the processes; incorporation of pt-centered care in MAID process | complete match |
| 22 | D'Angelo A et al | 2019 | Assessing genetic counselors' experiences with physician aid-in-dying and practice implications | Journal of Genetic Counseling, 28(1):164-73 | 10.1002/jgc4.1047 | MAID | qualitative interview study | to explore genetic counselors' experiences, understanding, training, and perspectives about MAID | genetic counselors (from US states where MAID is legal) | n=15 genetic counselors | ENG | USA, California, Oregon, Washington (MAID legalized in 1997 (Oregon), 2008 (Washington), 2016 (California)) | pt, genetic counselors, family members, other physicians/providers | | | pt initiate MAID conversation w/ genetic counselors | | genetic counselors may feel surprised/unprepared when MAID is brought up (b/c of lack of understanding of legal situation, no prior experience, worried about unintentionally misinforming pts); predictive genetic testing & its results may initiate pt thinking/asking about MAID; recurrent or metastatic cancer may initiate pt thinking/asking about MAID; changes in legality of MAID may initiate pt thinking/asking about MAID | genetic counselors may encounter pts early on disease trajectory (potential for long-term MAID considerations/planning); some participants thought other HCPs might be more helpful for MAID conversations; conscientious objection combined w/ referral seen as acceptable for genetic counselors; genetic counselors could be valuable converation partners for MAID requests because of ability to foster empathic and trusting relationships (incl/ used to encounter challenging emotions, 60-minute consultations), experience and skills in addressing challenging topics (incl/ emotions, counseling and advocacy), expertise in value-based shared decision-making, ability to refer to other providers; pt might have witnessed other family members suffer from same genetic disease; genetic counselors acknowledged lack of knowledge about MAID; MAID conversations might arise w/ pt who are not eligible for MAID | training needs on MAID for genetic counselors | | further characterize how and whether MAID discussions are occurring across genetic counseling contexts, including larger sample sizes, more diverse specialties, and greater geographic representation; research recording and analyzing conversations between genetic counselors and pts about MAID could provide important information to suggest best practices; to develop and test educational tools for genetic counselors about MAID | partial match |
| 23 | Damron L et al | 2022 | Physician-assisted dying: access and utilization in patients with movement disorders | Movement Disorders, 37(4):694-8 | 10.1002/mds.28964 | AS | description of one organization's processes (incl/ pt characteristics, qualitative data from HCPs, case vignettes) | to describe our experiences with AS as a tertiary care center for movement disorders and discuss the need for amendments to make the program more accessible to patients with these neurodegenerative diseases | hospital (=tertiary care center for movement disorders) | n=1 institution (incl/ n = 12 pt cases) | ENG | USA, California (MAID legalized since 2016) | pt, families, friends, clinician, MAID coordinator; interdisciplinary team (physicians, social workers, psychologists, pharmacists, nurses, chaplains) | preparing/practicing for MAID | request to be evaluated/re-evaluated (pt); Does pt meet criteria? (physician); | pt initiate topicof MAID | MAID coordinator supports by teaching about the inclusion criteria, help with identifying appropriate HCPs to certify required steps, ensuring state-required paperwork is completed, & psychosocial support for both pats & families | include MAID in ongoing discussion (after depression & adjustment disorder have been adressed); advanced care planning may bring up MAID sooner in disease trajectory | pt w/ motor disorders might inquire about MAID before meeting criteria | MAID program in place (consisting of MAID coordinator, specially trained social worker, physician advisor); palliative care may be particularly helpful for such MAID programs; education for HCPs on regular basis | | | partial match |
| 24 | Davila SLR et al | 2010 | Management of a request for physician-assisted suicide | American Journal of Hospice & Palliative Medicine, 27(1):63-5 | 10.1177/1049909109341874 | AS | case report | to present a case in which a patient with end-stage prostate cancer requested AS, with AS not being legal | pt (male, 84yrs, diagnosis of terminal prostate cancer) | n=1 pt | ENG | USA, Florida (AS not legal) | pt, physicians (incl/ psychiatrist) | long-standing belief in right to AS; transfer to hospice & request for AS | Is there evidence of psychiatric disorder or cognitive impairment? (physicians); Is request for AS caused by psychiatric disorder/symptoms? (physicians) | pt initiated topic AS | | need for comprehensive evaluation to properly assess & manage untreated physical & emotional suffering that may influence pt's decision to hasten death | | pt had long-standing personal belief in a person's right to commit suicide, including AS, in the face of terminal illness; pt had witnessed family member die from cancer; pt had been member of MAID advocacy organization for long time; clear standards of medical & psychiatric evaluation need to be incorporated into AS legislation | | | low match |
| 25 | De Bal N et al | 2006 | Involvement of nurses in caring for patients requesting euthanasia in Flanders (Belgium): A qualitative study | International Journal of Nursing Studies, 43(5):589-599 | 10.1016/j.ijnurstu.2005.08.003 | VE | qualitative interview study | to explore in detail nurses' experiences about their involvement in caring for people who request euthanasia | nurses | n=15 nurses (with different specialties) | ENG | Belgium (VE illegal at time of study, VE legalized after the study) | nurses, pts, family members, physicians, other HCPs | Different, not strictly separable phases: 1) before VE request; 2) confronting the VE request; 3) reporting request to other caregivers; 4) participating in decision-making (within medical team); 5) result of decision-making process (within the team); 6) aftercare | interpretation of (sometimes vague) VE request & determination of authenticity (nurses); formal decision about VE request (physician) | discussion and decision on VE request (physcians & nurses) | 1) before VE request: nurses with close relationship w/ pt sometimes "sense something brewing up", listen carefully & try to understand pt perspective;<br>2) confronting the VE request: careful listening (enable pt to tell story & determine reason(s) behind request), taking seriously & compassionate care by nurses who were aware of VE request (irrespective of their attitude towards VE); possibly also listening to family members; informing about palliative care principles, care alternatives, & dying; reassuring support; maximizing pain & symptom control;<br>3) reporting request to other caregivers: nurses inform physician & colleagues;<br>4) participating in decision-making (within medical team): some units have customs for debating VE requests that could be followed after VE request; nurses provided information about pt to team (e.g. pt suffering); physicians made formal decision;<br>5) result of decision-making process (within the team): maximize pts comfort & supporting psychosocial needs (also for family members); possibly refer pt to other institution that performes VE; | | pt's VE request interpreted as a cry for help; lack of time to adequately care for pt with VE requests (might make pt stop or stimulate VE request); nurses felt intense moral conflict and powerlessness when pt requestes VE (for different reasons depending on their own attitude towards VE); nurses felt positive feelings due to trustful relationship with pt & ability to talk openly about Ve request; lack of time for collegial support between nurses; contextual factors (physician's treatment aims, care unit's consultative structure (including hierarchical doctor-nurse relationship, possibility of discussing VE request, the nurse's contribution to this discussion, feedback from physician after conversation with pt), time allotted for patient care, legal context, internal VE policy) affected nurses reaction to VE request; hierarchical differences between physicains and nurses may limit nurses self-efficacy; more experience with VE requests helps to react to them | | 6) aftercare: no VE was actually being carried out, but nurses have important role in supporting families after pt death | quantitative and qualitative studies aimed at refining nurses' experiences with VE; do nurses' experiences change when VE becomes legalized?; research in other Belgian regions and internationally; comparing results of this study to another country where VE is illegal | complete match |

| | | | | | | | | | | | | | | | | | | | | | | | | |
|---|---|---|---|---|---|---|---|---|---|---|---|---|---|---|---|---|---|---|---|---|---|---|---|---|
| 26 | de Boer ME et al | 2019 | Pressure in dealing with requests for euthanasia or assisted suicide. Experiences of general practitioners | Journal of Medical Ethics, 45(7):425-429 | 10.1136/medethics-2018-105120 | VE/AS | qualitative interview study | to explore the content of the pressure experienced by GPs when dealing with requests for VE/AS | physicians (=GPs) | n=15 GPs | ENG | Netherlands (VE/AS legalized) | familiy, pt, physician | | eligibility assessment (physician) | | in pressure situations: pt may threaten with suicide or question physician's integrity; pt may have voice thier already made plans faster than the physician can form his own assessment; communication with pt how to deal with counterpressure from family members | | 6 categories of pressure GPs experienced in dealing with VE/AS requests: 1) emotional blackmail, 2) control and direction by others, 3) doubts about fulfilling the criteria, 4) counterpressure by patient's relatives, 5) time pressure around referred patients, 6) organisational pressure. Pressure experienced by GPs in dealing with VE/AS requests can be attributable to factors associated with the patient-physician relationship and/or the relationship between the physician and the patient's relative(s), the inherent complexity of the decision itself and the circumstances under which they have to make the decision to (not) grant VE/AS requests or perform VE/AS. | | | further investigation of the pressure GPs face when VE/AS is requested (e.g. education of the public; education of GPs and other physicians) | complete match | |
| 27 | de Boer ME et al | 2011 | Advance directives for euthanasia in dementia: How do they affect resident care in Dutch nursing homes? Experiences of physicians and relatives | Journal of the American Geriatrics Society, 59(6):989-96 | 10.1111/j.1532-5415.2011.03414.x | VE (advance directives) | multi-methods (survey w/ physicians, qualitative interviews with selected physicians & pt relatives) | to gain insight into how advance directives for euthanasia affect resident care in Dutch nursing homes | physicians (elderly care), family members (of pt w/ dementia) | surevy: n=4834 completed, n=110 indicated having treated person w/ dementia & advance directive; interviews: n=11 physicians, n=8 family members | ENG | Netherlands (VE/AS legalized) | pts, physicians, family members | | legal requirements met (incl/ voluntary, unbearable suffering, no prospect of improvement, pt informed, shared conclusion of lack of alternatives, second physician opinion, due medical care and attention during VE)? (physcian) | discussion about VE/advance directive often initiated by other ppl than pt (because of limited capability of pt w/ advanced dementia) | discussion of adherence to advance VE directive at various, sometimes multiple timepoints; pt sometimes informed about institution's policy for advance directives | in advanced dementia, much of communication is between physicians and family members (b/c pt is no longer capable) | physicians are not legallly obligated to adhere to advance VE directives; participants had not experienced adherence to advance VE directive for pt who was no longer capable to make the decision; physicians did sometimes not adhere to advance directive even though they thought pt situation matched the situation in advance directive (mostly because communication w/ pt was not possible; but directives may have had minor effect on treatment decisions); nursing home policy may involve limited adherence or nonadherence to advance directives for euthanasia; family sometimes reluctant to VE notwithstanding advance directive; presence of advance directive may be relieveing to pt | | | | low match | changed from partial match |
| 28 | de Casterlé BD et al | 2010 | Nursing care for patients requesting euthanasia in general hospitals in Flanders, Belgium | Journal of Advanced Nursing, 66(11):2410-20 | 10.1111/j.1365-2648.2010.05401.x | VE | qualitative interview study | to explore nurses' involvement in the care process for mentally competent, terminally ill patients requesting VE in general hospitals in Flanders, Belgium | nurses | n=18 nurses | ENG | Belgium, Flanders (VE legalized since 2002) | pt, nurses, physicians, family members, psychologists, palliative support team, senior nurses; multidisciplinary team approach | complex and dynamic process w/ 7 stages: 1) period preceding the request, 2) confrontation w/ request; 3) decision-making stage (ends w/ formal decision by physician); 4) preceding the VE moment (atmosphere changes after decision is made); 5) carrying out VE (say goodbye, medical and contextual aspects, drug administration is task of physician, be present and support during VE); 6) immediate aftercare; 7) later aftercare | | pt initiate VE request; final decision by physician | stage 2: ask questions, listen, understand request; stage 3: answer question about diagnosis and prognosis, define concepts, give information | decision-making differs distinctly between multidisciplinary team settings and not (senior nurse and physician take over; nurses simply informed and sometimes feel disregarded) settings; VE request never happens unexpectedly (being open, alert, and responsive is helpful); respond with seriousness, openness, and non-judgement to requests; report request to team; nurses as pt's advocate; take sufficient time for iterative communication process; give pt sufficient time to think about VE; nurses function as mediator between pt & family; senior nurses pay attention to how team feels | nurses wish to be involved in entire process; provide palliative care in meantime; good team cooperation deemed helpful in VE process; nurses' involvement: being alert for VE request, open & active listening, multidisciplinary team cooperation & analysis of group dynamics, continuously providing maximum palliative care, multi-tasking, organizing & directing VE, support for family, colleagues & oneself | | 6) immediate aftercare (provide standard mourning care for families; pay attention to team, administrative tasks); 7) later aftercare (formal or informal follow-up w/ family, debriefing w/ team) | need for thorough ethics policies, institutional & professional guidelines; clarification of involvement of nurses w/ conscientious objection; clarification of care process after no-VE decision; gather insights into VE care process w/ non-terminally ill pt, in home care & residential elder care; evaluate VE care process after several years of experience and assess potential changes | complete match | changed from partial match; 66&72&73: same sample, differen RQs |
| 29 | de Casterlé BD et al | 2006 | Nurses' views on their involvement in euthanasia: a qualitative study in Flanders (Belgium) | Journal of Medical Ethics, 32(4):187-92 | 10.1136/jme.2005.011783 | VE | qualitative interview study | to explore in depth the views of palliative care nurses on their involvement in the entire care process surrounding VE | nurses | n=12 nurses (5 palliative care unit nurses, 4 palliative home care team nurses, 3 palliative support team nurses) | ENG | Belgium, Flanders (VE legalized since 2002; interviews carried out bevor VE was legal) | pt, nurse, physician, family members, other health care professionals; interdisciplinary approach | nurses' tasks: 1) hearing the request, 2) participating in decision-making, 3) participating in execution of VE, 4) supportin family and team | | | understand reasons for VE request; offen palliative care as alternative | nurses try to accept VE requests w/ "active openness"; some VE requests go away when taken seriously and reasonst can be alleviated with alternative options; interdisciplinary approach helpful | interviewed nurses believed to have important role in caring for pt who seek VE; nurses' involvement begins at formulation of request and ends after pt death w/ offering support to family; nurses hav longitudinal relationship w/ pt and therewith trust and important insights (e.g. regarding longstanding request vs. impulse); nurse have holsitic training; during VE, nuses are present to support pt, family members & physician; most nurses saw administering the medication as physician's task | | support for family members & team members; debriefing w/ team and psychological support | assessment of nurses' involvement in VE requests after VE legalization; do viewpoints change as VE becomes legalized?; future research in other regions and with more diverse group of participants | partial match | |
| 30 | van Bruchem-van de Scheur GG et al | 2008 | The role of nurses in euthanasia and physician-assisted suicide in the Netherlands | Journal of Medical Ethics, 34(4):254-8 | 10.1136/jme.2006.018507 | VE/AS | cross-sectional quantitative survey | to assess the role of nurses in VE and AS, as part of a study of the role of nurses in medical end-of-life decisions | nurses (w/ experiences w/ VE/AS) | n=1,179 nurses (employed in 73 hospitals, 55 home care organisations, 63 nursing homes) | ENG | Netherlands (VE/AS legalized) | pt, physician, nurse, pastor/mental attendant, social worker, psychologist, physiotherapist, social worker | | VE/AS yes or no? (physician, nurses mostly agree or sometimes disagree or have doubts w/ physician's decision) | pt initiate VE w/ physician and/or nurse | | | differences between hospitals/nursing homes & home care e.g. re: with whom VE request is raised & if interdisciplinary consultations happen; nurses less involved in VE requests in home care; sometimes lack of agreement between physician & nurse re: VE request; during VE, nurses mostly support pt and/or relatives | | | need for multidisciplinary institutional guidelines to improve collaboration between physicians & nurses and in preventing procedural misunderstandings that could lead to ethical problems and legal offences (especially hard to achieve in home care) | low match | changed from partial match |
| 31 | de Veer AJ, Francke AL, Poortvliet EP | 2008 | Nurses' involvement in end-of-life decisions | Cancer Nurs, 31(3):222-8 | 10.1097/01.NCC.0000305724.83271.f9 | VE/AS, (also other medical end-of-life decisions) | cross-sectional quantitative survey | to investigate the views of nurses on their role with regard to discussing these kinds of decisions and their actual role in the decision-making process | nurses (involved in palliative terminal care) | n=489 nurses | ENG | Netherlands (VE/AS legalized) | nurses, pts, family members, physicians | | | | | | nurses wanted to be involved in end-of-life decision-making processes; smaller number of nurses wanted to be involved in VE/AS decisions than other end-of-life decisions; 19% of nurses were involves in VE decision-making; involvement in AS decision-making was scarce; nurses talked to family members, physicians, pts about VE (more w/ family members than w/ pts)decisions; nurses need more clarity on their role; nurses' roles related to background characteristics | | | | low match | changed from partial match |
| 32 | Dees MK et al | 2013 | Perspectives of decision-making in requests for euthanasia: A qualitative research among patients, relatives and treating physicians in the Netherlands | Palliative Medicine, 27(9):27-37 | 10.1177/0269216312463259 | VE/AS | qualitative interview study | to explore the decision-making process in cases where patients request VE and understand the different themes relevant to optimise this decision-making process | pt (w/ cancer, degenerative neurological diseases, psychiatric diseases, chronic pain syndrome, heart failure, being "tired of life"), family members; physicians | N=90 interviews from 32 cases (n=31 pt; n=31 relatives; N=28 physicians) | ENG | Netherlands (VE lagalized) | pt, relatives, treating physicians | 5 consecutive phases: 1) initiation of sharing views and values about VE/AS, 2) building relationships as part of negotiating, 3) fulfilling legal requirements, 4) detailed work of preparing and performing VE/AS, 5) aftercare and closing | eligibility assessment (physician); agreeing date and time (pt & physician) | VE/AS conversations mostly initiated by pts; recommendation for shared decision-making process during VE/AS decision-making | Pts reported phase of gathering information, sharing views with relatives & questioning treating physician (most often GP) about VE/AS; Participants valued recurrent, profound, open communication, demonstrating respect for each other's views and values; Communication facilitated when physicians showed empathy, were clear about their boundaries and helped pts organize thoughts and express feelings; Participants who were satisfied about the process mentioned the sharing of information, the involvement of relatives and decisions made together; 'Unbearable suffering' is core concept that guides decision-making in VE/AS; Careful assessment whether the pt's request was voluntary, well considered and durable; After agreement to undertake VE/AS, next difficult topics are date and time of procedure and making arrangements for saying farewell | Early discussions with physicians enabled pts to prepare themselves; Early awareness of resistance to VE/AS allowed pts to consider approaching another physician; Nature & quality of the pt-physician-relationship crucial during decision-making; Effective relationships included mutual respect for autonomy, clear communication & collaboration; Pts reported barriers when they were patronized or denied freedom to voice preferences and when physicians only considered medical issues; Discontentment was reported when participants were not well-informed or physicians were either patronizing or servile; Large differences in the roles of relatives; Consultation of independent physician was found to have limited impact on course of action; Roles altered during the actual VE/AS procedure (physicians stepped into professional role and took control); Protocols only in some settings but appreciated by physicians.; Pts, relatives and physicians value rituals that accompany VE/AS. | sharing views and values about end of life and VE/AS should be initiated early in palliative care to have enough time to build relationship, prepare, plan etc. | All recruited patients declared that they had already viewed VE/AS positively before the onset of their illness. Their preparation for making a request had started at different times: before becoming ill, when diagnosed with a life-threatening problem or when facing the terminal phase of illness. Personal characteristics, such as a strong wish to have a say over one's own death, experiences of medical care and witnessing 'unbearable suffering' were cited as reasons for considering VE/AS. | 5) VE/AS places burden on physicians and patient relatives. Processing takes time. Physicians need professional support. | address the hindrances that physicians encounter in the early communication of end-of-life preferences and the support they need to deal with the requests; address the communicational skills professionals require for such complex decision-making | complete match | |
| 33 | Denier Y et al | 2009 | Involvement of nurses in the euthanasia care process in Flanders (Belgium): An exploration of two perspectives | Journal of Palliative Care, 25(4):264-74 | 10.1177/082585970902500404 | VE | qualitative interview study | to explore nurses' involvement in the care process for mentally competent, terminally ill patients requesting euthanasia in general hospitals in Flanders, Belgium. | nurses (w/ experience in VE care process) | n=18 nurses | ENG | Belgium, Flanders (VE legalized since 2002; interviews carried out bevor VE was legal) | nurses, pts, family members, physicians | 7 stages: period preceding VE request, confronting Ve requiest, decision-making, period preceding VE moment, VE moment, immediate aftercare, later aftercare; not always all stages occur; stages are not distinct/seperable | | | procedural action-focused: focus on giving neccessary information;existential-interpretative: focus on what ppl are going through and how everyone experiences the process; nurses need to provide pt with all information necessary to make the right VE decision before discussing other subjects; holistic view needed; VE decision-making communication needs constant communication partners | need to pay attention to and take care of all ppl involved in VE proccess | differentiation between procedural, action-focused and existential-interpretative perspective; well-balanced integration of the two perspectives needed; procedural, action-focused: follow guidelines/protocols, use checklist approach, respect pt request; existential-interpretative: make contact, show respect for pt as whole person, existential need for the pt's life to end in a good way, all parties involved need to make their peace and find understanding; involvement of individual nurses develops over time (procedural, action-focused perspective = learning process, existential-interpretative perspective: personal growth process) | | | what determines which perspective to be dominant?; how to integrate both perspectives?; larger quantitative samples; role of physicians | partial match | 66&72&73: same sample, differen RQs |

| | | | | | | | | | | | | | | | | | | | | | | | | |
|---|---|---|---|---|---|---|---|---|---|---|---|---|---|---|---|---|---|---|---|---|---|---|---|---|
| 34 | Denier Y et al | 2010 | "It's intense, you know." Nurses' experiences in caring for patients requesting euthanasia | Med Health Care Philos, 13(1):41-48 | 10.1007/s11019-009-9203-1 | VE | qualitative interview study | to explore nurses' experiences in caring for patients requesting euthanasia | nurses (w/ experience in VE care process) | n=18 nurses | ENG | Belgium, Flanders (VE legalized since 2002; interviews carried out bevor VE was legal) | pt, nurses, family members | | VE yes or no? (sometimes decided by physician alone, nurses want involvement & team approach) | | | legalization of VE allows for open and transparent communication about it (allows discussability); nurses experience learning curve in how to talk about VE requests (on individual and team level) | nurses decribed VE process as grave and difficult process w/ high intensity (on emotional & organizational/practical level); trying to understand the request is essential for nurses' experience (also for nurses wit conscious objection) | training needs and development of own moral stance needed from early training stages and continously | coping & self-care of nurses within the professional and private sphere | to refine nurses' experiences with euthanasia (their own role in developing critical self-reflection & the responsibilities of the hospitals in creating a context of transparency); study nurses with conscientious objection | low match | changed from partial match; 66&72&73: same sample, differen RQs |
| 35 | Descheppe r R et al | 2006 | Communication on end-of-life decisions with patients wishing to die at home: the making of a guideline for GPs in Flanders, Belgium | British Journal of General Practice, 56(522):14-9 | n/a | VE/AS, (also other medical end-of-life decisions) | guideline development w/ literature review & qualitative intervies & focus groups | to describe the development of a guideline for GPs on end-of-life communication with patients who wish to die at home, in a context where patient autonomy and euthanasia are legally regulated | GPs, pts, family members, nurses, "other caregivers", oncologists/hematologists, lawyers, ethicists | n=113 persons (17 pts, 17 family members, 25 GPs, 3 oncologists/haematologist, 8 nurses, 2 "other caregivers", 41 experts (lawyers/ethicists /oncologists)) | ENG | Belgium, Flanders (VE legalized since 2002; interviews carried out bevor VE was legal) | | clinical guideline w/ 11 steps: incl/ clarifying the patient's request, consultation with other caregivers, making formal arrangements with the patient, aspects of technical preparations (for example, choices and dosages of drugs used for euthanasia), and aftercare of next-of-kin and all the involved caregivers | | shared decision-making | aspects of GP-pt communication: truth-telling on diagnosis/prognosis/inevitability of imminient death (pt needs good understanding), exploration of pt wishes for end-of-life, disproportionate interventions for pt w/ poor prognosis, dealing w/ VE requests; thoroughly explore pt motivations, wishes, values, expectations; discuss alternatives; forgoing food and fluid administration in home care | VE requsts can be implicit or explicit; iteratively assess if pt still prefers VE; communicating technical aspects of VE can be challenging | VE requests are emotionally and practically challenging for GPs; assure continuity of care; guideline includes frequent obstacles/pitfalls (e.g. lack of open communication, ambivalence re/ VE) | | | | partial match | guideline not included in publication |
| 36 | Dobscha SK et al | 2004 | Oregon physicians' responses to requests for assisted suicide: a qualitative study | J Palliat Med, 7(3):451-61 | 10.1089/1096621041349374 | AS | qualitative interview study | to improve understanding of physicians' reactions to AS requests | physicians (diff/ specialties, from Oregon, who had received AS requests) | n=35 physicians | ENG | USA, Oregon (AS legalized since 1997) | pt, physician | | willingness to participate (physicians) | AS brought up by pts | offer all appropriate treatments | AS requests often a surprise; more experience w/ AS requests can lead to being more comfortable w/ responding to them (thought through AS requests, handle them as other end-of-life communication); physicians voiced avoidance & discomfort responding to requests; desire to offer optimal care, not abandon pts; difficult to communicate conscientious objection to pts (different approaches); lack of communication between colleagues; physicians more likely to discuss challenges within private than professional sphere | physicians voiced apprehesion re/ responding to VE requests despite general support of AS (e.g. legal/moral uncertainty, reputational concerns); AS considered a last resort or a pt's right to self-determination; worry to abandon pt when referring; phys/ w/ conscientious objection often refer to colleagues; participating in AS is intense experience/emotional investment (incl/ feelings of obligation and dedication); sometimes urgency and sense of momentum; time commitment required (incl/ house calls); formal AS process helpful for some/make sure to completely comply w/ law; not knowing pt well makes it harder; interpersonal factors shape decision-making (e.g. being determined was often admired); issues of control between pt and physician | physicians should prepare (how would I respond incl/ moral, legal & professional aspects; think through scenarios; be prepared for time & emotional commitment; find supportive ppl/forums) | some physicians want to be present for AS, others not; after AS most physician felt that they had acted appropriately/no major regrets; both discomfort and growth | | partial match | |
| 37 | Driftmier P & Shaw J | 2021 | Medical Assistance in Dying (MAiD) for Canadian Prisoners: A Case Series of Barriers to Care in Completed MAiD Deaths | Health Equity, 5(1):847-53 | 10.1089/heq.2021.0117 | MAID | qualitative interview study | to understand the process ofcare as described by physicians involved in each of the cases that resulted in MAiD | physicians | n=3 physicians (being interviewed about 3 completed MAID cases of incarcerated persons in Canada) | ENG | Canada (MAID legalized since 2015) | pts, physicians, friends, nurses | | where to take place (in prison or outside of prison?); who to be present for assessment & AS? (e.g. prision guards, police officers) ; what is allowed? (e.g. has pt to be in hand cuffs?) | | | | especially challenging due to e.g. lack of loved-ones, long waiting times, lack of access for communication; assessment partly w/in prison or in community setting | | | needs of prisoners re/ MAID; understand how incarcerated end-of-life care pt experience denail of release/MAID; how to offer MAID for incarcerated pts | low match | changed from partial match |
| 38 | Emanuel EJ e | 1998 | The practice of euthanasia and physician-assisted suicide in the United States - Adherence to proposed safeguards and effects on physicians | JAMA, 280(6):507-13 | 10.1001/jama.280.6.507 | VE/AS | longitudinal qualitative interview study | to determine whether the practices of VE and AS are consistent with proposed safeguards and the effect on physicians of having performed VE or AS | oncologists (w/ experience in VE/AS) | n=38 oncologists w/ experience w/ Ve and/or AS (out of 355 oncologists who started an interview) | ENG | USA (VE/As illegal when interviews were conducted) | | | | VE/AS mostly initiated by pts, seldom by family members or physician | pt often possessed medication and asked for how to use it for AS | physicians sometimes discussed case w/ colleague; family members were sometimes involved/supportive, sometimes not involved, seldom opposing | havin long term relationship w/ pt critical for willingnes to be involved in AS/VE; VE/AS experience shaped physisians' openness, alertness, empathy for future pts (some more open, some less open) | lack of correct understanding/knowledge of concepts AS/VE | some regret involvement in VE/AS; some fear prosecution | | low match | changed from partial match |
| 39 | Evenblij K et al | 2019 | Euthanasia and physician-assisted suicide in patients suffering from psychiatric disorders: a cross-sectional study exploring the experiences of Dutch psychiatrists | BMC Psychiatry, 19:1-10 | 10.1186/s12888-019-2053-3 | VE/AS | cross-sectional quantitative survey | to estimate the incidence of VE/AS (requests) in psychiatric practice in The Netherlands and to describe the characteristics of psychiatric patients requesting VE/AS, the decision-making process and outcomes of these requests | psychiatrists | n=207 psychiatristis | ENG | Netherlands (VE lagalized) | relatives, pts, physicians, other psychiatrist, SCEN-physician ("support and consultation on euthanasia in the Netherlands"), medical specialist | | | VE/AS initiated by pt | | | VE/AS requests in psychiatric pts increasing but relatively rare; many pts treated by physician for less than a months before requesting VE/AS; decision-making processes took between 2 months and 2,5 years before granting, 0 months and 1 year before rejecting VE/AS; refusal of VE/AS request mostly because at least one of due care criteria was not met (different reasons for refusal reported); after refusal sometimes referral to other institutions/providers; family members sometimes not involved, supportive, neutral, divided, or opposing; other physicians sometimes consulted | | | | low match | changed from partial match |
| 40 | Gamondi C et al | 2019 | Responses to assisted suicide requests: an interview study with Swiss palliative care physicians | BMJ Supportive & Palliative Care, 9(1):1-9 | 10.1136/bmjspcare-2016-001291 | AS | qualitative interview study | to describe palliative care physician's perspectives and involvement in assisted suicide practices | palliative care physicians | n=23 physicians | ENG | Switzerland (AS legalized) | | pt request; assume responsibility toinvestigate request; iterative circle of: explaining limits in participation, exploring pt's reasons to seek AS, present/discuss alternatives & risks of AS, include families in discussion | | AS communication initiated by pts (participants mostly would not initiate AS communication) | setting boundaries early on; playing active role in AS decision-making (incl/ exploring reasons, offering alternatives, informing about risks; often iteratively); engaging w/ family about AS; provision of eligibility certificate; interaction w/ right-to-die organizations; physician's unwillingness to prescribe; being present at death | be open for right-to-die organizations to communicate/visit w/ pt; no communication w/ other providers outside of institution (despite desire for better collaboration w/ right-to-die organizations) | AS requests often interpreted as requests for information rather than actual AS; fear of being stigmatized as physician; need to more clearly define roles and responsibilities | a third each saw AS as actually/potentially a tool in palliative care, no tool, or were undecided; individual palliative care physician's willingnes for involvement/prescribing determined by their personal attitudes towards AS (some conscientiously objected, ethical dilemmas when personal and institutional stance are in conflict); learning by doing w/ guidelines or senior role models | | further explore domains to address inc ommunication about AS requests; further explore training/education needs; further explore personal objections | partial match | |
| 41 | Gamondi C et al | 2015 | Exploring the experiences of bereaved families involved in assisted suicide in Southern Switzerland: a qualitative study | BMJ Supportive & Palliative Care, 5(2):146-52 | 10.1136/bmjspcare-2013-000483 | AS | qualitative interview study | to explore family involvement in decision making prior to AS, and to examine their ways of coping during the bereavement period | family members (of pt deceased of AS) | n=11 family members (of 8 pts) | ENG | Switzerland (AS legalized) | pts, physicians, relatives | | | | | | process of the AS preparation: 1. pts' reasons for AS, 2. moral dilemmas, 3. type of family involvement, 4. negotiations in decision-making phase; modalities of AS realisation: 1. when, 2. perception of type of death, 3. rituals, 4. interaction with police authorities ; secrecy management during decision-making process (some pts/relatives choose carefully whom to talk to, others very open); two types of involvement of relatives during decision-making: active and passive (sometimes different for different family members); feeling of isolation of families; worries about potential stigma of AS | | coping and bereavement: secrecy management, isolation, moral dilemmas | proper recognition and assessment of famiyl members' experience furing AS process; assess impact of future guidelines including recommendations for family members | partial match | |
| 42 | Gamondi C, Pott M & Payne S | 2013 | Families' experiences with patients who died after assisted suicide: a retrospective interview study in southern Switzerland | Annals of Oncology, 24(6):1639-1644 | 10.1093/annonc/mdt033 | AS | qualitative interview study | to explore patients' reasons for choice of AS and family perceptions of the interactions with HCPs | family members (of patients w/ cancer, cared for by a palliative care team & deceased of AS) | n=11 family members (of n=8 diseased pt) | ENG | Switzerland (AS legalized) | pt, relatives, right-to-die organization, GP, oncologist, palliative care physician, district nurses, palliative care nurses some pt did not want to discuss AS with physicians; most pt were reported to be uniterested in involving palliative care professionals; in 3 cases, pt/relative found way to get documents needed without involvement of their physicians | At some point during decision-making process, some relatives recalled an urgency to undertake AS before physical condition of pt deteriorated too much (rendering them unable to actively take the medication) or a natural death could occur. | | | in some cases, pts threatened with suicide to put pressure on relatives opposing AS | | some professionals lacked awareness of & were unsupportive of the pt's existential suffering; family members' needs should be conidered in addition to those of the pt | long-standing personal beliefs that patients held in favour of AS if these circumstances arose (sometimes long before cancer diagnosis); sometimes transgenerational acceptance of AS within family; pts had fear of the future, fear of loss of dignity, a lack of independence in daily activities and bodily functions; pt had feelings of anger, impotence, desperation and hopelessness; diagnosis of life-threatening illness as trigger to act | | clinical guidelines for AS requests are needed | complete match | |

| | | | | | | | | | | | | | | | | | | | | | | | | |
|---|---|---|---|---|---|---|---|---|---|---|---|---|---|---|---|---|---|---|---|---|---|---|---|---|
| 43 | Gamondi C et al | 2018 | Family Caregivers' Reflections on Experiences of Assisted Suicide in Switzerland: A Qualitative Interview Study | Journal of Pain and Symptom Management, 55(4):1085-1094 | 10.1016/j.jpainsymman.2017.12.482 | AS | qualitative interview study | to explore family caregivers' reflections on experiences of assisted suicide in Switzerland | family members (of pt deceased of AS) | n=28 family members (of 18 deceased pt) | ENG | Switzerland (AS legalized) | pt, family member, physician various forms of involvement of family members | 1) contemplation; 2) gaining acceptance; 3) gaining permission; 4) organization; 5) aftermath<br>AS appeared to be the result of numerous negotiations happening within the family circle and outside. | eligibility assessment (right-to-die organization); choice of the day (pt & right-to-die organization); form of family member involvement (family members) | initiation of contact with right-to-die organization mostly jointly by pt & family member | 2) After the diagnosis of life-threatening disease, pt expressed fears to family members about indignity, loss of control, & loss of independence and began to discuss AS intentions with selected family members. Pt seeking for allies in family and HCPs. Acceptance of AS appeared to be discussed initially only within the family, marginally involving HCPs. | 3) family members help search for physicians to provide certificates & prescription; family members act as advocates; family members went with pt to initially contact right-to-die organizations; family members had to overcome dilemmas concerning their involvement in AS; 4) practical organization of AS: eligibility assessment with right-to-die organizations; setting a date; planning funeral, rituals, people present during AS | Family members are involved in AS in various forms; family members needs need to be addressed; pt and families discussed in depth before coming to an agreement | 1) pt contemplated assisted suicide for a long time (sometimes long before illness); reasons for seeking AS have been established over a lifetime (fears for the future, actual loss of meaning in life, fears of suffering & decay, perception of medical care as aggressive, inhumane, and invasive | 5) respecting the pt decision for AS and suffering; intense process; disclosure was careful and partial, depending on the participants' capacity for anticipating or overcoming actual or anticipated stigma; none of participants received nor asked for professional support during bereavement | pay greater attention to family involvement in decision making and organization of AS, whether obtained in the patient's own country or abroad. | complete match | 92&93: same sample, different RQs |
| 44 | Gamondi C et al | 2020 | Swiss Families' Experiences of Interactions with Providers during Assisted Suicide: A Secondary Data Analysis of an Interview Study | Journal of Palliative Medicine, 23(4):506-12 | 10.1089/jpm.2019.0286 | AS | qualitative interview study | to explore how Swiss families interact with HCPs and right-to-die associations regarding AS and their choices around disclosure | family members (of pt deceased of AS) | n=28 relatives (of 18 deceased pt) | ENG | Switzerland (AS legalized) | familiy, pt, physician, other physician, close friends, right-to-die associations, HCPs, palliative care staff | | | pts or family members initiate AS process | | families mostly avoided discussing AS considerations beyond the neccessary w/ HCPs (e.g. due to expected conflicting values); some participants reported helpful open communication w/ HCPs; HCPs often wanted to discuss reasons for AS request, but not help moving request along; referral to psychologists/chaplains not appreciated | AS decision was not perceived as a medical, buta personal decision; families need to be recognized as closely involved in AS processes & need to be aware of their needs; expectation that physician would provide neccessary certificates/prescription (if not, other physician was sought); once neccessary certificates/prescription were obtained, mostly interaction w/ right-to-die association (as competent and trustworthy partner); some family members felt AS and palliative care to be mutually exclusive; reasons for AS related to personal values | | some participants reported sense of isolation after pt's death | families' role should be investigated in all countries that legalized AS | partial match | 92&93: same sample, different RQs; inconsistenciy in reporting of sample in the publication |
| 45 | Gastmans C, Van Neste F & Schotsmans P | 2004 | Facing requests for euthanasia: a clinical practice guideline | Journal of Medical Ethics, 30(2):212-217 | 10.1136/jme.2003.005082 | VE | guideline development by consensus meeting (opinion based) method | to make explicit how medical and nursing expertise could effectively be employed in the interdisciplinary care context, so that the competent, terminally ill patient who requests euthanasia receives the best human care available | experts | | ENG | Belgium (VE legalized) | pt, physician, second physician, friends and family, relatives, nurses, healthcare institution, palliative support team | 1) palliative care for all; 2) when VE is requested (often initially to nurses): attentive listening to VE request, need for open dicussion, "palliative filter function", involve palliative care team to consult; 3) if VE request persists: accept request, assess legal criteria, consult a second independent physician (knoowledgeable about determination of requets is voluntary, well-considered, repeted & palliative care), 4 steps: consultation w/ first physician + review medical file + examine patient and their request + write report; 4) making a decision: primary physician makes decision, but has to be based on close consultation w/ nurses, healthcare team, patient, relatives; 5) primary physician writes report to Federal Control and Evaluation Committee and informes authorities | approval of VE request yes or no (primary physician) | process of interpersonal agreement (patient, relatives, physician, nurses, and palliative support team who enter into a dialogue) | Inform patient about their medical situation; explore palliative care alternatives and other alternatives (with support of palliative care support team); ensure that VE request is based on autonomous, free, informed choice (inquire about appropriate interpretation of the request, motivation, sufficient information?, current menatl competency, other people involved in considerations, voluntariness, no coercion/pressure) | caregivers should listen, show openness for dicussing VE and pt's view; give pt opportunity to consult with palliative care team; physician will communicate with relatives about VE request at pt's reqeust; phyisican will disucss VE request with nursing team (e.g., give option for conscientious objection) | VE requests need to be understood, specified, and interpreted in a shared reflection process to go from vague requests to informed decisions; good cooperation between physicians and nurses is pivotal | | | evaluate Caritas Flanders' clinical practice guideline's success in building bridges between ethics, law, and clinical practice; evaluate the use of clinical practice guidelines for the care of patients requestin VE; implementation research regarding guideline use for VE requests | partial match | changed from complete match |
| 46 | Goh E, Wen J &Yu CE | 2021 | Lessons from the Departed: A Planned Behavior Approach to Understand Travelers' Actual Physician-Assisted Suicide Behavior | Journal of Hospitality & Tourism Research, 20(10):1-15 | 10.1177/1096348020987626 | AS | qualitative interview study (using interviews from news channel documentaries) | to examine the underlying motivations of actual individual travelers who have travelled to Switzerland as their final life destination to commit physician-assisted suicide; including reasons, important people who influence an individual, difficulties, anticipated guilt | pts ("travelers" (travelling to Switzerland to end their life there through AS)) | n=26 pts/"travelers" | ENG | Switzerland (AS legalized also for "travelers") | family members, pts, relatives, friends | | | | | | lack of access to AS, high financial cost, lack of knowledge, faith as a perceived difficulties; family members & friends as most important groups of ppl involved in AS process (some supportinve, some opposed) | pts reasons to consider AS: end suffering, die w/ dignity/on own terms, end poor quality of life, basic human right | | explore perspectives of other stakeholders involved in the AS process to develop a more complete picture of AS; quantitatively test the elicited items with a larger sample of individuals eligible for AS to better understand their AS intentions and behavior; develop theoretical frameworks to understand and support the decision-making process of AS travelers, such as information searches and selection of a AS facility, in order to provide better support | low match | |
| 47 | Groenewoud J H | 2004 | Psychiatric consultation with regard to requests for euthanasia or physician-assisted suicide | General Hospital Psychiatry, 26(4):323-330 | 10.1016/j.genhosppsych.2004.02.004 | VE/AS | cross-sectional quantitative survey | to describe the practice of psychiatric consultation with regard to explicit requests for euthanasia or physician-assisted suicide in the Netherlands | psychiatrists | n=549 psychiatrists | ENG | Netherlands (AS/VE legalized since 2002) | psychiatrist, pt, physician | physicians (nonpsychiatrists or psychiatrists) consult psychiatrist for further evaluation (happens rarely) | Is the request well-considered? (psychiatrist); Is a mental disorder present? (psychiatrist); Was the decision influenced by (counter)transference? (psychiatrist) | | | | pt in the Netherlands can request VE/AS due to physicial or mental disorder or both; 30% of VE/AS requests referred to a psychiatrist resulted in VE/AS; vast majority of cases where VE/AS was acceeded for non-psychiatrists' pt; requests due to physical disorder more likely to result in VE/AS than requests due to mental disorder; older pt more likely to be granted VE/AS; no sex difference; cases in which the psychiatrist had examined pt personally more likely led to VE/AS; requests that were acceeded and those that were not differed in several aspects; psychiatrists were most frequently asked to assess possible mental disorders and well-consideredness of request | pt requesting VE/AS due to mental disorder were younger and more likely female than pt requesting due to physcial disorder; most frequently reported motives were pt hopelessness or unbearable suffering | | What circumstances require psychiatric consultation? What is the potential of psychiatrists in exploring pt's request for VE/AS? Implications of psychiatrist's judgment of pt's competence, presence of mental disorder, etc? Prevalence of mental disorder among pt requesting VE/AS? Other aspects that may influence pt's competence? | complete match | |
| 48 | Hagens M et al | 2021 | Experiences with counselling to people who wish to be able to self-determine the timing and manner of one's own end of life: a qualitative in-depth interview study | Journal of Medical Ethics, 47(1):39-46 | 10.1136/medethics-2019-105564 | AS | qualitative interview study | to explores the experiences with counselling that counselees receive(d) from counsellors facilitated by Foundation De Einder (offers counselling to people who wish to be able to self-determine the timing and manner of their end of life int he Netherlands). | counselees of Foundation De Einder | n=17 counselees (in 14 interviews, 3 interviews with couples) | ENG | Netherlands (AS/VE legalized since 2002) | counsellors, physician, pt, family&friends | | | | In context of counselling: screening of reason to seek counselling; information about different ways to end one's life; knowing how to obtain medication; when to end one's life; preparation for situation after death; psychological aspects of the process; moral and philosophical aspects of ending one's life and meaning of life; effects on family and friends | great variety between 1 and 24 counselling sessions; possibility to involve other ppl in counselling; counselling entails more than just giving information on medication | having means to end one's life can be reassuring and enhance quality of life; new worries about the medication may arise; in general positive experiences of counselling (trustworthy, careful, open, non-judgemental, good listeners, available, no coercion); also criticism about e.g. feeling dependent on counsellor, secrecy, competence of counsellor; physicians should be more open to counselling and appreciate role of counsellors; counselees reported having gotten medication from internet or black market; counselees did not worry about impulsivity | | | | complete match | |
| 49 | Hamilton NG & Hamilton CA | 2005 | Competing paradigms of response to assisted suicide requests in Oregon | Am J Psychiatry, 162(6):1060-5 | 10.1176/appi.ajp.162.6.1060 | AS | case report | to compare the traditional approach to evaluating and treating suicidal symptoms with the assisted suicide competency model delineated in an assisted suicide guidebook used in Oregon | pt w/ lung cancer & psychiatric history | n=1 pt | ENG | USA, Oregon (AS legalized since 1997) | pt, family members, primary care doctor, friends, nurse, psychiatrist, emergency room physician, psychiatric social worker, mental health technician, social worker, palliative care consultation, Physicians for Compassionate Care volunteer, assisted suicide activist physician | | determine decision-making competency (physician), do depression/other mental illness affect decisions about AS? (psychiatrist) | AS initiated by pt | conversation w/ Physicians for Compassionate Care volunteer: collect information on diagnosis, prognosis, treatment options; evaluate emotional processing of pt; assess family support; offer support e.g. to find providers to help w/ depression/pain | | difficult to mix traditional clinical model for evaluating and treating suicidal ideation and assisted suicide competency model (dilemma) and may prove confusing to clinicians & pts, contribute to substandard medical care, and endanger seriously ill pts, particularly those with a history of preexisting mental illness | pt w/ psychiatric history (several suicide attempts, alcohol use disorder (sober)) | | | low match | |
| 50 | Handley MA | 2020 | The "Foresty Way": My Mother's Brave Choice of Medical Aid in Dying | Annals of Family Medicine, 18(6):553-4 | 10.1370/afm.2606 | MAID | case report (written by family member) | n/a | pt w/ atypical presentation of lymphoma | n=1 pt | ENG | USA, California (AS legalized since 2016) | palliative care physician, daughter, pt, oncologist, physician from AS group | | how, when, where, with whom (pt & daughter) | AS discussion initiated by pt | pain; wishes pt has in his/her life; ways hospice might help; treatment options; prognosis; how the AS process works | | | prior treatments | | | low match | changed from partial match |
| 51 | Ho A et al | 2021 | How does Medical Assistance in Dying affect end-of-life care planning discussions? Experiences of Canadian multidisciplinary palliative care providers | J Palliat Med, 24(2):189-194 | 10.1177/26323524211045996 | MAID | qualitative interview study | to explore how the Canadian Medical Assistance in Dying legislation affects end-of-life care discussions between patients and multidisciplinary specialist palliative care providers. | specialist palliative care providers from Vancouver and Toronto including physicians, nurses, social workers, and allied health professionals (AHPs) from different settings | n=48 specialist palliative care providers (26 from Vancouver, 22 from Toronto; 22 physicians, 15 nurses, 7 social workers, 4 allied health professionals) | ENG | Canada (MAID legalized) | pt, family members, physician, nurses | | differentiate between requests for palliative care and MAID requests (clinicians) | hesitation to initiate MAID topic as palliative care provider | conversation around procedural matters is complex; discussions about suffering and quality of life (including existencial and psychosocial suffering); inform about alternative options | concerns regarding proactive discussion of MAID; challenges of promoting open-ended discussions; need for iterative encounters; how to manage pt-family-clinican tryad; uncertainties regarding wish-to-die statements; should there be comprehensive exploration of all end-of-life care option before discussing MAID?; availability of MAID may change image of palliative care and how palliative care conversations unfold (MAID as conversation stopper) | worry about potentially subjective interpretations of eligibility criteria and the implementation pathway for MAID | | | whether or how may the gradual normalization and potential expansion of MAID affect the roles and practices of ghealth care providers; how can we improve professional curricula to enhance health care providers' ability to engage in open and compassionate end-of-life care discussions | complete match | |
| 52 | Hopkins HJ & Hopkins KD | 2017 | When choosing death is an affirmation of life: A public remembrance of our family's journey | Review & Expositor, 114(3):414-423 | 10.1177/0034637317722150 | AS | case report | to theologically reflect as family members' of a person who died by AS | pt; daughter and son-in-law (authors) | n=1 pt | ENG | USA, California (As legalized) | pt, family members, friends, physician, hospice nurse, end of life option program respresentatives | pt must make two verbal and one written requests; consultation with second physician necessary; | decision to seek and go through with MAID (pt); are prerequisits met (physician); when to take medication (patient); who should/wants to be present for MAID (pt, family) | | End of Life Option Program respresentatives explain process and requirements; conference calls between pt and family about the why of MAID request, financial situation, car, plans for memorial service, what to do with ashes after being cremated, quality of life; ask family members who wants to be present for MAID | request must be discussed between pt and physician without anyone else being present (to safeguard agains coercion) | MAID to maintain control and command in face of pain and uncertainty; practical aspects (e.g. medication cumbersome to administer (90 capsules that have to be opened individually) and expensive (more than 2,000.00 USD not covered by insurance)) | pt had had surgery and chemotherapy for cancer; pt had experience of being hospitalized and in nursing facility | MAID brought family together; being at peace with pt's decision to utilice MAID; questions, regrets, and uncertainties of family members after MAID | MAID and dementia?; access to MAID?; affordability of MAID? | complete match | |

| | | | | | | | | | | | | | | | | | | | | | | | | |
|---|---|---|---|---|---|---|---|---|---|---|---|---|---|---|---|---|---|---|---|---|---|---|---|---|
| 53 | Inghelbrecht E et al | 2010 | The role of nurses in physician-assisted deaths in Belgium | Canadian Medical Association Journal, 182(9):905-910 | 10.1503/cmaj.091881 | VE, (also life-ending medication without explicit request) | cross-sectional quantitative survey | to investigat the involvement of nurses in the decision-making and in the preparation and administration of life-ending drugs with a patient's explicit request (VE) or without an explicit request; to examine factors associated with these deaths | nurses involved in cases of assisted death | n=248 nurses (128 VE, 120 without explicit request) | ENG | Belgium (VE legalized) | nurse, pt, physician | | | deciding together between physcian and nurse in about 1/4 of cases | | | nurses reported VE as well as administration of life-ending drug without explicit pt request; around 2/3 of nurses of pt who received VE reported involvement in decision-making (with varying experiences, dependent on setting and nurse's age); involvement of nurses in physican's decision-making mostly exchange of information about pt's or relatives' wishes and about pt's condition; nurses were sometimes present during VE and offered support to pt, relatives, physician, other nurses; sometimes physician not present during administration of VE drug by nurse (not legal) | | | closely monitor and examine the involvement of nurses in these practices nationally and internationally to allow comparisons between countries with and without VE legislation | partial match | changed from complete match |
| 54 | Jansen-van der Weide MC et al | 2009 | How patients and relatives experience a visit from a consulting physician in the euthanasia procedure: a study among relatives and physicians | Death Studies, 33(3):199-219 | 10.1080/07481180802672272 | VE/AS | multi-methods (secondary analysis of quantitative and qualitative analysis of relatives' interview and GPs' questionnaire data from two studies) | to investigate the impact of a visit from a consulting physician on the patient and the relatives during the euthanasia procedure in The Netherlands; to explore the expectations and experiences of patients and relatives with regard to the consultant's visit | pt relatives; GPs | n=86 relatives (from one of the two included studies), n= 1,421 GPs (from the other study) | ENG | Netherlands (AS/VE legalized since 2002) | GPs, pt, relatives, consulting physcician | formal consultation by 2nd physician (incl/ discussion btw/ consultant and physician, reading pt's file, and talking to pt (as required)) | | | | | some pts are reluctant to see consultant (e.g. bc/ of fear of negative advice); reasons for consulting physicians given by GPs were legal requirements, confirmation of prudent practice, independent judgement; GPs had often promised approval of VE/As request prior to consultation w/ second physician; some pts have negative experiences w/ consultants; to maximize positive experiences, attention could be paid to possible tiring effect of visit, consultant's attitude, clear decision-making by attending physician, introduction of consultation that focuses on safeguarding of the physician's practice | | | | low match | |
| 55 | Jansen-van der Weide MC et al | 2005 | Granted, undecided, withdrawn, and refused requests for euthanasia and physician-assisted suicide | Archives of Internal Med, 165(15):1698-704 | 10.1001/archinte.165.15.1698 | VE/AS | cross-sectional quantitative study (secondary analysis) | to obtain information about the characteristics of requests for VE/AS and to distinguish among different types of situations that can arise between the request and the physician's decision | GPs | n= 3,614 GPs | ENG | Netherlands (AS/VE legalized since 2002) | physician, 2nd consulting physician, pt | formal consultation by 2nd physician | | VE/AS reuqests initiated by pt | VE/AS requests mostly completely/highly explicit | | VE/As requests can be granted, refused, pt might bie before VE/AS performance or before final decision, pt might withdraw request; GPs sometimes already certain about granting request before initiating formal consultation by 2nd physician; physicians reported uncertainity re/ granting/refusing of VE/AS requests; most frequently reported reasons for requesting VE/AS: pointless suffering, loss of dignity, and weakness | | | | low match | changed from partial match |
| 56 | Jeba J | 2019 | Did I Hear You Right? | Journal of Pain & Palliative Care Pharmacotherapy, 3381-2):59-61 | 10.1080/15360288.2019.1620401 | VE | case report (personal experience of a palliative care physician) | to provide an example of a unique experience, w/ early requests for euthanasia that transitioned to aggressive end of life care later | 1 pt case described by a palliative care physician | n=1 pt | ENG | India (VE/AS not allowed) | physician, pt, family member | | | | explore underlying reasons for VE/As request; consider alternatives such as symptom control, support by chaplain or clinical psychologist; give comprehensible information against lack of information and unrealistic fear | VE/AS requests signpost unbearable suffering; VE/AS requasts should be dealt with respect and empathy; use good communication skills; empower pt and family to regain sense of control | requests for VE/AS might be changing over time and seem conflicting w/ other requests (e.g. for more treatment) | | | | low match | changed from partial match |
| 57 | Kompanje EJO, de Beaufort ID & Bakker J | 2007 | Euthanasia in intensive care: A 56-year-old man with a pontine hemorrhage resulting in a locked-in syndrome | Critical Care Medicine, 35(10):2428-2430 | 10.1097/01.Ccm.0000281854.12364.F5 | VE | case report | to describe a case of VE in intensive care of a 56-year-old man with a pontine hemorrhage resulting in a locked-in syndrome (could communicate by eye-opening) | pt w/ pontine hemorrhage resulting in locked-in syndrome | n= 1 pt | ENG | Netherlands (AS/VE legalized since 2002) | physician, second intensivist, third physician, relatives, pt, nurses | request by relatives; checking prerequisits by physicians & interviewing the pt by eye-opening; physician informed the hospital board and nursing staff; administering the drug & death of pt; informing the autorities about VE; regional review committee reviews case; pt relatives visited hospital after death of pt and voiced their contentment | are prerequisits for VE met? (3 physicians) | | information about prognosis; Does pt wish prolonged treatment?; communication of options withdrawal of ventilation & fluids or VE; several re-checks if pt wished VE (always answered with yes) | pt could communicate by eye opening; primary care physician was not informed upon relatives' request | | described as intelligent, independent, active, and proponent of VE by relatives; no written advance directive for life-sustaining treatment; locked-in syndrome after hemmorhage | two weeks after VE, relatives visited hospital and expressed their thankfulness | | complete match | |
| 58 | Kouwenhoven PSC et al | 2015 | Opinions about euthanasia and advanced dementia: a qualitative study among Dutch physicians and members of the general public | BMC Medical Ethics, 16(6):1-6 | 10.1186/1472-6939-16-7 | VE | qualitative interview study | to investigated the differing opinions of physicians and members of the general public on the acceptability of euthanasia in patients with advanced dementia | physicians (including medical specialists, general practitioners, elderly physicians); members of the general public | n=67 persons (16 medical specialists, 19 general practitioners, 16 elderly physicians, 16 members of the general public) | ENG | Netherlands (AS/VE legalized since 2002; can be performed according to written advance euthanasia directive (AED) when a patient is incompetent as long as all legal criteria of due care are met) | | | Can criteria of due care be met in pt w/ advanced dementia? (physician) | | explicit confirmation of wish for VE and unbearable suffering at time of request | | meeting the criteria of due care for VE in late stage dementia seems to be hard to achieve; is unbearable suffering forseeable?; is suffering in dementia unbearable to pt or their relatives?; while advance euthanasia directives are legally possible on NL, they seem to often be insufficient tp physicians making the decision about VE | | | | complete match | |
| 59 | Lehto RH et al | 2016 | When a patient discusses assisted dying: nursing practice implications | Journal of Hospice & Palliative Nursing, 18(3):184-191 | 10.1097/njh.0000000000000246 | AS | case report | to describe practical strategies and resources for palliative care and hospice nurses who may have dialogue with patients relative to end-of-life concerns | pt w/ cancer | n=1 pt | ENG | USA, Oregon (AS legalized since 1997) | nurse, pt, family members, oncology mental health provider, health-care team (in interdisciplinary family meetings), hospital ethics committee | | move to Oregon to have option of AS (pt), decision about further treatments (pt) | | How long has pt been considering AS?; Is pt of "sound mind"?; where has pt gone for information?; are treatable physical symptons such as pain or dyspnea contributin to wish to die?; spiritual and cultural beliefs; reseources; explore other complementary treatments such as yoga, massages etc.; clarifying available options with knowledge of pt values and expectations; explore psychological symptoms; inform pt what information from confidential discussion will be shared and what will not | nurses are instrumental in building relationships w/ pt and initiating discussions relative to problematic concerns; allow pt to openly share and be non-judgemental; facilitate discussion w/ other care providers (e.g. mental helath providers, clinical ethics consultants); facilitate family meetings | decision agains further treatment or for AS might complicate relationship with relatives; provide high-quality education materials (such as advance directives, managament of finances and personal resources, cancer internet resources) and other resources (support groups, advocacy organizations); nurses should document when discussing end-of-life w/ pt; nurses should understand their own position about end-of-life care and know resources | Pt reported having informed herself through media about MAID | Pt did not follow through with moving to Oregon to have option of AS | | partial match | changed from complete match |
| 60 | Lemiengre J et al | 2008 | How do hospitals deal with euthanasia requests in Flanders (Belgium)? A content analysis of policy documents | Patient Education and Counseling, 71(2):293-301 | 10.1016/j.pec.2007.12.010 | VE | document analysis | to describe the form and content of ethics policies on euthanasia in Flemish hospitals and the possible influence of religious affiliation on policy content. | policy documents from Flanders (Belgium) | n=42 policy documents | ENG | Belgium (VE legalized) | physicians, palliative support team, nurses, pt, relatives, interdisciplinary teams for euthanasia decision-making (incl. nurses, physicians, palliative experts, social workers) | 1) confrontation with VE request; 2) decision-making process; 3a) care process if decision agains VE (incl. inform about reasons, discuss alternatives, social/psychosocial/spiritual care); 3b) preparation and performance of euthanasia if decision for VE (incl. care plan, medication protocol; only physicians can administer medication) ; 4) aftercare | who is to be informed about VE request (pt); whether to attend act of VE (relatives) | | react to VE request by listening (primarily physcian in charge and nurses); informing about symptom management options (primarily palliative support team); informing about hospitals VE protocol; informing about act of VE; offering social/psychosocial/spiritual care; pt should participate in formulation of VE care plan | guidelines can inform who should repond how when confronted with VE request; in addition to legal procedures, primary physician should consult palliative support team, nurses, relatives, GPs; formation of interdisciplinary team to discuss VE request (incl. primary physician, palliative support team, nurses, GPs, pastors/moral consultants, social workers; tasks: giving advice, checking legal requirements, forming VE care plan, clarifying stances & venting emotions, considering alternative options; 1 or more meetings, morderated by primary physician or senior nurse); relatives should also be involved and informed; professional attitudes should be e.g. empathetic, serene, respectful, caring, trustworthy; communication skills are needed | policies described hospital's stance on VE, VE care process, involvement of clinicians/pt/relatives, ethical aspects, other issues (e.g. support for clinicians (e.g. through palliative support team, ethics team, attending), reporting, professional attitudes, communication skills); reporting: documentation of discussions had and legally required documents; VE care plan should include role of relatives; clinicians have right to have conscious objections (guidelines inform what to do then) | | aftercare for deceased pt; aftercare for relatives (systematically planned vs. provided upon request; one or several conversations); aftercare for clinicians (systematically planned vs. upon request; debriefing; possibly w/ external counselor); administrative aftercare (documentation, legal forms, reportinng to ethics committeet, reporting to attendants) | How are quidelines implemented?; Do guidelines empower/support participants? | complete match | |
| 61 | Li M & Kain D | 2018 | The other side of sorrow: physician reflections on assisted dying | CMAJ, 190(6):E169-E170 | 10.1503/cmaj.171114 | MAID | case report | n/a | 2 pt cases (cancer, bowel obstruction), described from perspectives of palliative care physician & cancer psychiatrist | n=2 pt | ENG | Canada (MAID legalized since 2015) | pt, palliative care physician, oncology psychiatrist, relatives, intervention physician (administering MAID meds) | | initiate/request for MAID (pt); eligibility (physican); changing one's mind about MAID (pt) | | personal attributes and legacy of the person requesting MAID; reasons for MAID; alternatives; date and time | | | | | | partial match | changed from complete match |

| | | | | | | | | | | | | | | | | | | | | | | | | |
|---|---|---|---|---|---|---|---|---|---|---|---|---|---|---|---|---|---|---|---|---|---|---|---|---|
| 62 | Li M et al | 2017 | Medical assistance in dying - implementing a hospital-based program in Canada | NEJM, 376(21):2082-2088 | 10.1056/NEJMms1700606 | MAID | implementation report (i.e., process description) | n/a | University Health Network (UHN) in Toronto including 4 tertiary care teaching hospitals | n/a | ENG | Canada (MAID legalized since 2015) | pt; clinical team (nurses, attending/most responsible physician (MRP), allied health professionals, consultant physicians); assessment team (all have volunteered to participate; two physicians skilled in assessing patients' prognosis, suffering, and capacity to provide informed consent, if disagreement then involvement of MAID oprogram leaders); intervention team (all have volunteered to participate; physician or nurse practitioner | MAID framework: 1) clinical team (pt requests MAID, notify MRP, MRP to further discuss, refer to palliative care/psychiatry/social work as required, pt's MAID decision, if yes: MRP complete MAID request and contact assessment team), 2) MAID assessment team (is pt eleigible?, if no, pt may seek MAID outside elsewhere, if yes: pt MAID request form, mandatory reflection period), 3) intervention team (intervention physician/nurse review all documents, meet w/ pt, complete MAID consent form, re-assessment of pt decision, discuss timing and setting, complete prescription, deliver MAID) | initiate MAID (pt); refer to palliative care/psychiatry/social work for additional support? (MRP, pt); initial MAID decision (pt); copacity for informed consent? (different physicians at different steps); pt eligible? (assessment team); acceptance of request after review of documents and pt encounter (intervention team); repeat MAID decision (pt); timing and setting (pt, intervention team) | initiation of the topic by pt; different responsibilities for different steps withing the process | diagnosis & prognosis; adequacy of symtom management & options for further management; reasons for request; eligibility for MAID; hospital's process and reflection period; timing and setting for MAID; mandatory referral to provincial organ donation | three team model (clinical team, assessment team, intervantion team; to respect conscientious objection/personal discomfort + requirement for reasonable knowledge, care and skill); reasons for not proceeding to assessment after request: withdrawal of request by pt, primary mental illness, delirium, active dying; 86% of assessed pt were approved for MAID (reasons for non-approval: lack of capacity to provide informed consent, indecision about receiving MAID at the time of assessment); 76% of pt granted approval received MAID (reasons for not proceeding: natural death, change in the pt's decision) | early experience: many patients who request MAiD do not receive it because death or loss of capacity supervenes; neither desirable nor practically feasible for MAID to be delivered on an emergency basis at the very end of life (might compromise symptom management, palliative sedation as possible alternative); prohibition of advance directives on MAID may lead to pt feeling pressured to request MAID prematurely | **pt**: loss of autonomy as primary reason for request; also wish to avoid burdening others or losing dignity & intolerability of not being able to enjoy one's life; seldom inadequate control of pain or other symptoms; **organizational level**: responsibility for implementing MAID institutional framework and plan for educating staff about engaging in conversations with pts (website, e-learning module); framework to provide equitable access to MAID and to ensure that the substantive procedural safeguards would be upheld; decision made to only offer IV medication administrationat this institution; education and support for staff members directly involved in MAiD, but also for those indirectly involved; designating a specific and consistent location for MAiD delivery has logistical advantages, but distributed model was adopted bc/ no clinical unit was willing to be identified as the "MAID unit"; multidisciplinary MAiD quality committee provides oversight, reports performance, and stewards MAID data for use in quality assessment and research; MAID education must be included in undergraduate medical education curricula and in training for a variety of specialties | In all cases, death within minutes after injection of intravenous drugs for MAID; measures to diminish potential adverse effects of MAiD on the families of the deceased and on staff (e.g., short-term psychosocial support, debriefing sessions for staff before and after MAID | | complete match | |
| 63 | Loggers ET et al | 2013 | Implementing a Death with Dignity Program at a Comprehensive Cancer Center | NEJM, 368(15):1417-24 | 10.1056/NEJMsa1213398 | AS | implementation report (i.e., process description) | to describe the implementation and results of our Death with Dignity program, designed to adhere to legal regulations, maintain safety, and ensure the quality of patient care, at a comprehensive cancer center | Seattle Cancer Care Alliance, the site of care for the Fred Hutchinson–University of Washington Cancer Consortium, a comprehensive cancer center in Seattle that serves the Pacific Northwest | n/a | ENG | USA, Washington (AS legalized) | pts, family members, patient advocate, social workers | | | | pts need to be aware of all options for high quality end of life care; pts need opportunity to have concerns and fears addressed | | AS participants typically have known their oncologist 8 months; good acceptance of AS program | Physicians AS involvement: 36% of physicians willing to consult & prescribe, 26% willing to consult, 38% unwilling or undecided | pts and families grateful after receiving prescription; death described as peaceful; no unexpected complications; one pt had protracted course of dying and died after 1 day taking medication (stressful for family ond HCPs) | | low match | |
| 64 | Marina S et al | 2021 | Views of Psychologists About Their Role in Hastened Death | Omega, 0(0):1–16 | 10.1177/00302228211045166 | VE/AS | cross-sectional quantitative & qualitative survey | to contribute guidance for psychological practice in the context of requests to hasten death. | psychologists | n=499 psychologists (473 from Portugal, 26 from Luxembourg) | ENG | Portugal (VE/AS legalized since 2020), Luxembourg (VE/AS legalized) | psychologist within multiptofessional team; family's doctor, lawyer, nurse, psychiatrist, religious assistant, social worker, specialist doctor of the disease | | | | psychologists can contribute through 1) psychological assessment, 2) psychological support, 3) exploring pt decision-making, and 4) reorientation of the pt; potential functions of psychologists: 1) try to reverse the request, 2) discuss consequences and alternatives to request, 3) evaluate pt's capacity, 4) evaluate consistency of request, 5) support pt in request, 6) write report that legitimizes request for purposes that pt understands, 7) if pt situation does not fit criteria for requesting VE/AS (under law of country), psychologist should advocate for pt, perhaps in legal proceedings, so that their wishes are fulfilled, 8) provide family support | | psychological interventions can enable the pt to make most adequate decision; psychologists' attitudes & roles towards VE/AS differed depending on pt being terminally ill / uncurably non-terminally ill / without physical illness / a terminally ill child; psychologists' attitudes & roles differed between Portugal and Luxembourg; lack of guidelines for psycholgists' roles & training opportunities | | | improve knowledge about psychological approaches regarding VE/AS; explore perspectives of other proffessions on psychological interventions; examine experiences of psychologsits globally in order to establish appropriate training & practice; create networks | partial match | changed from complete match |
| 65 | Matzo ML & Emanual EJ | 1997 | Oncology nurses' practices of assisted suicide and patient-requested euthanasia | Oncol Nurs Forum, 24(10):1725-32 | n/a | VE/AS | cross-sectional quantitative survey | to provide reliable and valid emprical data related to New England Oncology Nursing Society (ONS) members' self-reported practices of AS and VE | oncology nurses, oncology physicians | n=441 oncology nurses; n=180 oncology physicians | ENG | USA, New England (AS not legalized at time of study) | nurses, pt, physician, family members, clergy members, social workers, friends, support group, pt's lawyer, ethics committee, nursing supervisor | | | initiation of the topic by pt | | nurses refer pts to other members of the health care team | nurses and physicians had experience w/ recieving requests for VE/AS (more for AS) | | | differentiate between the effects of gender and profession; further study process of how decisions are made when nurses make referrals; replication of this research with a larger national sample & nurses w/ diverse specialties; insights into attitudes and practices of VE/AS | low match | changed from partial match |
| 66 | Miller DG et al | 2019 | Advance euthanasia directives: a controversial case and its ethical implications | Journal of Medical Ethics, 45(2):84-89 | 10.1136/medethics-2017-104644 | VE (advance directives) | case report | to provide a detailed description of the case, review the major ethical issues it raises and briefly discuss potential weaknesses in the existing regulation of advanced euthanasia directives (AEDs) | pt (w/ Alzheimer's disease) | n=1 pt | ENG | Netherlands (AS/VE legalized since 2002, advance euthanasia directives (AEDs) permitted) | pt w/ Altzheimer's disease and AED, geriatrician (at nursing home), two addtional physicians (one a psychiatrist), family physician, pt family; additionally in case of uncertainty/ambivalence (not used in this case): ethicists, institutional officials, legal authorities | preparation of advanced euthanasia directive; activation of AED; realization of VE | preparation of AED and anticipation of "trigger" point (pt); decision-making competence when preparing advance directives (family physician); interpret AED activation criteria (when is suffering unbearable & right time to execute EAD?) (geritarician & two additional physicians) | rather pt-led during AED preparation; rather physician-led during AED realization | pt wishes, expectations, preferences; AED activation criteria; wishes for hastened death; pt suffering | geriatrician had extended visits and conversations with pt | AED more complicated task than immediate request for VE/AS | pt husband cared for her at home until 6 months before death; then day care; 7 weeks before death, pt full-time at nursing home; pt had previously observed family member deteriorate from dementia & had always said she did not want to experience that; little evidence that pt actually weighed relevant pros & cons of requesting future VE; AED contained notable ambiguities; unclear whether pt wanted AED document alone to satisfy euthanasia request | geriatrician sedated pt without informing her -> state of reduced consciousness -> paramedic inserted infusion line -> geriatrician decided not to administer lidocaine before injecting thiopental because pt 'had scarcely responded to pain stimuli' during the placement of the infusion line -> pt tried to get up during injection of thiopental -> pt's family helped to hold her in place & physician administered rest of thiopental & neuromuscular blocker | | partial match | changed from complete match |
| 67 | Oczkowski SJW et al | 2021 | How we can improve the quality of care for patients requesting medical assistance in dying: A qualitative study of health care providers | Journal of Pain and Symptom Management, 61(3):513-21 | 10.1016/j.jpainsymman.2020.08.018 | MAID | qualitative interview study | to describe MAID clinicians' perspectives on quality of care in MAID, including challenges, successes, and clinical practice suggestions | MAID health care providers | n=20 HCPs (involved in MAID) | ENG | Canada (MAID legalized since 2015) | HCPs, pts, family members | | | | | MAID = new & unique conversation to have w/ pts; careful listening & exploration required; standardized assessment & documentation guides are helpful as conversation guides & educational tools; good communication about MAID requires time | MAID HCPs sometimes asked as consultants, but not part of primary care team; peer support for HCPs essential to identify quality improvement opportunities and provide personal/emotional support; some centers have care coordination services to facilitate the MAID process; HCPs frustrated by challenges pts face when requesting MAID | need for more education on MAID | | further study suggested improvements to facilitate high-quality MAID care; assess challenges for MAID in unique pt groups such as minors, pts w/ psychiatric conditions | low match | changed from partial match |
| 68 | Oczkowski SJW et al | 2021 | How can we improve the experiences of patients and families who request medical assistance in dying? A multi-centre qualitative study | BMC Palliative Care, 20(1):1-12 | 10.1186/s12904-021-00882-4 | MAID | qualitative interview study | to understand patient and family perspectives of quality of care for MAID | pts eligible for MAID (w/ cancer/ALS/cardiovascular dieseases/other neurological diseases); pt family members (spouse, partner, child, sibling, friend) | n(pt)=21; n(family members)=17 | ENG | Canada (MAID legalized since 2015) | pts, family members, health care providers | 1) MAID requests, 2) MAID assessments, 3) preparation for dying, 4) death and aftercare; plus transcendent themes of 5) coordination, 6) patient-centered care | Is MAID the right option? (pt, reflecting w/ family members); come to terms w/ pt's decision for MAID (family members); organ doantion?: eligibility (physician), agreement (pt); location, route, and timing of death, attendees (pt, often in conjunction w/ family and clinical team) | | early exploratory conversations (could be source of reassurance or dissuasion; need for clinicians to acknowledge and explore a pt's wish to die and to address their suffering); supporting pt & family reflection (e.g. reflect on tradeoff btw/avoiding suffering and reduced life); preparing for pt death (e.g. getting affairs in order); option of organ donation; ask repeatedly if pt/family wants to change mind; worry about losing capacity to make decisions (sometimes connected to pain medication and worry about delirium); support pt choice for location, route, and timing of death; concrete practical information about MAID; prepare family & offer support afterwards | 18 themes realted to MAID quality of care: 1) MAID request: prmote awareness, explore request, facilitate written request, move request forward quickly; 2) MAID assessment: in safe & comfortable space, need to build trust, transparent procedural safeguards, patient education (CAVE: pt might feel pressure to perform during assessment); 3) preparation for dying: getting affairs in order, monitor pt status, pt & family reflection, incorporate spiritual/religious/cultural death practices, offer organ doantion (if eligible); 4) death and aftercare: support choice for location/route/timing, choreograph the assisted death, prepare family for death and offer support afterwards; 5) coordination: continuity of care, facilitate access to MAID, education and expectation management; 6) patient-centered care: dignity & privacy, sensitivity to stigma, explore previous experiences w/ death/dying, care for whole family (restoring dignity by providing human connection and compassionate care) | need for compromise and balancing of needs & preferences of pt/family members/clinicians | promote MAID awareness for general community & clinicians | family support after the pt's death | Recalibrate what pt-centered care means in the context of MAID; explore complex triadic decision-making btw/ pt, family members & clinicians; pt & family member satisfaction; testing communication strategies; screening tools for pt; bereavement support for families | complete match | |
| 69 | Onwuteaka-Philipsen BD & Van der Wal G | 2001 | A protocol for consultation of another physician in cases of euthanasia and assisted suicide | Journal of Medical Ethics, 27(5):331-7 | 10.1136/jme.27.5.331 | VE/AS | guideline development (by literature review & consensus) | to develop a protocol for consultation by another physician to support GPs who were going to work as consultant and to endure uniformity | experts (incl/ GPs, psychiatrist, social geriatrician, professor in health law, public prosecutor), GPs (who were going to use the protocol) | n=10 experts, n=10 GPs | ENG | Netherlands (AS/VE legalized since 2002) | | consultation by a 2nd physician | | | | consultant's tasks: talk to pt alone to assess voluntariness of request; take notice of family dynamics & other reasons for request such as fear; is written will sufficient? | 4 important aspects of consultation: independence of the consultant, expertise of the consultant, tasks of the consultant, judgment of the consultant; consultation consists of 4 steps: discussion w/ attending physician, study of medical records, see pt, write written report; is there special reasons for physician not to need to see pt (must be well documented) | | | | low match | |

| | | | | | | | | | | | | | | | | | | | | | | | | |
|---|---|---|---|---|---|---|---|---|---|---|---|---|---|---|---|---|---|---|---|---|---|---|---|---|
| 70 | Onwuteaka-Philipsen BD et al | 2000 | Consultation with another physician on euthanasia and assisted suicide in the Netherlands | Social Science & Medicine, 51(3):429-38 | 10.1016/s0277-9536(99)00476-1 | VE/AS | multi-methods (cross-sectional quantitative survey & qualitative interviews from two studies) | to describe the frequency and characteristics of consultation in cases of VE/AS in the Netherlands; the following research questions were addressed: how often does consultation take place and if it does take place, what is the quality of the consultation, what effect does it have on the physician's decision-making, and have there been changes in the practice of consultation over time? | physicians (GPs, nursing home physicians, diff/ specialist fieds) | approx. n=4,666 questionnaires, n=405 interviews, (n=1,042 GPs from another study for comparison) | ENG | Netherlands (AS/VE legalized since 2002) | 2nd consulting physician, pt, family members, attending physician | consultation by a 2nd physician | are requirements for prudent practice met? (2nd physician) | | | consultation = informal discussion of the case w/ a colleague | criteria for good consultation practice: consultant should not work in same practice or be co-attending physician (independence), should see pt, should (also) consult psychiatrist if pt has psychiatric illness; consultant should draw conclusions re/ pt's condition & estimated life-expectancy, possibility of alternatives, whether pt's request is voluntary, well-considered & persistent | | | | low match | |
| 71 | Pasman HRW et al | 2013 | What happens after a request for euthanasia is refused? Qualitative interviews with patients, realtives and physicians | Patient Education and Counselling, 92(3):313-8 | 10.1016/j.pec.2013.06.007 | VE/AS | qualitative interview study | to obtain in-depth information from both pt and physician perspectives about what happens after a request for VE/AS is refused | pt, pt relatives, physicians | n=9 pts, n=11 physicians, n=3 relatives | ENG | Netherlands (AS/VE legalized since 2002) | pt, physician, family members | | approval of VE/AS request yes or no? (physician) | | | some physicians lacked good communication skills to communicate about wishes for hastened death (which results in suboptimal care and potential harm to pt-physician-relationship) | | | after refusal: mostly not additional communication about VE/AS request, no other plans, no change of physician (although they lost confidence in physician re/ VE/AS, but change too burdensome) | | low match | |
| 72 | Pesut B et al | 2020 | Constructing good nursing practice for medical assistance in dying in Canada: an interpretative descriptive study | Global Qualitative Nursing Research, 7: 1-11 | 10.1177/2333393620938686 | MAID | qualitative interview study | to explore how nurses construct good nursing practice in the context of MAiD | nurses | n=59 nurses | ENG | Canada (MAID legalized since 2015) | pt, nurses | establishing relationship, planning meticulously, orchestrating the MAID death, supporting the family | setting date and time (pt); is pt eleigible? (nurses) | | eligibility assessment by nurses referred to as "legacy conversations" (eligibility assessment at same time as truly listening to pt's story); reasons why pt considers MAID (e.g. current symptom burder, aticipated suffering); planning of the moment of MAID (what do pts want, manage expectations, establish rituals) | nurses offer relationship focused approach; need to establish relationship; humanize the experience and the request; being open to talk about dying and death | some pts do want control that comes w/ being prepared for MAID (but maybe do not actually carry out MAID); need to manage nurses' own anxieties | | | | partial match | 191&192: same data sat, different RQs |
| 73 | Pesut B et al | 2020 | The rocks and hard places of MAiD: a qualitative study of nursing practice in the context of legislated assisted death | BMC Nursing, 19:12 | 10.1186/s12912-020-0404-5 | MAID | qualitative interview study | to better understand the implications of a legislated approach to assisted death for nurses' experiences and nursing practice | nurses | n=59 nurses | ENG | Canada (MAID legalized since 2015) | pt, nurses, physicians, palliative care physicians, family members, IV team members | | | | | | nurse practitioners can be MAID assessors and prescribers; being involved in MAID is highly complex for nurses; team-based MAID care approach helpful/team support (also nurses w/ (palliative care) physicians); sometimes withdrawal of palliative care services after MAID decision has been made (makes good care complicated); leadership support in developing systems to support MAID helpful; sometimes lack of practical guidelines (leading to uncertainty); system need capacity to support MAID | workplace contexts still lack the necessary supports for nurses to confidently meet the precision required of a legislated approach to MAID; lack of palliative care, sufficient providers, a supportive team, practice supports, and a context that allowed nurses to have a range of responses to MAID; nurses perform central role in negotiating initial inquries about MAID, provide important 'wrap-around' care for pts & family | two HCPs being present during MAID was experiences as helpful to support each other; participating in MAID was impactful for nurses & required significant moral work | | partial match | 191&192: same data sat, different RQs |
| 74 | Richards N | 2017 | Assisted suicide as a remedy for suffering? The end-of-life preferences of British "suicide tourists" | Medical Anthropology, 36(4):348-62 | 10.1080/01459740.2016.1255610 | AS | qualitative interview and participant observation study | to provide some insights into what the option of 'going to Switzerland' represents through an analysis of the experiences of people all actively making arrangements to undertake this journey | individuals who applied for AS in Switzerland as foreigners (i.e., patients) | n = 7 individuals | ENG | UK (AS not legal) | pts | | try to get professional assistance in another country or self-deliver/un-assisted suicide? (pt) | | | | often pts did not want to involve family members/physicians; for some, AS offers possibility to share responsibility for the decision; need to reconcile moral issues (e.g. christain belief and suicide, social stigma); AS was seen as a "good death" | | | | low match | |
| 75 | Rodriguez Davila SL et al | 2010 | Management of a request for physician-assisted suicide | American Journal of Hospice & Palliative Medicine, 27(1):63-5 | 10.1177/1049909109341874 | AS | case report | to discuss the need fora comprehensive evaluation to properly assess and manage the untreated physical and emotional suffering that may influence a patient's decision to hasten death | pt (w/ end-stage prostate cancer) | n=1 pt | ENG | USA, Florida (AS not legal) | pt, physician, hospice unit, family | | are psychiatric symptoms couse for AS request? (HCPs); are there alternative treatments to reduce symptom burden? (HCPs) | | psychiatric workup; reasons for AS request; pt's experiences w/ death and dying; symptom reduction; facilitating pt's control over treatment; social worries (wife's welfare after pt's death); worries about loss of dignity | | | longstanding belief in right to AS; several emergency dept. visits and hospital admission immediately prior to request for AS | | need for clear standards of medical and psychiatric evaluation | low match | |
| 76 | Schmahl O et al | 2021 | Request for euthanasia by a psychiatric patient with undetected intellectual disability | BMJ Case Reports, 14(8):e239862 | 10.1136/bc-2020-239862 | VE/AS | case report | to show that extending the VE/AS assessment procedure w/ an independent psychiatric evaluation is important | pt (68 yrs, diagnosed with bipolar I disorder) | n=1 pt | ENG | Netherlands (AS/VE legalized since 2002) | pt, psychiatrist, physician, family members | independent psychiatric assessment; optimization of psychiatric and somatic treatment | | | alternative views on diagnoses; alternative treatments; other possibilities for finding peace and rest within the framework of life review | | independent psychiatric assessment recoemmended | | | | low match | |
| 77 | Schmid M et al | 2016 | Medical end-of-life decisions in Switzerland 2001 and 2013: Who is involved and how does the decision-making capacity of the patient impact? | Swiss Medical Weekly, 146(0910):w14307 | 10.4414/smw.2016.14307 | VE/AS, (also other medical end-of-life decisions) | cross-sectional quantitative study (two death certificate studies) | to compare the results of the 2001 and 2013 studies with a special focus on shared decision-making and patients' decision-making capacity | attending physicians | 2013: n=3,173 questionnaires; (comaprison w/ 2001: n=3,355 questionnaires) | ENG | Switzerland (AS legalized) | pt, family members, physician | | | shared decision-making/high level of pt/family involvement for end-of-life decisions | end-of-life issues/practices/options | end-of-life decisions are most often discussed w/ pt and/or family members; involvement of pt depends on pt's decision-making capacity | | | | analyze social disparities' (e.g. educational level) connection to lack of involvement in end-of-life decision-making | low match | |
| 78 | Schuurmans | 2020 | Supporting GPs around euthanasia requests from people with dementia: a qualitative analysis of Dutch nominal group meetings | British Journal of General Practice, 70(700):E833-42 | 10.3399/bjgp20X713093 | VE | qualitative consensus study (using the nominal group technique) | to identify and prioritise forms of support that should be used or implemented to help GPs confronted with euthanasia requests frompts w/ dementia | experts (incl/ Dutch care physicians for older people, GPs, legal experts, a healthcare chaplain, a palliative care consultant, a psychologist) | n=15 experts (8 in the 1st meeting, 7 in 2nd meeting) | ENG | Netherlands (AS/VE legalized since 2002) | physicians, pts, other HCPs | | VE yes/no for a pt w/ dementia (physician) | | give good information abour possibilities/impossibilities | need for communication skills (e.g. specialised conversational techniques, discussing end-of-life issues, conducting family meetings) | "buddy system" to support HCPs (by e.g. end-of-life clinic physician or nurse, care physician for older people, SCEN physician, spiritual care provider); team approach/support (also through organisations); multidisciplinary team meetings (e.g. interdisciplinary consultation, moral case deliberation) | community activities to shape public opinion/educate the public | impact of VE participation on physicians | compare the present qualitative research findings on end-of-life care for persons with dementia across other countries, as cultural, medical, and legislative differences may result in different end-of-life care questions in dementia care | low match | |
| 79 | Sellars M et | 2022 | Medical practitioners' views and experiences of being involved in assisted dying in Victoria, Australia: A qualitative interview study among participating doctors | Social Science & Medicine, 292:114568 | 10.1016/j.socscimed.2021.114568 | AS | qualitative interview study | to describe the perspectives and experiences of doctors who had provided AS during the first 12-months of its operation in Victoria, Australia and to understand how they conceptualized their professional roles in the practice of providing AS and inform ways to optimize health systems implementing AS | physicians (who had been involved in the AS process) | n=32 physicians | ENG | Victoria, Australia (AS legalized since 2019) | pt, physicians, AS navigators, AS pharmacists | | Do I want to be involved in AS? (physician); is prognosis less than 6 months? (physician) | pt intitate AS discussion | pt communicate preferences to physicians & family (incl/ what is "vital" to pt); be explicit about prognosis; pts have to explicitly consent to AS (as opposed to other end-of-life practices); clarify pts' intent; assess understanding of AS and cognitive capacity; inform about alternatives; make sure that pt not coerced; set realistic expectations (e.g. timeframe until when medication can be dispensed); inform about need for pt to have capacity to make decisions; provide emotional support to pt & family (alleviate worry, guide through process) | AS as intimate, "holistic" and person-centered approach to care; remaining uncertainty re: prognosis and freedom from coercion | having a relationship w/ pt made AS support easire for physicians (would not support unknown pts); AS process can foster intimate relationship btw/ pt, physician & family; family might disagree w/ pt decision to seek AS; diff/ opinions on whether physician should have to be present for AS moment; team support helpful; AS navigators and pharmacists as helpful mentors and support structure; AS process needs time from physicians; personal, interpersonal, structuralk resources needed | | AS as strange and challenging, but also rewarding experience for physicians; need to recognize role-based and team-based challenges; potential for personal, social, financial impact on physicians; involvement in AS experienced as being consistent w/ being a "good doctor"; need for administrative and emotional/wellbeing support during AS for physicians; support for family members during AS moment; AS might imporve bereavement process for family members (due to planned and calm death) | | partial match | |
| 80 | Shaw J et al | 2021 | Perceptions and experiences of medical assistance in dying among illicit substance users and people living in poverty | Journal of Death & Dying, 84(1):267-88 | 10.1177/0030222819889827 | MAID | qualitative interview study | to explore the beliefs and experiences of people who are marginalized in relation to MAID; to understand what questions, concerns, and desires they have about the availability of MAID and other aspects of end-oflife care; and to continue to develop a research partnership network with community members as a basis for future collaboration | individuals who were service users, peer community leaders, and formal service providers | n=46 individuals (27 service users, 12 peer community leaders, 7 formal service providers) | ENG | Canada (MAID legalized since 2015) | pts, physicians, community health services/community service providers<br>marginalized people | | | | | need of knowing where to go/having someone to talk to about MAID considerations; fear of being sent to psychiatry/involuntary hold when inquiring about MAID | lack of knowledge about end-of-life practices/options; stigma/lack of autonomy as barriers to end-of-life options; some marginalized ppl would prefer suicide to MAID; importance of family/friends/community at end of life; power imbalance & lack of trust between marginalized ppl & HCPs; need to be aware of socioeconomic factors influencing healthcare decisions | need to provide MAID services also in remote communities | | address best practices for knowledge dissemination and ways of decreasing stigma in the health-care system; explore experiences of end-of-life care of other cultural groups whose experiences were not captured in these interviews; investigate Indigenous ways of knowing at the end of life using participatory, decolonizing methodologies | low match | |
| 81 | Snijdewind M | 2014 | Complexities in euthanasia or physician-assisted suicide as perceived by Dutch physicians and patients' relatives | Journal of Pain and Symptom Management, 48(6):1125-34 | 10.1016/j.jpainsymman.2014.04.016 | VE/AS | qualitative interview study | to identify and categorize the characteristics of VE/AS requests that are more complex than others | physicians, family members (of patients who had died by VE/AS) | n=54 ppl (28 physicians, 26 family members) | ENG | Netherlands (AS/VE legalized since 2002) | pts, family members, physicians, HCPs | | Is VE/AS organizable in the timeframe available? (HCPs) | | | possibility for miscommunication due to different implicit expectations | VE/AS request can be surprising (physicians & family members often did not realize severity of suffering); VE/AS should be regarded as a process (not isolated event); VE/AS decision-making needs time (to grow, to develop or deepen relationship, to balance different options); importance of involving family (important for most physicians; need for coping capacity of family members); VE/AS process as process of growth where all involved grow together; need for pre-established physician-pt-relationship; physicians sometimes feel pressured by pt/family to perform VE/AS; is it ethical to reject VE/AS requests because a case is complex? | | | quantitative research to show how often these unusually complex situations occur and whether there is a need for additional education or guidelines to handle these complex aspects | partial match | 246&247: same physician sample, different RQs |

| | | | | | | | | | | | | | | | | | | | | | | | | |
|---|---|---|---|---|---|---|---|---|---|---|---|---|---|---|---|---|---|---|---|---|---|---|---|---|
| 82 | Snijdewind MC et al | 2018 | Developments in the practice of physician-assisted dying: perceptions of physicians who had experience with complex cases | Journal of Medical Ethics, 44(5):292-6 | 10.1136/medethics-2016-103405 | VE/AS | qualitative interview study (secondary analysis) | to see if and how physicians who had experience with complex cases of VE/AS perceive any important developments in the practice of VE/AS and to further explore these possible developments | physicians | n=28 physicians | ENG | Netherlands (AS/VE legalized since 2002) | pts, family members, HCPs, physicians | | | | | pts have to voice themselves that they want VE/AS | diff/ perceptions of VE/AS between physicians (difficult, exceptional) and pts/family members (easy); physicians sometimes feel pressured to perform VE/AS; acceptable reasons for VE/As have broadened over time | | | | low match | 246&247: same physician sample, different RQs |
| 83 | Starks H et al | 2007 | Family member involvement in hastened death | Death Studies, 31(2):105-30 | 10.1080/07481180601100483 | AS, (also other forms of hastened death incl/ euthansia & suicide) | qualitative interview study (secondary analysis) | to characterize what participation in a hastened death adds to typical family caregiving responsibilities at the end of life and the subsequent positive and negative consequences that family members experience as a result of their participation | family members | n= 48 family members (28 reporting on 23 deceased pts, 20 reporting on 12 pts from ongoing cases) | ENG | USA, Washington & Oregon (AS leagl in Oregon and illegal in Washington at time of study) | pts, family members (caregivers), physicians, HCPs, advocacy organizstions | | | | | | important to recognize and address family members needs; pts rely on family members to seek information about how to receive AS/hastened death; where AS is illegal, obtaining information for HCPs is even more challenging (open requests might lead to undesired outcomes); meeting legal requirements for AS can be challenging & burdensome; sometimes impression physicians were taking longer than neccessary ("draggingtheir feet") | | knowing the date of the death makes closure possible; hastened death can be experienced as a bonding experience for family; professional support can reassure family members and increased their comfort with their role and involvement; complications in dying process can delay bereavement process; maintaining secrets about the hastened death can isolate family members; worries about prosecution when AS is not legal | | low match | changed from partial match |
| 84 | Stewart DE et al | 2018 | Consultation-liaison psychiatry and physician-assisted death | General Hospital Psychiatry, 55:15-9 | 10.1016/j.genhosppsych.2018.08.011 | MAID | description of one organization's processes (incl/ requirements, team experience, roles/issues/concers, pt examples) | to describe our Canadian AS experience over 2 years, highlighting the role of consultation-liaison psychiatrists, at a large hospital network that provides MAID | hospital | n=1 hospital | ENG | Canada (MAID legalized since 2015) | | | is pt eleigible? (MAID assessment team); can MAID accomodated at hospital or rather at home? (MAID team) | | reasons for MAID request (e.g. high preference for personal control); personal experiences w/ deaths and dying of family/friends; try to accomodate pt's wishes for contextual factors (e.g. whom to be present, what to wear, flowers, music); inform about psychiatric tretament options and assess capacity to give informed consent (if pt has comorbid psychiatric disorder) | extensive discussion between MAID team and other specialties caring for pt to help understand and differetiate between pt's and own values and opinions; family meeting can help to resolve conflict btw/ pts and families | some pts worried about losing capacity to make decisions; discerning whether perceived suffering of family & friend is undue influence is challenging; sometimes family&friends oppose MAID; physicians w/ conscientious objection do not have to participate, but to refer pt (list of willing providers) | | provide short-term support for family members; grief seems to be more about loss of loved-one than about MAID | | partial match | |
| 85 | ten Cate K, van Tol DG &van de Vathorst S | 2017 | Considerations on requests for euthanasia or assisted suicide: a qualitative study with Dutch general practitioners | Family Practice, 34(6):723-9 | 10.1093/fampra/cmx041 | VE/AS | qualitative interview study (secondary analysis) | to better understand the considerations that play a role when physicians decide on a request for VE/AS; RQ: what considerations play a role in practice when Dutch GPs have to decide on an VE/AS request? | physicians | n= 33 physicians (from two sets of interviews) | ENG | Netherlands (AS/VE legalized since 2002) | pts, family members, physician | | whether to support the request? (physician) | pt initiate VE/AS request | | VE/AS communication takes time (sometimes multiple conversations between pt and physicians, potentially daily) | diff/ GPs have diff/ opinions on what unbearable suffering entails (e.g. not acceptable if pt can still walk, talk, eat; only acceptable in terminal phase of illness); family members play important role in decision-making process; sometimes family does not agree w/ request; in case of unresolved conflicts w/in family, help pts find clusore first | | | study how physicians' attitudes/opinions influence VE/AS decision-making (especially re: consideration unrelated to legal requirements); add quantitative assessment to assess relation btw/ respondents' characteristics and degree to wich physicians' own opinions influence VE/AS decision-making; expland scope to other countries and other end-of-life-practices | partial match | |
| 86 | van Wesemael Y et al | 2010 | Consulting a trained physician when considering a request for euthanasia: an evaluation of the process in Flanders and the Netherlands | Evaluation & the Health Professions, 33(4):497-513 | 10.1177/0163278710370605 | VE | cross-sectional quantitative study (two death certificate studies) | to evaluate and compare the consultations of the Flemishand Dutch euthanasia consultation services | attending physicians, specialized VE consultants | n=569 questionnaires (96 Belgian VE consultants, 40 Belgian attending physicians, 433 Dutch VE consultants, 433 Dutch attending physicians) | ENG | Belgium (VE legalized) & Netherlands (AS/VE legalized since 2002) | | consultation by a 2nd physician | | | most frequently discussed: pt's unbearable suffering, hopelessness of medical situation, well-considered nature of request; least frequently discussed: alternatives (esp. in Blegium) | consultants mostly not co-attendings and work in diff/ institution; consultants often do not know pt; physicians often communicate w/ each other via telephone, less frequently face-to-face; ; diff/ between Belgium / Netherlands re/ what is disussed btw/ physicians (re/ alternatives, unbearable suffering); only in the Netherlands there are checklists what physicians should talk about; consultants talk to pt alone or w/ family present; consultants talk an average of 3 to 4 hrs w/ pts | attending physicians often already decided before contacting consultant; legal requirements for consultants (Belgium: read medical file, examine pt, ascertain that suffering is persistent & unbearable, produce written report; Netherlands: see pt, formulate written advice covering due care requirements for VE (i.e. voluntariness & well-considered nature of request, hopeless & unbearable suffering of pt, lack of any reasonable clinical alternative)) | | | | partial match | |
| 87 | Verhofstadt et al | 2019 | Psychiatric patients requesting euthanasia: Guidelines for sound clinical and ethical decision making | International Journal of Law and Psychiatry 2019 Vol. 64 Pages 150-161 | 10.1016/j.ijlp.2019.04.004 | VE | document analysis | to critically reflect five initiatives offering guidance on VE requests by psychiatric patients, focusing on all legal due care criteria in order to identify: 1) proposed measures to operationalise the evaluation of the legal criteria; 2) suggestions of additional safeguards going beyond these criteria; 3) remaining fields of tension | guideline documents | n=5 guidelines | ENG | Belgium (VE legalized) | pts, family members, attending/treating physician, consulting physicians (psychiatrists), HCPs, religious official | guidelines for psychiatric pts include 1) positive assessments by at least 2 psychiatrists, 2) a-priori evaluation system; 3) two-track approah w/ simultaneous focus on VE request and continuing treatment (by same of diff/ providers) | VE approval yes/no? (physician) | | underlying reasons for VE request; try to understand motives and clarify if pt does not want to involve family members | not all physicians (attending physician, 2 consulting physicians) need to assess all aspects to the same extent (2nd consulting physician assesses strictly due-care criteria to avoid risk of bias due to (counter)transference); involvement of family members/important others (interpretative dialogue, oppotunity to strengthen relationships, does not mean that they have to agree w/ request); | guidelines support VE process by providing information and operationalizations of legal criteria; guidelines for VE requests by psychiatric pts go beyond legal requirements; strict differentiation between attending and consulting physician not always applicable in routine care; establish interdisciplinary evaluation committee coordinated by chief physician incl/ 1) in-house staff members (attending physician, dept. chief physician, 2 other staff members (at least one non-physician)), 2) at least two colleagues w/ expertise in VE in psychiatry context (not in-house employed), 3) at least two outside ppl w/ expertise in VE topic | | appropriate aftercare | establish prospective longitudinal regstry for Ve for psychological suffering; collaboration and systematic research between government VE institutions and research institutions to assess interpretation/operationalization of legal requirements & potential problems/complexities of review system | low match | |
| 88 | Wiebe et al | 2021 | Assessment of capacity to give informed consent for medical assistance in dying: a qualitative study of clinicians' experience | CMAJ Open, 9(2):E358-63 | 10.9778/cmajo.20200136 | MAID | qualitative interview study | to explore how clinicians assessed capacity in people requesting MAID, what challenges they had encountered and what tools they used | MAID assessors/providers (diff/ specialties) | n=20 MAID assessors/providers | ENG | Canada (MAID legalized since 2015) | pt, physician | | | | | assessment of capacity to make decision mostly through conversation and w/o formal assessment tool; assessment of capacity to make decisions challenging w/ pts who are nonverbal or tire too quickly to have lengthy conversations, w/ fluctuating capacity, w/ early dementia; assesors try to foster conditions for pts to demonstrate capacity; secondary assessment through colleagues or family members | | | | | low match | |
| 89 | Health Canada | 2019 | First annual report on medical assistance in dying in Canada | n/a | n/a | MAID | annual report w/ case reports | | pts | | | Canada (MAID legalized since 2015) | pt, family members, physician | | | | | | most pts either received palliative care or had access to palliative care w/o using it | vast majority of MAID providers were GPs, less frequemtly palliative care physicians & anesthesiologists | MAID was carried out primarily in hospitals or ptivate homes, less frequently on palliative care units or in residential care | | low match | |
| 90 | Health Canada | 2020 | Second annual report on medical assistance in dying in Canada | n/a | n/a | MAID | annual report w/ case reports | | pts | | | Canada (MAID legalized since 2015) | pt, family members, HCPs | | | | | | most pts either received palliative care or had access to palliative care w/o using it | vast majority of MAID providers were physicians, less frequently nurse practitioners | MAID was carried out primarily in hospitals or ptivate homes, less frequently on palliative care units or in residential / long-term care | | low match | |
| 91 | Health Canada | 2021 | Third annual report on medical assistance in dying in Canada | n/a | n/a | MAID | annual report w/ case reports | | pts | | | Canada (MAID legalized since 2015) | pt, family members, physician | | | | | | | increase in number of MAID providers and increasing provision by nurse practitioners | shift from hospital setting to private homes for MAID | | low match | |
| 92 | Regional euthanasia review committees RTE | 2002 | 2002 annual report | n/a | n/a | VE/AS | annual report w/ case reports | | pts | n=2 pts | | Netherlands (AS/VE legalized since 2002) | pt, physician, family members (children), home carer, specialised home care team, lung specialist, VE/AS physician | pt makes request (repetedly w/ diff/ ppl, written); GP talks to other providers; GP contacts independant VE/AS physician for assessment of due care criteria | | pt initiated VE/AS request | sufficient information on situation & prospects; alternative treatments | pt voiced VE/AS request often to several HCPs & family members | family members backed pts request | | post-VE/AS review committee | | low match | |
| 93 | Regional euthanasia review committees RTE | 2003 | 2003 annual report | n/a | n/a | VE/AS | annual report w/ case reports | | pts | n=3 pts | | Netherlands (AS/VE legalized since 2002) | pt, physician, nurse, family members, VE/AS physician, GP, psychiatrist | pt makes request (repetedly w/ diff/ ppl, written, potentially advance directive); attending physician contacts independent consulting VE/As physician; consulting physician assesses due care criteria (soemtimes several consultations w/ pt) | | pt initiated VE/AS request | is suffering unbearable?; are there any treatment alternatives or prospect of improvement? | pt repeated request w/ diff/ ppl | family members sometimes opposed to VE/AS request (may change over time) | | post-VE/AS review committee | | low match | |
| 94 | Regional euthanasia review committees RTE | 2004 | 2004 annual report | n/a | n/a | VE/AS | annual report w/ case reports | | pts | n=3 pts | | Netherlands (AS/VE legalized since 2002) | pt, physician, family members, VE/AS physician, psychologist, nursing home doctor, geriatric psychiatrist, independent physicians | pt makes request; attending physician contacts independent consulting VE/As physician; consulting physician assesses due care criteria | | pt initiated VE/AS request | adequate information on situation & prospects | family members sometimes involved in consultations w/ consulting physicians | even if consulting physicians are uncertain about due care criteria or see them not fulfilled, attending physicians sometimes grant VE/AS requests (sometimes w/ additional consultation w/ specialists or another consultation w/ the first consulting physician); sometimes pts want to know that VE/AS is an option for them, but do not seek it immediately | | post-VE/AS review committee; review committee might inquire if granting and aadministration of VE/AS raises questions | | low match | changed from partial match |
| 95 | Regional euthanasia review committees RTE | 2005 | 2005 annual report | n/a | n/a | VE/AS | annual report w/ case reports | | pts | n=2 pts | | Netherlands (AS/VE legalized since 2002) | pt, physician, independent VE/AS physician/GP, family members (parents), paediatrician | pt makes request (repeatedly w/ diff/ ppl); attending physician contacts independent consulting VE/As physician; consulting physician assesses due care criteria | | pt initiated VE/AS request | assess if due care criteria are met during communication w/ pt | involvement of family members; long conversation w/ family members, physician, other HCP about VE/AS request (3hrs) | for minor, partents consented to procedure; time course of decision-making sometimes brief (due to clinial situation of pt) | | post-VE/AS review committee | | low match | changed from partial match |

| | | | | | | | | | | | | | | | | | | | | | |
|---|---|---|---|---|---|---|---|---|---|---|---|---|---|---|---|---|---|---|---|---|---|
| 96 | Regional euthanasia review committees RTE | 2006 | 2006 annual report | n/a | n/a | VE/AS | annual report w/ case reports | pts | Netherlands (AS/VE legalized since 2002) | pt, physician, independent VE/AS physician, geriatric psychiatrist, family members (wife) | pt makes request (repeatedly); attending physician contacts independent consulting VE/As physician; consulting physician assesses due care criteria | are due care criteria met? (attending physician, consulting physician); is request cause by depression or unbearable suffering? (physician) | pt initiated VE/AS request | adequate information on situation & prospects; reasons for unbearable suffering; no alternative treatments available | time needed (sometimes daily or twice daily visits w/ pts in days before VE/AS) | GP & consulting physician may communicate w/ pts during decision-making process & other physician asministers lethal medication; possibility for advance directives | | post-VE/AS review committee | | low match | changed from partial match |
| 97 | Regional euthanasia review committees RTE | 2007 | 2007 annual report | n/a | n/a | VE/AS | annual report w/ case reports | pts | Netherlands (AS/VE legalized since 2002) | pt, physician, independent VE/AS physician, geriatric psychologist, clinical geriatrician (=physician) | pt makes request (repeatedly); attending physician contacts independent consulting VE/As physician; consulting physician assesses due care criteria | is suffering palpable and unbearable? (physician) | pt initiated VE/AS request | what does pt's daily life look like?; what suffering does pt experience? | some pts physically very restricted (e.g. atrophied muscles) w/ hard time to talk | | | post-VE/AS review committee | | low match | changed from partial match |
| 98 | Regional euthanasia review committees RTE | 2008 | 2008 annual report | n/a | n/a | VE/AS | annual report w/ case reports | pts | Netherlands (AS/VE legalized since 2002) | pt, physician, family member (husband), second physician | pt makes request (repeatedly); attending physician contacts independent consulting VE/As physician; consulting physician assesses due care criteria | | pt initiated VE/AS request | | | second physician has to be independent from attending physician | | post-VE/AS review committee | | low match | changed from partial match |
| 99 | Regional euthanasia review committees RTE | 2009 | 2009 annual report | n/a | n/a | VE/AS | annual report w/ case reports | pts | Netherlands (AS/VE legalized since 2002) | pt, physician, independent VE/AS physician, family members (wife, children), psychiatrist, neurologist, nursing staff, hospital management, trainee doctor on service, attending specialist | pt makes request (repeatedly); attending physician contacts independent consulting VE/As physician; consulting physician assesses due care criteria | | pt initiated VE/AS request | | some pts have limited ability to talk (aphasia) | pts may be worried of losing capacity to make decisions; in pts w/ dementia, greater caution must be exercised in responding to any request for VE/AS; lack of time (due to severity of condition & suffering of pt) may prevent physician from involving independent physician for assessment; independent VE/AS assessment can be time-consuming and intensive (multiple visits in short time) | | post-VE/AS review committee | | low match | changed from partial match |
| 100 | Regional euthanasia review committees RTE | 2010 | 2010 annual report | n/a | n/a | VE/AS | annual report w/ case reports | pts | Netherlands (AS/VE legalized since 2002) | pts, physician, family members (wife and children), psychiatrist, geriatrician, independent VE/AS physician | pt makes request (repeatedly); attending physician contacts independent consulting VE/As physician; consulting physician assesses due care criteria | | pt initiated VE/AS request | adequate information on situation & prospects; experience of pts' family members/loved ones having suffered from a disease | experience of pts' family members/loved ones having suffered from a disease might influence their view on the prospects; some pts have limited ability to talk; advance directives and secondary accounts from HCPs & family members can substantiate VE/AS request if pt has limited ability to express themselbes | pts may be worried of losing capacity to make decisions; ; possibility for advance directives | | post-VE/AS review committee | | low match | changed from partial match |
| 101 | Regional euthanasia review committees RTE | 2011 | 2011 annual report | n/a | n/a | VE/AS | annual report w/ case reports | pts | Netherlands (AS/VE legalized since 2002) | pt, physician, independent VE/AS physician, geriatrician, family members (husband, wife) | pt makes request (repeatedly); attending physician contacts independent consulting VE/As physician; consulting physician assesses due care criteria | is unbearable suffering plapable to physician? (physician) | pt initiated VE/AS request | sufficient information about situation & prognosis; medical history; what effect does medical diagnosis have on pt; are there treatment alternatives?; circumstances in which pt would want VE/AS; life history & personality aspects | | if VE/AS is not administered within 6 weeks of independent assessment, new independent assessment needed; | | post-VE/AS review committee; physician might be invited for personal interview by committee, if committee lacks information about the case | | low match | changed from partial match |
| 102 | Regional euthanasia review committees RTE | 2012 | 2012 annual report | n/a | n/a | VE/AS | annual report w/ case reports | pts | Netherlands (AS/VE legalized since 2002) | pt, physician, nurse, family members, carers, external expert | pt makes request (repeatedly); attending physician contacts independent consulting VE/As physician; consulting physician assesses due care criteria | | pt initiated VE/AS request | sufficient information on situation & prospects; no alternatives; unbearable suffering | some pts have limited ability to talk; experience of pts' family members/loved ones having suffered from a disease might influence their view on the prospects; time-intensive communicatin needed btw/ pt, family members, diff/ physicians | requests by pt w/o capacity to make deicisons can be granted if advance directive exists; GP did not feel comfortable w/ administering VE/AS himself, pt contacted end-of-life clinic to assess and provide VE/AS (new physician needs to become sufficiently aquainted w/ pt & contact independent VE/AS physician); | | post-VE/AS review committee | | low match | changed from partial match |
| 103 | Regional euthanasia review committees RTE | 2013 | 2013 annual report | n/a | n/a | VE/AS | annual report w/ case reports | pts | Netherlands (AS/VE legalized since 2002) | pts, physician, independent VE/AS physician, nursing assistant, psychiatrist, mental health nurse | pt makes request (repeatedly); attending physician contacts independent consulting VE/As physician; consulting physician assesses due care criteria | is communication sufficiently independet? (in case of limited ability to talk and limited motor skills) (physician) | pt initiated VE/AS request | sufficient information on situation & prospects; no alternatives | some pts have limited ability to talk (incl/ limited motor skills) | | | post-VE/AS review committee | | low match | changed from partial match |
| 104 | Regional euthanasia review committees RTE | 2014 | 2014 annual report | n/a | n/a | VE/AS | annual report w/ case reports | pts | Netherlands (AS/VE legalized since 2002) | pt, psychologist, attending physician/GP, psyhiatrist, clinical geriatrician, independent VE/AS physician (sometimes more than 1), family members | pt makes request (repeatedly); attending physician contacts independent consulting VE/As physician; consulting physician assesses due care criteria | | pt initiated VE/AS request | reasonable alternative to the pt's situation; prospect of improvement | couple request: who communicates how much w/whom? (need for independent assessments?); talk to pt alone to avoid influencing by others; potentially need for more independent assessments in complex situations (e.g. psychiatric problems) | pt requested VE/AS after wife died and he saw no meaning & quality in life afterwards; especially challenging, when pt's request is connected to partner seeking VE/AS (two independent assessment by independent physicians needed); GP/attending physician did not approve VE/AS request, pt contacted end-of-life clinic to assess and provide VE/AS; family members backed pt's (w/ psychiatric disorder) request | | post-VE/AS review committee | | low match | changed from partial match |
| 105 | Regional euthanasia review committees RTE | 2015 | 2015 annual report | n/a | n/a | VE/AS | annual report w/ case reports | pts | Netherlands (AS/VE legalized since 2002) | pt, family members, GP, geriatrician, HCPs, independent VE/AS physician (sometimes more than 1) | pt makes request (repeatedly, sometimes incl/ advance directives); attending physician contacts independent consulting VE/As physician; consulting physician assesses due care criteria | | pt initiated VE/AS request | | some pts have limited ability to talk (aphasia) | couple w/ two diff/ attending physicians & one independent physician | | post-VE/AS review committee | | low match | changed from partial match |
| 106 | Regional euthanasia review committees RTE | 2016 | 2016 annual report | n/a | n/a | VE/AS | annual report w/ case reports | pts | Netherlands (AS/VE legalized since 2002) | pts, family members, physicians, independent VE/AS physician (sometimes more than 1) | pt makes request (repeatedly, sometimes incl/ advance directives); attending physician contacts independent consulting VE/As physician; consulting physician assesses due care criteria | | pt initiated VE/AS request | sometimes pts voice ambiguous/changing wishes (wanting to die vs. not yet) | | physicians might reject involvement in VE/AS | | post-VE/AS review committee | | low match | changed from partial match |
| 107 | Regional euthanasia review committees RTE | 2017 | 2017 annual report | n/a | n/a | VE/AS | annual report w/ case reports | pts | Netherlands (AS/VE legalized since 2002) | pt, family members, HCPs | pt makes request (repeatedly, sometimes incl/ advance directives); attending physician contacts independent consulting VE/As physician; consulting physician assesses due care criteria | | pt initiated VE/AS request | | | sometimes repeated visits of independent VE/AS physician; physician rejected involvement in VE/AS because she did not deem suffering unbearable & felt pressured by pt; sometimes if attending physicians reject VE/AS request, pts seek other physician from specialized VE/AS clinic | | post-VE/AS review committee | | low match | changed from partial match |
| 108 | Regional euthanasia review committees RTE | 2018 | 2018 annual report | n/a | n/a | VE/AS | annual report w/ case reports | pts | Netherlands (AS/VE legalized since 2002) | pt, physician, GP, psychiatist, mental health nurse, case manager, family members, mental health service team | pt makes request (repeatedly); attending physician contacts independent consulting VE/As physician; consulting physician assesses due care criteria | | pt initiated VE/AS request | palpable unbearable suffering | | in case of suicidaility include psychiatrist; sometimes if attending physicians reject VE/AS request, pts seek other physician from specialized VE/AS clinic | | post-VE/AS review committee | | low match | changed from partial match |
| 109 | Regional euthanasia review committees RTE | 2019 | 2019 annual report | n/a | n/a | VE/AS | annual report w/ case reports | pts | Netherlands (AS/VE legalized since 2002) | pts, family members, physicians, indipendent physicians, psychiatrist, geriatrician, nurses, notary | pt makes request (repeatedly); attending physician contacts independent consulting VE/As physician; consulting physician assesses due care criteria | | pt initiated VE/AS request | | personal life experiences of pts might shape their wish for VE/AS; some pts have limited ability to talk | sometimes several physicians consult on one case; if attending physicians are not able/willing to assist w/ request or if case is too complex, pts are referred to specialized VE/AS clinic; especially challenging if ppl are incarcerated/in forensic psychiatric hold | | post-VE/AS review committee | | low match | changed from partial match |
